# Population-based Normative Reference for Retinal Microvascular Atlas

**DOI:** 10.1101/2024.10.25.24316087

**Authors:** Mayinuer Yusufu, Algis J. Vingrys, Xianwen Shang, Lei Zhang, Danli Shi, Nathan Congdon, Mingguang He

## Abstract

**Objective:** To establish the normative range of a comprehensive set of retinal vascular measurements to better understand their value as biomarkers for assessing ocular and systemic health.

**Methods:** This cross-sectional study used data from the UK Biobank. Retina-based Microvascular Health Assessment System (RMHAS) software was used to extract retinal vascular measurements, including Calibre, Complexity, Density, Branching Angle, and Tortuosity, differentiating between arteries and veins, and between the macula and retinal periphery. In addition, we explored relationships between those measurements and health metrics, including age, systolic blood pressure (SBP), body mass index, glycated hemoglobin, and intraocular pressure.

**Results:** Among 10,151 healthy participants, we reported a normative range for 114 retinal vascular measurements, stratified by sex and age. The mean values of Central Retinal Artery Equivalent (CRAE) and Central Retinal Vein Equivalent (CRVE) were 152 (standard deviation=14.9) μm and 233 (21.5) μm respectively. The mean value of Fractal Dimension (FD) was 1.77 (0.032), with arterial FD 1.53 (0.039) and venular FD 1.56 (0.025). Age and SBP showed the strongest associations with most retinal parameters among health metrics. CRAE, CRVE, Density, and Complexity decreased with increasing age and SBP. Changes in arterial measurements with age and SBP were generally greater than for venous measurements. Generalized Additive Models further revealed that observed associations were mainly linear.

**Conclusions:** By establishing population normative data for a comprehensive set of retinal vascular measurements, our study enables quantifiable approaches to better understand retinal vascular changes.

## Introduction

The retina is unique as a segment of the vascular system permitting direct, non-invasive visualization. It has been widely studied to understand the link between ocular manifestations and systemic disease since the invention of direct ophthalmoscopy in 1851^1^. While previous research has devoted considerable attention to studying the retinal vascular network, and how it reflects systemic health, the scope of work has been limited by the labour-intensive nature of manual labeling. In recent decades, advances in artificial intelligence (AI) have accelerated progress in assessing systemic health through characterizing the retinal vascular geometry.^2^

Increasing evidence shows that the retinal vascular features are indicative of various systemic diseases, including hypertension, diabetes, cardiovascular and neurodegenerative diseases, and even mortality risk^3-5^. The convenience, non-invasive nature, and affordability of fundus photography make it a valuable tool for risk assessment in these and other health conditions, allowing more timely intervention. Population normative data on retinal vascular features provides a valuable foundation for studying retinal vascular alterations.

Existing studies, generally small, report population normative data on a limited range of retinal vascular features, such as calibre, fractal dimension, and vascular density^6,7^. This limits their usefulness for a comprehensive assessment of systemic health.

The current study addresses this gap by establishing a reference database of retinal vascular parameters derived from fundus photographs in a large population cohort, the UK Biobank^8^. Using the Retina-based Microvascular Health Assessment System (RMHAS)^9^, a deep learning model, we report on the gender-stratified normative ranges of 114 arterial and venous measurements in the macula and peripheral retina for five geometric features of the retinal vascular network: including vascular Calibre, Complexity, Density, Branching Angle, and Tortuosity.

## Methods

### Study design

We used de-identified data from the UK Biobank, a large population-based cohort study including participants aged 40 to 69 years. The study was launched in 2006 in the United Kingdom, and introduced ocular examinations, including fundus photography, in 2009. The details of the study have been described elsewhere^8^.

### Ethical consideration

The current study utilized data from the UK Biobank, which obtained ethical approval from the North West Multi-Centre Research Ethics Committee (reference number 06/MRE08/65). Our study was conducted under UK Biobank project application #94372 and adhered to their ethical guidelines and data access protocols.

### Inclusion and exclusion criteria

We included non-smoking participants with a body mass index <=30 (kg/m^2^) who identified themselves as of white ethnicity. Those with a self-reported history of ocular disease, diabetes, cardiovascular or neurodegenerative disease, intraocular pressure (IOP) ≥ 21 mmHg, systolic blood pressure ≥ 140 mmHg, or diastolic blood pressure ≥ 90 mmHg were excluded.^6,10^ Table S1 details definitions of variables used in the current study.

### Retinal vascular network analysis

The retinal fundus images were taken in a dark room without pupil dilation using using Topcon 3D OCT-1000 Mark II system^11^. The image quality was assessed using RMHAS and those classified as “Reject” were removed.^9^ Subsequently, only one image with the best quality was kept for each participant. Retinal vascular parameters were obtained using RMHAS^9^, a deep learning algorithm for automated segmentation and quantification of retinal vascular networks. We extracted measurements of the following 5 retinal geometric features: Calibre, Density, Tortuosity, Branching Angle, and Complexity. Multiple measure types, namely the segmentation and quantification methods, were extracted for each retinal geometric feature. (Figure 1) Table S2 provides definitions of those measure types. Building on that, measurements of arterial and venous vessels in the macula and peripheral retina were further quantified.

**Figure 1.**
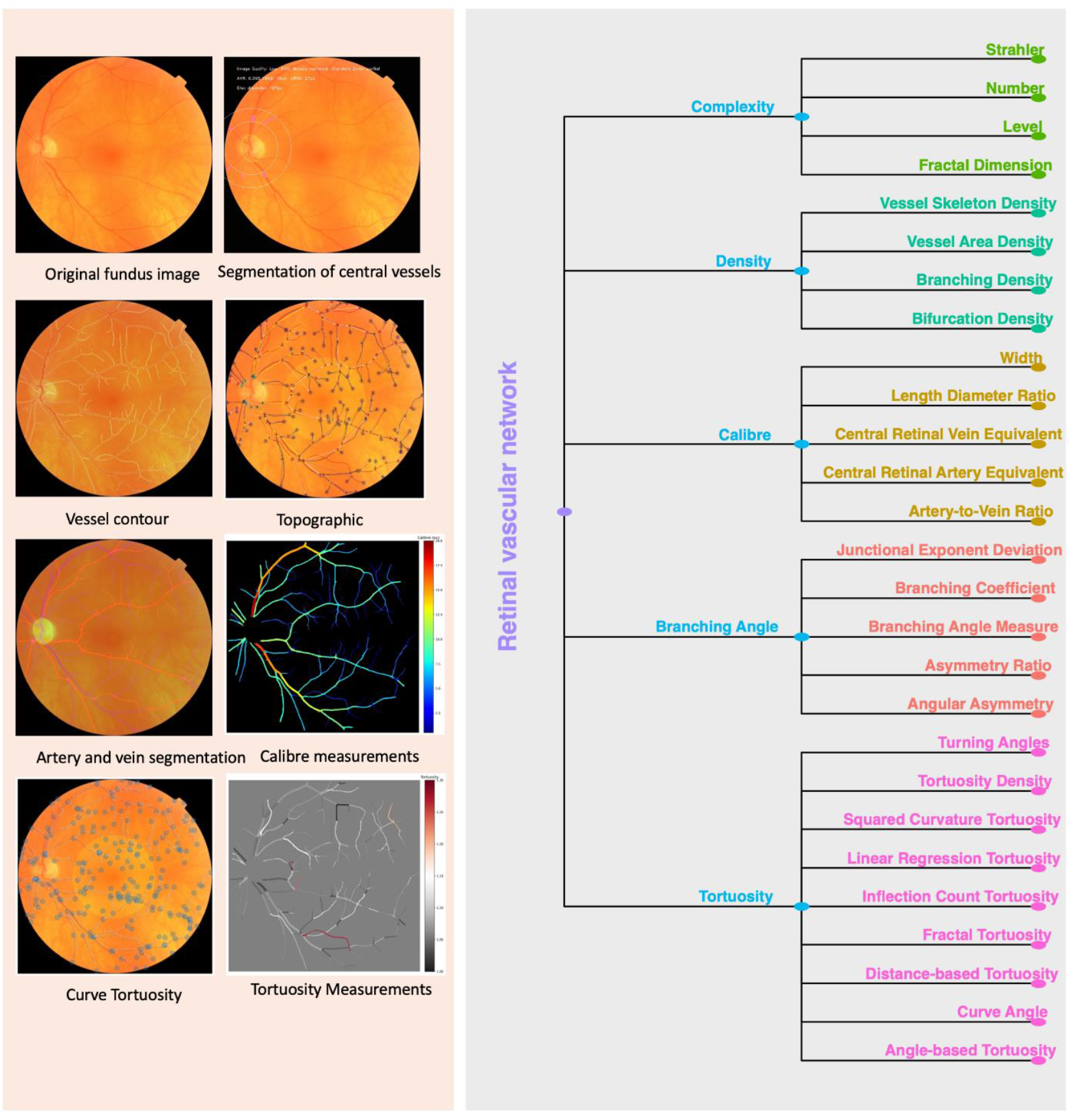
Retinal vascular network analysis. Notes: After retinal vascular network segmentation, we used 27 segmentation and quantification methods to extract measurements of the following 5 retinal vascular geometric features: Calibre, Density, Tortuosity, Branching Angle, and Complexity. Definitions can be found in Supplementary Table 6.

### Statistical analysis

Continuous variables were presented as means (standard deviation) and counts (percentage) were used for categorical variables. For RMHAS-generated retinal vascular measurements, outliers were removed using the Robustbase package, setting the range at 3 to account for skewness^12^. Multivariate imputation by chained equations was used to impute missing values.

The Kolmogorov-Smirnov test was performed to assess the normality of distributions. For comparison between men and women, when data for both groups followed a normal distribution, a T-test was applied, otherwise, the Mann-Whitney U test was used. For comparison between age groups, when measurements were normally distributed, an Analysis of Variance was performed, otherwise, the Kruskal-Wallis H test was used. Violin plots were generated to show the distribution of retinal vascular measurements for men and women separately, further stratified by the decade of age, with intergroup differences labelled.

To explore changes in retinal vessel parameters (calibre, tortuosity, complexity, density, and branching angle) with age, systolic blood pressure (SBP), BMI, glycated haemoglobin (HbA1c), and IOP, we constructed heatmaps depicting these associations. For correlation analyses, the Anderson-Darling test was used for the normality test. If variables were normally distributed, Pearson’s correlation coefficient was used, otherwise, Kendall’s rank correlation coefficient was calculated. To account for multiple comparisons, we applied the Bonferroni correction, adjusting the significance threshold accordingly. For significantly correlated variables after correction, we applied Generalized Additive Models for Location, Scale, and Shape (GAMLSS) separately for men and women to capture both linear and non-linear associations. P-splines were utilized to smooth and enhance the localization of model fit.

We treated a two-sided alpha < 0.05 as significant for all statistical tests, with Bonferroni correction used for multiple comparisons. All analyses were performed using R version 4.2.3 (The R Foundation for Statistical Computing, Vienna, Austria).

## Results

### Characteristics of participants

A total of 10,151 participants were included in the final analysis. Figure 2 details the participants included and excluded at each stage. The mean age was 53.0 years (SD 7.74). The overall sex distribution was 63.8% women and 36.2% men, with similar proportions observed across the age groups. Systolic blood pressure (SBP) measurements showed an increasing trend with age, with means of 121 mmHg, 123 mmHg, and 126 mmHg in the 40-50, 50-60, and >60 years groups, respectively. Table 1 shows sample characteristics by decade age groups and for all participants.

**Figure 2.**
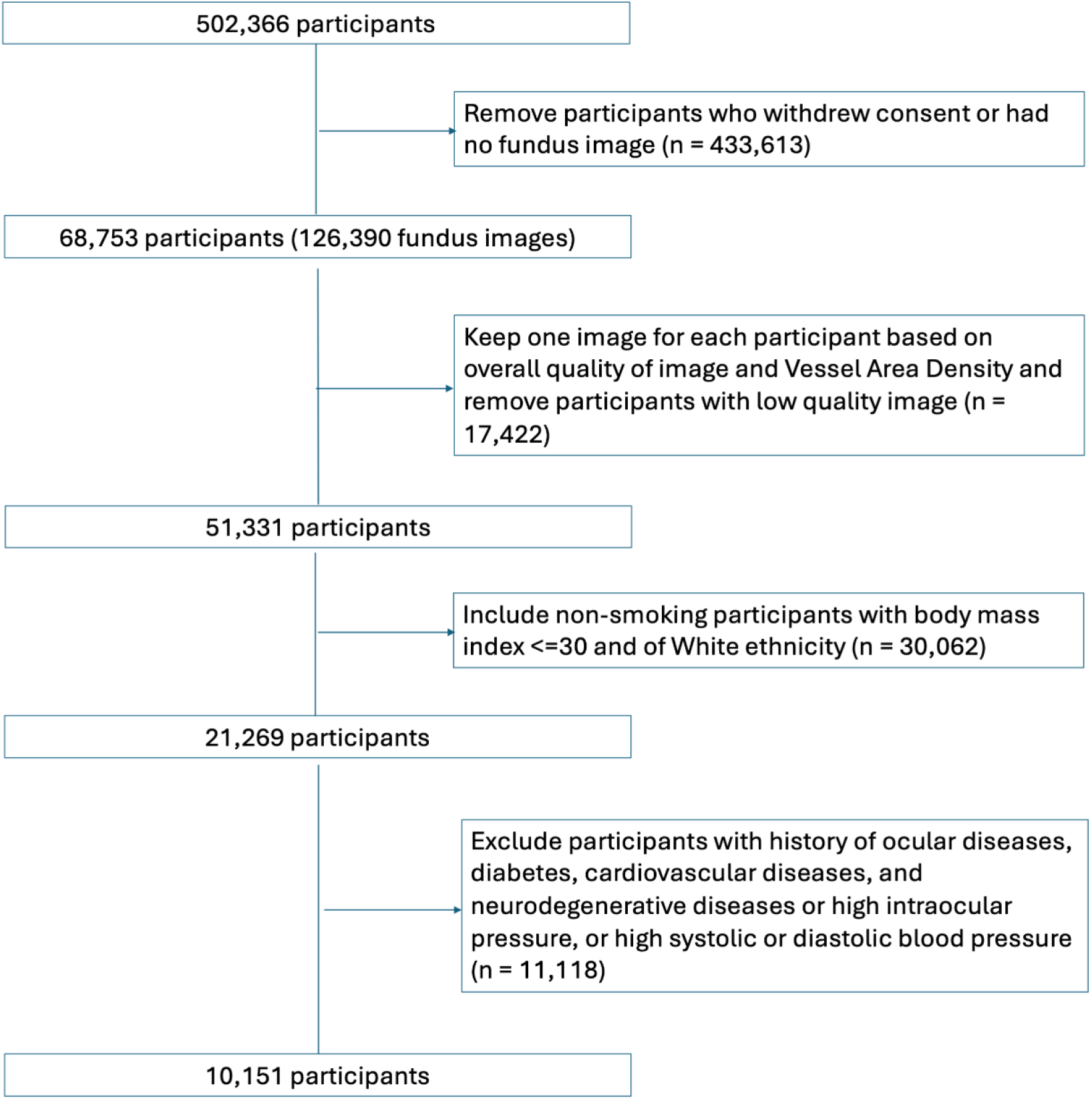
Participants’ selection process. Notes: Out of 502,366 participants, 68,753 participants had fundus images. We first kept one image for each participant based on quality and then further removed those with poor-quality images. Among 51,331 participants with eligible images, we kept only non-smoking participants of White ethnicity and further removed those with conditions that would affect retinal vessels.

**Table 1.** Baseline Characteristics of Participants.

| Characteristics | Overall<br>n = 10151 | 40-50 years<br>n = 3888 | 50-60 years<br>n = 3803 | >60 years<br>n = 2460 |
| --- | --- | --- | --- | --- |
| **Age, years** |  |  |  |  |
|  | 53.0 (7.74) | 45.0 (2.73) | 54.3 (2.86) | 63.5 (2.75) |
| **Sex** |  |  |  |  |
| Female, N (%) | 6476 (63.8%) | 2444 (62.9%) | 2455 (64.6%) | 1577 (64.1%) |
| Male, N (%) | 3675 (36.2%) | 1444 (37.1%) | 1348 (35.4%) | 883 (35.9%) |
| **Body Mass Index (kg/m<sup>2</sup>)** |  |  |  |  |
| Mean (SD) | 24.5 (2.73) | 24.5 (2.77) | 24.5 (2.75) | 24.5 (2.66) |
| Missing, N (%) | 31 (0.31%) | 13 (0.33%) | 10 (0.26%) | 8 (0.33%) |
| **Diastolic Blood Pressure (mmHg)** |  |  |  |  |
| Mean (SD) | 75.2 (7.07) | 75.3 (7.10) | 75.5 (7.10) | 74.6 (6.93) |
| Missing, N (%) | 32 (0.32%) | 11 (0.28%) | 12 (0.32%) | 9 (0.37%) |
| **Systolic Blood Pressure (mmHg)** |  |  |  |  |
| Mean (SD) | 123 (10.1) | 121 (10.2) | 123 (9.91) | 126 (9.30) |
| Missing, N (%) | 32 (0.32%) | 11 (0.28%) | 12 (0.32%) | 9 (0.37%) |
| **High-Density Lipoprotein (mmol/L)** |  |  |  |  |
| Mean (SD) | 1.58 (0.379) | 1.52 (0.348) | 1.62 (0.398) | 1.61 (0.386) |
| Missing, N (%) | 1414 (13.9%) | 511 (13.1%) | 569 (15.0%) | 334 (13.6%) |
| **Low-Density Lipoprotein (mmol/L)** |  |  |  |  |
| Mean (SD) | 3.54 (0.789) | 3.31 (0.729) | 3.63 (0.780) | 3.77 (0.797) |
| Missing, N (%) | 889 (8.75%) | 319 (8.21%) | 351 (9.23%) | 219 (8.90%) |
| **Cholesterol (mmol/L)** |  |  |  |  |
| Mean (SD) | 5.73 (1.03) | 5.38 (0.932) | 5.87 (1.01) | 6.05 (1.04) |
| Missing, N (%) | 871 (8.58%) | 315 (8.10%) | 347 (9.12%) | 209 (8.50%) |
| **Intraocular Pressure (mmHg)** |  |  |  |  |
| Mean (SD) | 14.7 (2.96) | 14.5 (2.96) | 14.7 (3.01) | 15.0 (2.88) |
| Missing, N (%) | 267 (2.63%) | 103 (2.65%) | 101 (2.66%) | 63 (2.56%) |
| **Glycated Haemoglobin (mmol/mol)** |  |  |  |  |
| Mean (SD) | 33.9 (3.59) | 32.7 (3.23) | 34.2 (3.18) | 35.3 (4.14) |
| Missing, N (%) | 1220 (12.0%) | 402 (10.3%) | 493 (13.0%) | 325 (13.2%) |

Notes: SD, standard deviation; N, number. Categorical variables are presented as counts (percentage), while continuous variables are presented as means (SD).

### Distribution of retinal vascular measurements

Supplementary tables 3 and 4 present the mean (SD) and the median (inter-quartile range [IQR]) of retinal vascular measurements in the entire sample and by age group, and supplementary table 5 shows the mean (SD) of retinal vascular measurements for men and women separately with and without age stratification. Figures S1-5 show the median (IQR) of retinal vascular measurements by sex and age group.

#### Calibre

The mean values of Artery-to-Vein Ratio equivalent (AVRe), Central Retinal Artery Equivalent (CRAE), Central Retinal Vein Equivalent (CRVE), and Width of arteries and veins were 0.653 (0.068), 152 (14.9) μm, 233 (21.5) μm, 56.6 (2.96) μm, and 63.3 (3.53) μm respectively. Length-to-Diameter Ratio (LDR) was 12.8 (1.52) for arteries, 12.2 (1.06) for veins, and 13.6 (1.86) for vessels in the macular region. (Supplementary table 3) Values of measurements for women and men and stratified by age groups can be found in Supplementary table 5.

#### Complexity

The mean value of Fractal Dimension (FD), a quantitative, dimensionless measure for the complexity of the vascular network, was 1.77(0.032), with arterial FD being 1.53 (0.039) and venular FD being 1.56 (0.025). The mean Number of Segments was 172 (45.2) and 174 (33.6) for arteries and veins, and 85.5 (22.0) in the macular region. The mean Number of Branching and Bifurcation was 97.9 (28.5) and 51.8 (15.0) for arteries, and 100 (23.1) and 50.1 (11.3) for veins. (Supplementary table 3)

#### Density

The mean Vessel Area Density (VAD) of arteries and veins was 5.75% (0.915%) and 7.22% (0.735%) respectively, and the mean Vessel Skeleton Density (VSD) of arteries and veins was 1.45% (0.243%) and 1.57% (0.178%). The mean Branching and Bifurcation Density was 1.39% (0.172%) and 1.33% (0.204%) respectively for arteries and 1.34% (0.142%) and 1.29% (0.159%) respectively for veins. Chord Length and Arc Length, which inversely reflect the bifurcation density of the vascular network (with longer lengths indicating less bifurcation and therefore lower network density), were 584 (82.3) and 634 (89.1) for arteries, and 596 (63.1) and 646 (68.9) for veins, respectively. (Supplementary table 3)

#### Branching Angle

The mean Angular Asymmetry (AA) was 41.1 (3.736) degree and 39.4 (3.434) degree for arteries and veins respectively, and the Asymmetry Ratio (AR) was 2.13 (0.334) and 2.49 (0.434) respectively. The mean Branching Angle (BA) was 47.1 (1.73) degree and 46.7 (1.36) degree respectively for arteries and veins. The mean Branching Coefficient (BC) was 1.29 (0.092) and 1.22 (0.089) for arteries and veins. (Supplementary table 3)

#### Tortuosity

The mean Tortuosity Density was 0.543 (0.037) and 0.590 (0.032) for arteries and veins respectively. The mean Curve Angle (CA) and Fractal Tortuosity (FT) were 9.71 (1.49) degree and 0.84 (0.009) respectively for arteries and 10.1 (0.992) degree and 0.838 (0.008) respectively for veins. In addition, Inflection Count Tortuosity and Distance-based Tortuosity were 0.226 (0.017) and 1.09 (0.009) for arteries and 0.245 (0.016) and 1.08 (0.005) for veins. (Supplementary table 3)

### Associations of retinal vascular measurements with health metrics

Figure 3 presents correlations between retinal vascular measurements and health metrics, including age, SBP, IOP, HbA1c, and BMI with P value adjusted for Bonferroni correction. Coefficient and adjusted P values can be found in Table S6. Across all health metrics included in the analysis, age, and SBP were significantly associated with the most retinal vascular measurements with the highest coefficients. Figure 4 shows the associations of selected retinal vascular measurements with age and SBP assessed with GAMLSS with P-splines, accounting for the effect of remaining health metrics.

**Figure 3.**
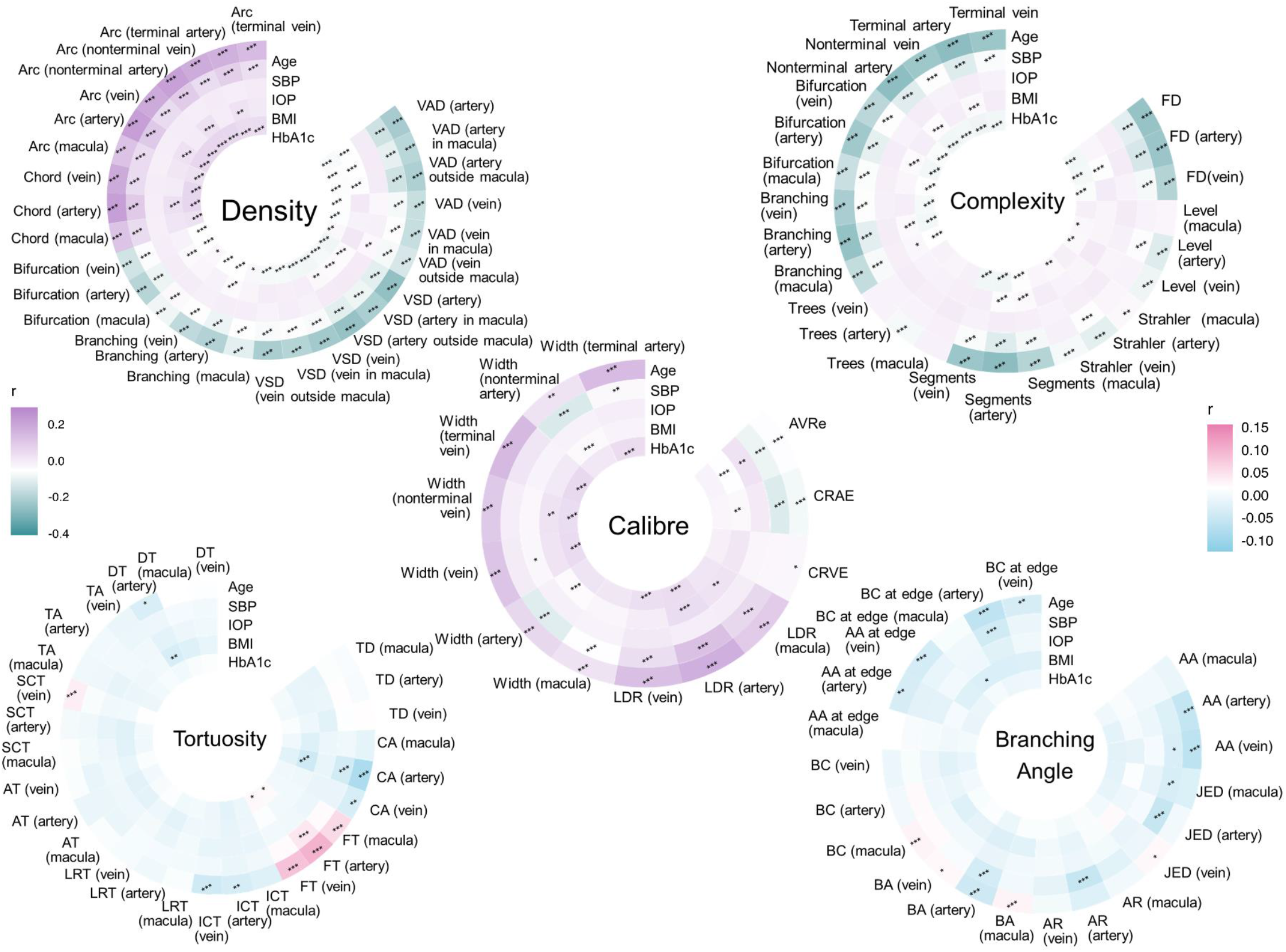
Correlations between Retinal Vascular Measurements and Age, SBP, IOP, HbA1c, and BMI. Notes: BMI: Body Mass Index; SBP: Systolic Blood Pressure; IOP: Intraocular Pressure; HbA1c: Glycated Hemoglobin; VAD: Vessel Area Density, VSD: Vessel Skeleton Density. The P value was adjusted with Bonferroni correction. ***: P <0.001, **: P <0.01, *: P <0.05.

**Figure 4.**
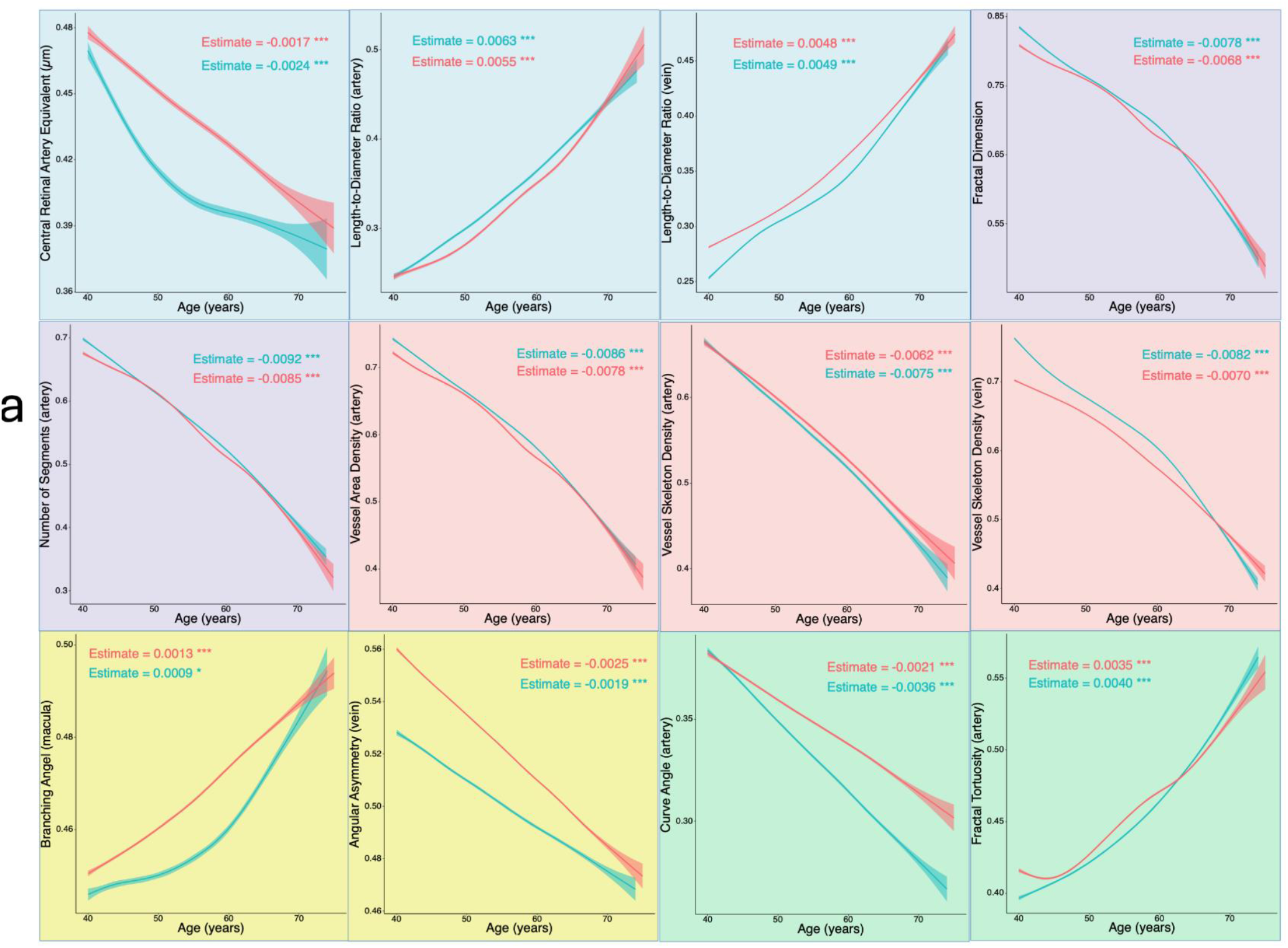
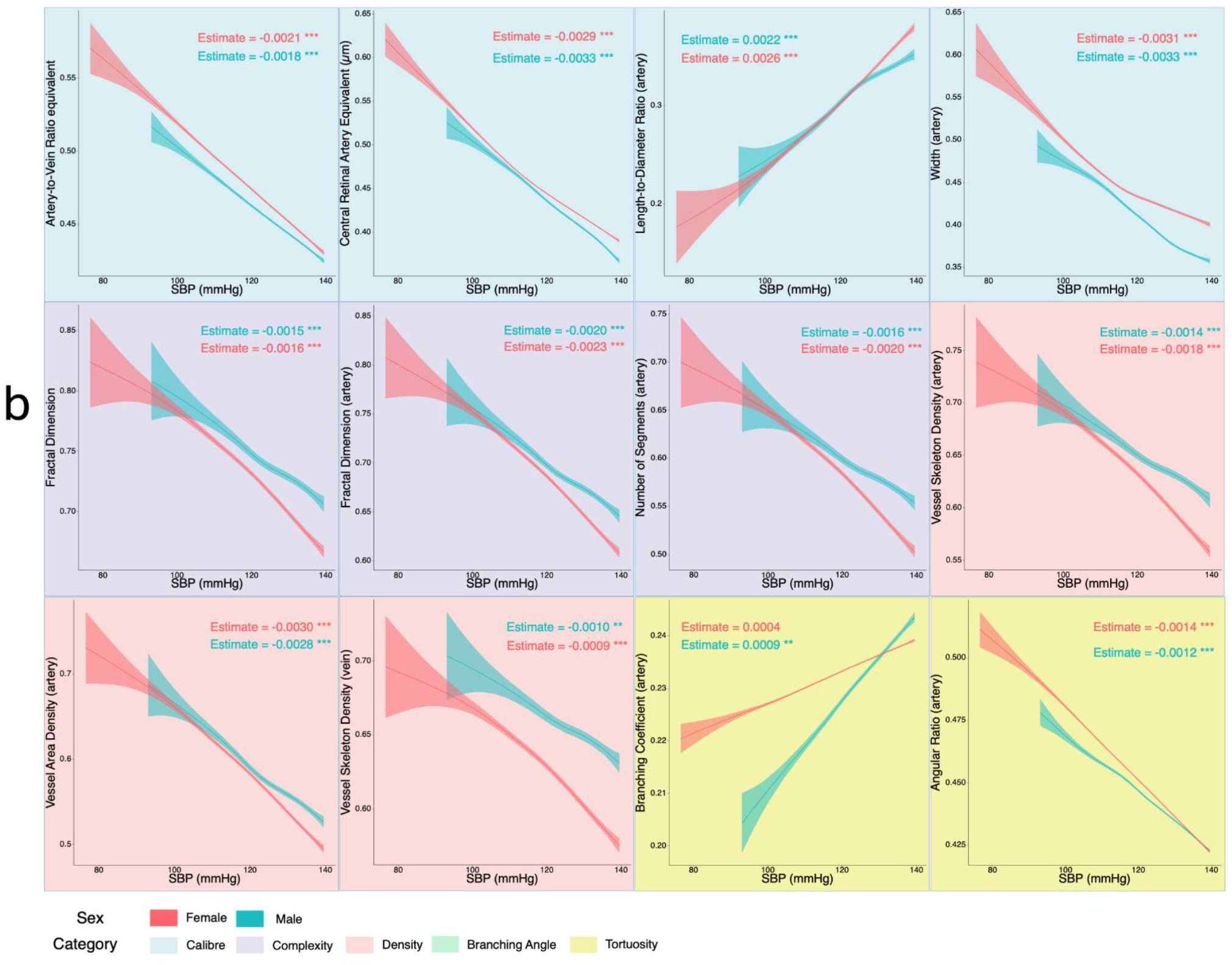
Associations of Selected Retinal Vascular Measurements with Age and SBP for men (n= 3675) and women (n= 6476) Notes: Figure 4a: Associations of Selected Retinal Vascular Measurements with Age. Figure 4b: Associations of Selected Retinal Vascular Measurements with SBP. Generalized Additive Models for location, scale, and shape (GAMLSS) were fitted for men and women separately with P-splines. SBP: Systolic Blood Pressure. Y axis shows the partial effect of age and SBP on retinal vascular measurements accounting for the impact of other health metrics. The retinal vascular measurements were rescaled to the 0-1 range to improve the comparability (the association assessed using original values can be found in Figure S6).

#### Calibre

Age was associated with all Calibre measurements. SBP showed association with all arterial measurements as well as measurements in the macular region. Among venular measurements, SBP showed with venular LDR only. BMI also showed associations with all arterial measurements, as well as AVRe and LDR in the macular region. IOP showed association only with the width of veins and AVRe, and HbA1c was associated with both arterial and venular LDR, as well as the Width of veins.

#### Complexity

IOP was not associated with any Complexity measurements, while age was associated with all except the Number of venular Trees and the Number of Trees in the macular region. SBP and HbA1c were associated with all measurements of Number of Bifurcation, Number of Branching, Number of Segments, and FD. BMI was associated with arterial FD, Number of arterial Branching, Bifurcation, Segments, and Number of Branching and Segments in the macular region.

#### Density

All Density measurements showed a strong association with age, SBP, and HbA1c. IOP was only associated with VAD of veins outside the macular region. BMI was associated with all arterial Density measurements and none of the venular measurements or measurements in the macular region.

#### Branching Angle

Age was associated with all venular measurements, except for venular BA at the edge. In addition, age was arterial AA, BA, as well as AA and BA at the edge. For measurements in the macular region, age showed an association with only BA. SBP was associated with arterial AA, Junctional Exponent Deviation (JED), BA, and BA at the edge, as well as JED in the macular region. IOP and HbA1c did not show association with any of the Branching Angle measurements, and BMI was only associated with BA at the edge in the macular region.

#### Tortuosity

Age was associated with arterial and venular Inflection Count Tortuosity, FT, and CA, as well as venular Squared Curvature Tortuosity and arterial Distance-based Tortuosity. SBP was associated with arterial CA and FT. IOP was not associated with any of the Tortuosity measurements. BMI with none but arterial CA and Distance-based Tortuosity, and HbA1c with none but arterial and venular FT.

## Discussion

The current study selected 10,000 healthy participants from a large database to establish normative ranges for a wide variety of retinal vascular parameters. Ours is the first study to present a comprehensive suite of age- and sex-stratified retinal vascular measurements, categorized into five distinct groups and providing separate parameters for veins and arteries and locations within and outside the macular region. This analysis provides a foundation for future research exploring the clinical significance of variations in retinal vasculature parameters in disease diagnosis and risk assessment. As more retinal vascular measurements are increasingly identified as biomarkers for systemic conditions, the normative ranges established in our study become more useful.

In our study, the mean values of CRAE, CRVE, and AVRe, 152 (14.9) μm, 233 (21.5) μm, and 0.653 (0.068) respectively, were similar to those reported in the Australian population, (152 [14.0] μm, 221 [19.0] μm, and 0.69 [0.06])^13^. However, measurements in prior studies have ranged from 163 μm to 192 μm for CRAE and 210 μm to 250 μm for CRVE^6,14,15^. While studies differed in age and inclusion criteria, the most important reason for discrepancies may lie in varying measurement approaches. The arterial and venular FD values in the current report were 1.53 (0.04) and 1.56 (0.03) in women and 1.53 (0.04) and 1.57 (0.03) in men, which were somewhat higher than those reported in another recent study^16^. As prior reports show FD is negatively associated with cardiovascular disease and mortality risk^5,17^, a higher FD in our study may reflect our strict inclusion criteria, excluding all participants with cardiovascular disease risk factors.

Among all the health metrics we examined, age had the strongest association with retinal vessel measurements. CRAE, CRVE, and AVRe all decreased with age, consistent with data reported for another healthy cohort^18^. In contrast, LDR of all veins and arteries, as well as those in the macular region, were all positively associated with age, consistent with a previous study reporting a positive and independent association between higher arteriolar LDR and older age^19^. In addition, age showed a strong negative association with all Density measurements and most Complexity measurements, consistent with previous studies reporting sparser vascular networks and less complex microvasculature with increasing age^20-22^. Specifically, in our study, the correlation matrix revealed correlations of -0.24 and -0.21 for age with arterial FD and VAD, showing consistent trends with findings from a previous study (−0.17 and -0.25)^16^.

SBP was associated with reduced CRAE and CRVE, with a greater reduction in CRAE and, consequently, a reduced AVRe. The same trend was found previously in children as well, indicating such trends start early in life^23,24^. As with age, SBP was also associated with all LDR measurements. A previous study also revealed a positive association of SBP with arteriolar LDR^19^, while they reported no association for venular LDR. SBP exhibited a negative association with both Complexity and Density measurements, for both arteries and veins, within and outside the macular region. A similar inverse association has also been reported^20,22^, including in a multiethnic population study^25^. As shown in Figure 4, the associations of retinal vascular measurements with age and SBP were generally linear. Since we adopted strict inclusion criteria to include only participants without cardiovascular disease risk factors, caution is warranted when interpreting these results.

HbA1c was also associated with retinal vascular measurements. However, the association was mainly found for Complexity and Density measurements, both negative. An increase in HbA1c could affect endothelial function and vascular bifurcation^26-28^, thereby leading to lower Complexity and Density of the retinal network. The association with BMI was mainly found for Calibre measurements, which were positively associated with CRVE and width of veins and negatively associated with CRAE, AVRe, and width of arteries, consistent with previous studies^23,29^. We also showed that, after excluding those with ocular conditions that could affect the retinal vascular network, including high IOP, glaucoma, and retinal diseases, in participants with relatively healthy retinal vascular networks, no association with IOP was found for most retinal geometric measurements. We found IOP was negatively associated with both CRVE and the width of veins, which may also explain its positive association with AVRe and venular VAD (across the fundus and outside the macular region). Most studies in glaucoma patients have also identified a reduction in venular calibre^30^.

Our study has several strengths. First, we reported normative data stratified by sex and age. This provides a valuable reference database for age- and sex-specific studies exploring retinal vascular damage. Second, separate measurements of arteries and veins allow researchers to precisely investigate diseases that primarily affect one or the other. Third, we reported over a hundred measurements. This provides a large set of candidates for potential biomarker identification and assessment of retinal vascular alteration. In addition, we found the changes in arterial measurements with age and SBP were more prevalent and significant than those of venular measurements. This suggests that arterial measurements may serve as more sensitive biomarkers for age- and/or blood pressure-related systemic conditions and are of value for further exploration.

We also recognize limitations in our research. Firstly, our analysis was based on UK Biobank participants of white ethnicity. Consequently, there is a need for future studies to establish normative ranges within multiethnic populations. Additionally, due to the strict inclusion criteria necessary for reporting normative ranges, we found that associations with other health metrics were generally linear. Therefore, caution is advised when interpreting these results beyond the normal range. Lastly, our study did not compare vessel parameters with those generated by alternative software, which may limit the generalizability of our findings to analyses conducted with different analytic algorithms. This limitation arises in part from the inability of other software to produce as extensive a set of parameters. Nonetheless, the algorithm we used is validated ^4,5,31,32^, and the associations we identified are consistent with those reported in previous studies. Future research is required to provide a more comprehensive perspective on this issue.

## Conclusion

With AI-facilitating segmentation and quantification of retinal vascular network, increasing evidence validates the associations of retinal vascular biomarkers with systemic conditions and mortality risk, underscoring their potential as a low-cost, non-invasive, easy-to-use, and widely available screening tool. By reporting population normative data for a comprehensive set of retinal vascular measurements, our study enables a better understanding of the severity of alterations in the retinal vascular network. This offers a quantifiable approach to the use of retinal vascular biomarkers as an assessment and diagnostic tool.

## Supporting information

STROBE_checklist_cross-sectional

## Data Availability Statement

This study used data from a public, open access repository, the UK Biobank study (https://www.ukbiobank.ac.uk/).

## Supplementary Material

**Supplementary Figure 1**. Distribution of Calibre Measurements

**Supplementary Figure 2**. Distribution of Complexity Measurements

**Supplementary Figure 3**. Distribution of Density Measurements

**Supplementary Figure 4**. Distribution of Branching Angle Measurements

**Supplementary Figure 5**. Distribution of Tortuosity Measurements

**Supplementary Figure 6**. Associations of Selected Retinal Vascular Measurements with Age and SBP

**Supplementary Figure 7**. Correlations among retinal vascular measurements

**Supplementary Figure 8**. Deviation of retinal vascular measurements in the excluded participants

**Supplementary Table 1**. Mean (standard deviation) of retinal vascular measurements by age groups

**Supplementary Table 2**. Median (1st quartile to 3rd quartile) of retinal vascular measurements by age groups

**Supplementary Table 3**. Normative data of retinal vascular parameters by sex and age groups

**Supplementary Table 4**. Correlation between retinal vascular parameters and health metrics

**Supplementary Table 5**. Definitions of variables used in the current study

**Supplementary Table 6**. Definitions of measure types

## Notes

**Funding:** This work was supported by the Global STEM Professorship Scheme (P0046113). The Centre for Eye Research Australia receives Operational Infrastructure Support from the Victorian State Government. M.Y. is supported by the Melbourne Research Scholarship established by the University of Melbourne. The funding source had no role in the design and conduct of the study; collection, management, analysis, and interpretation of the data; preparation, review, or approval of the manuscript; and decision to submit the manuscript for publication.

### Competing Interest Statement

The authors have declared no competing interest.

### Funding Statement

This work was supported by the Global STEM Professorship Scheme (P0046113). The Centre for Eye Research Australia receives Operational Infrastructure Support from the Victorian State Government. M.Y. is supported by the Melbourne Research Scholarship established by the University of Melbourne. The funding source had no role in the design and conduct of the study; collection, management, analysis, and interpretation of the data; preparation, review, or approval of the manuscript; and decision to submit the manuscript for publication.

### Author Declarations

The current study utilized data from the UK Biobank, which obtained ethical approval from the North West Multi-Centre Research Ethics Committee (reference number 06/MRE08/65).

