## Supplementary figures 1-8, Supplementary tables 1-6 for "Population-based Normative Reference for Retinal Microvascular Atlas"

### Supplementary Figure 1. Distribution of Calibre Measurements

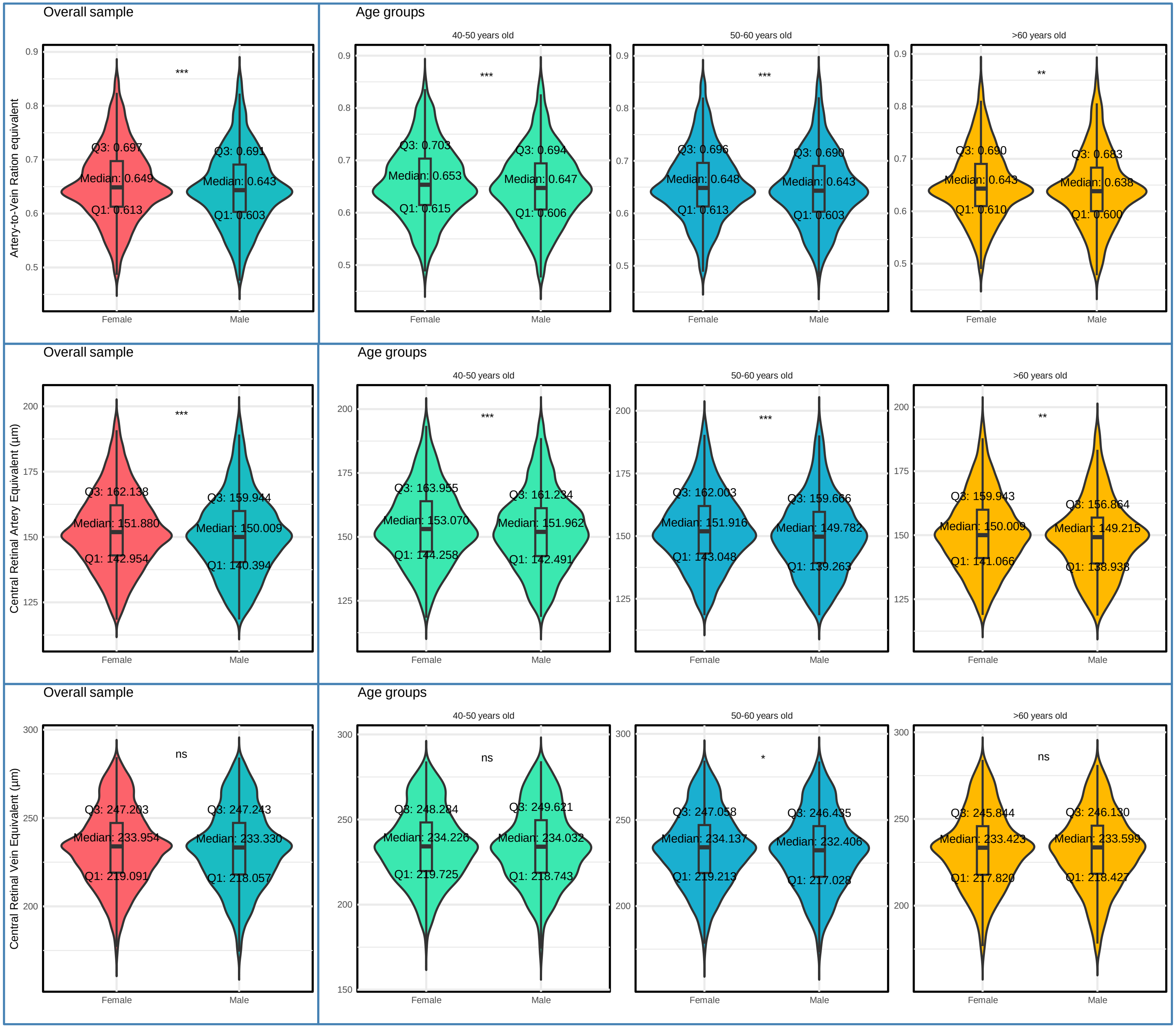

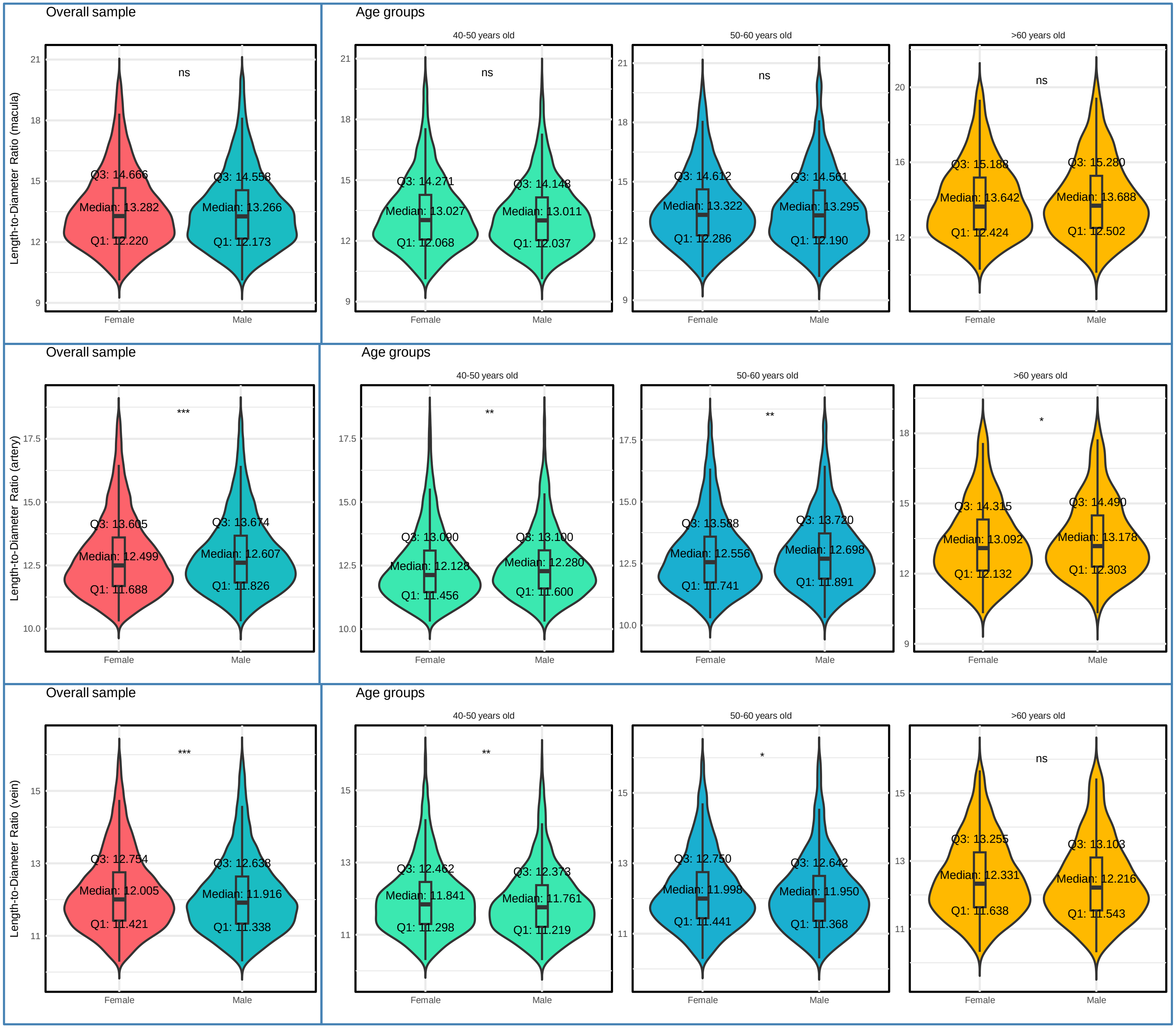

Overall sample
Age groups
40-50 years old
50-60 years old
>60 years old
Length-to-Diameter Ratio (macula)
ns
Q3: 14.666
Median: 13.282
Q1: 12.220
Q3: 14.558
Median: 13.266
Q1: 12.173
Q3: 14.271
Median: 13.027
Q1: 12.068
Q3: 14.148
Median: 13.011
Q1: 12.037
Q3: 14.612
Median: 13.322
Q1: 12.286
Q3: 14.561
Median: 13.295
Q1: 12.190
Q3: 15.188
Median: 13.642
Q1: 12.424
Q3: 15.280
Median: 13.688
Q1: 12.502
Length-to-Diameter Ratio (artery)
***
**
*
Q3: 13.605
Median: 12.499
Q1: 11.688
Q3: 13.674
Median: 12.607
Q1: 11.826
Q3: 13.090
Median: 12.128
Q1: 11.456
Q3: 13.100
Median: 12.280
Q1: 11.600
Q3: 13.588
Median: 12.556
Q1: 11.741
Q3: 13.720
Median: 12.698
Q1: 11.891
Q3: 14.315
Median: 13.092
Q1: 12.132
Q3: 14.490
Median: 13.178
Q1: 12.303
Length-to-Diameter Ratio (vein)
Q3: 12.754
Median: 12.005
Q1: 11.421
Q3: 12.638
Median: 11.916
Q1: 11.338
Q3: 12.462
Median: 11.841
Q1: 11.298
Q3: 12.373
Median: 11.761
Q1: 11.219
Q3: 12.750
Median: 11.998
Q1: 11.441
Q3: 12.642
Median: 11.950
Q1: 11.368
Q3: 13.255
Median: 12.331
Q1: 11.638
Q3: 13.103
Median: 12.216
Q1: 11.543
Female
Male

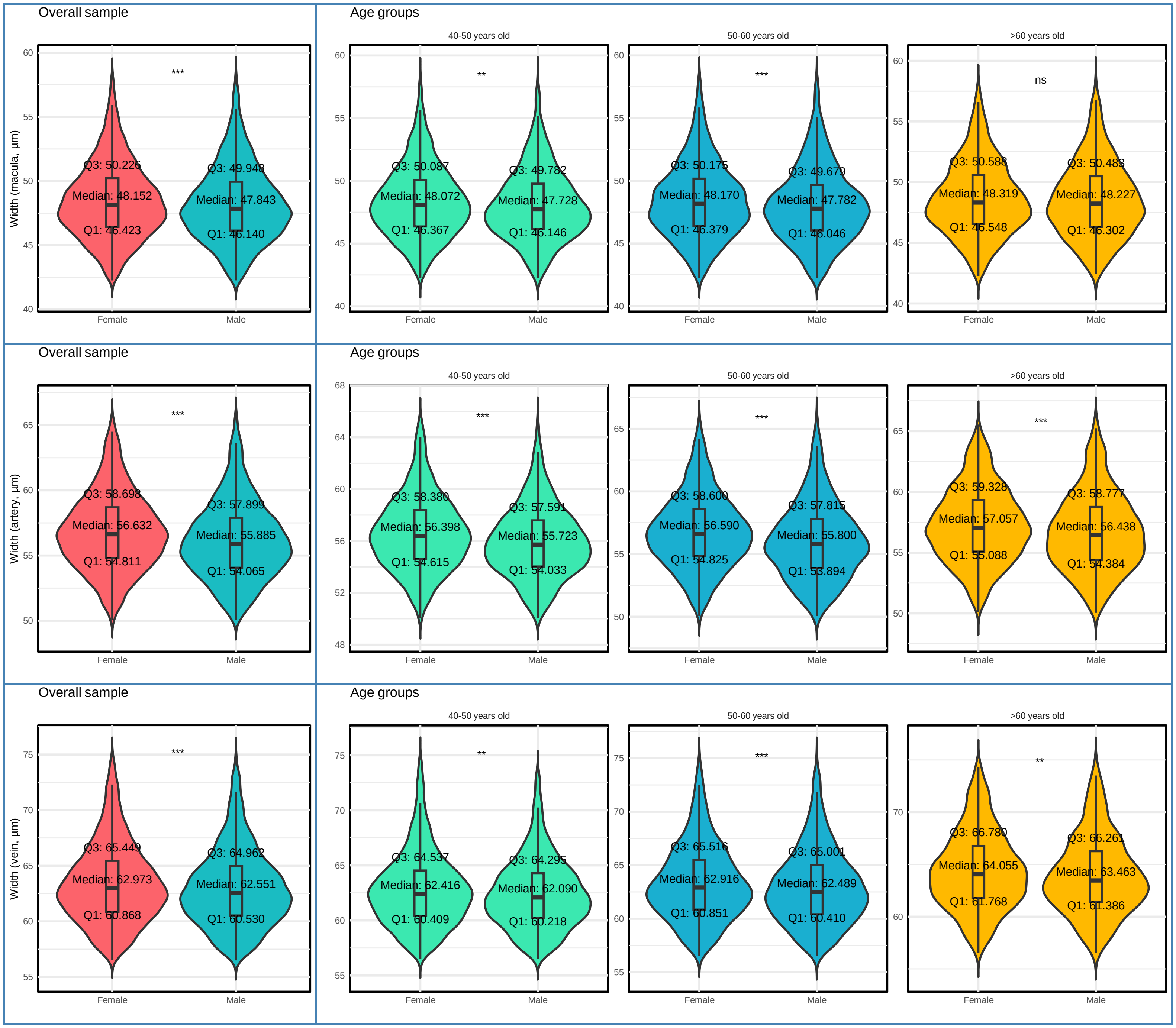
Overall sample
Width (macula, μm)
***
Q3: 50.226
Median: 48.152
Q1: 46.423
Q3: 49.948
Median: 47.843
Q1: 46.140
Female
Male
Age groups
40-50 years old
**
Q3: 50.087
Median: 48.072
Q1: 46.367
Q3: 49.782
Median: 47.728
Q1: 46.146
50-60 years old
***
Q3: 50.175
Median: 48.170
Q1: 46.379
Q3: 49.679
Median: 47.782
Q1: 46.046
>60 years old
ns
Q3: 50.588
Median: 48.319
Q1: 46.548
Q3: 50.483
Median: 48.227
Q1: 46.302
Overall sample
Width (artery, μm)
***
Q3: 58.698
Median: 56.632
Q1: 54.811
Q3: 57.899
Median: 55.885
Q1: 54.065
Age groups
40-50 years old
***
Q3: 58.380
Median: 56.398
Q1: 54.615
Q3: 57.591
Median: 55.723
Q1: 54.033
50-60 years old
***
Q3: 58.600
Median: 56.590
Q1: 54.825
Q3: 57.815
Median: 55.800
Q1: 53.894
>60 years old
***
Q3: 59.328
Median: 57.057
Q1: 55.088
Q3: 58.777
Median: 56.438
Q1: 54.384
Overall sample
Width (vein, μm)
***
Q3: 65.449
Median: 62.973
Q1: 60.868
Q3: 64.962
Median: 62.551
Q1: 60.530
Age groups
40-50 years old
**
Q3: 64.537
Median: 62.416
Q1: 60.409
Q3: 64.295
Median: 62.090
Q1: 60.218
50-60 years old
***
Q3: 65.516
Median: 62.916
Q1: 60.851
Q3: 65.001
Median: 62.489
Q1: 60.410
>60 years old
**
Q3: 66.780
Median: 64.055
Q1: 61.768
Q3: 66.261
Median: 63.463
Q1: 61.386

### Supplementary Figure 2. Distribution of Complexity Measurements

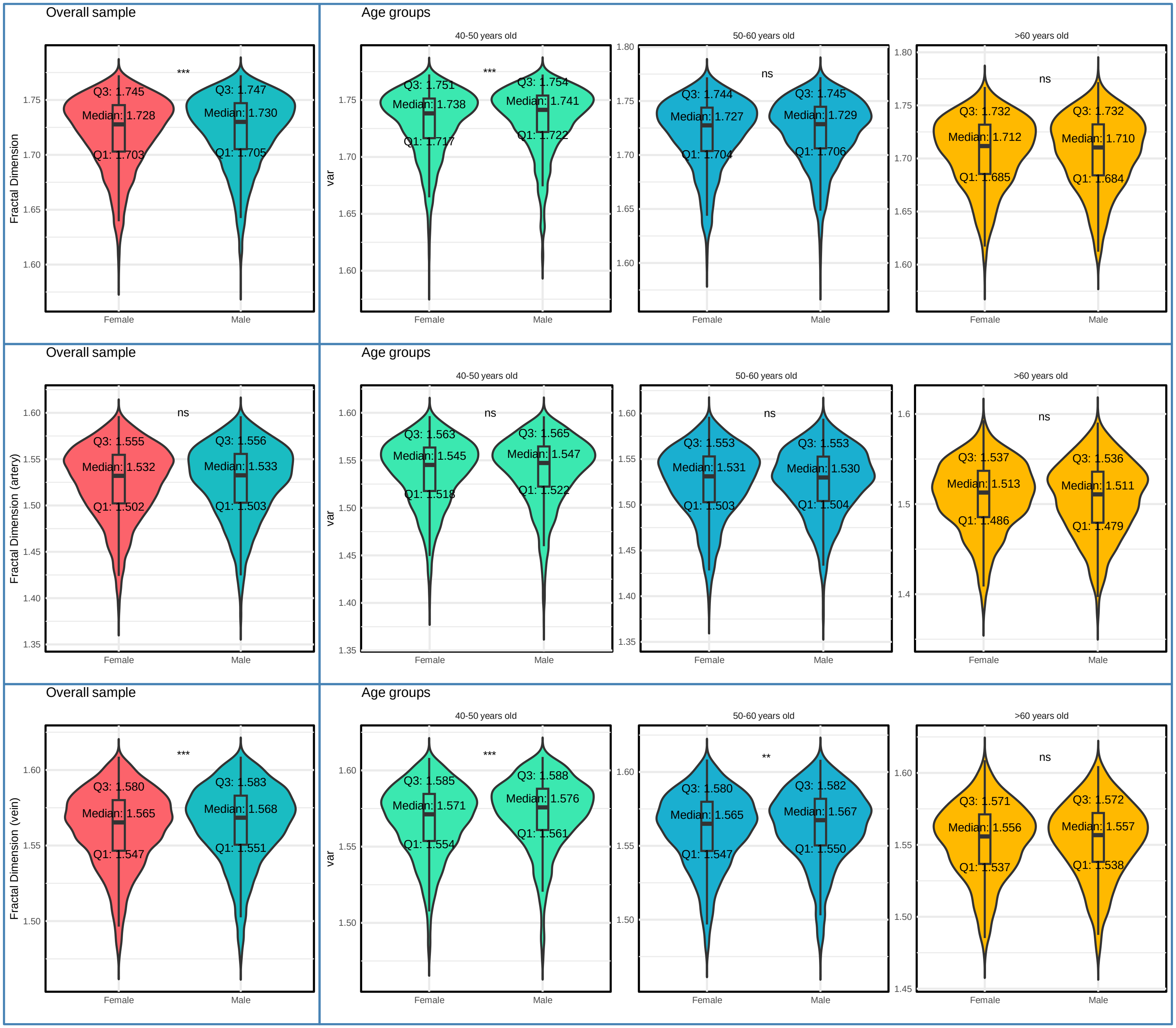

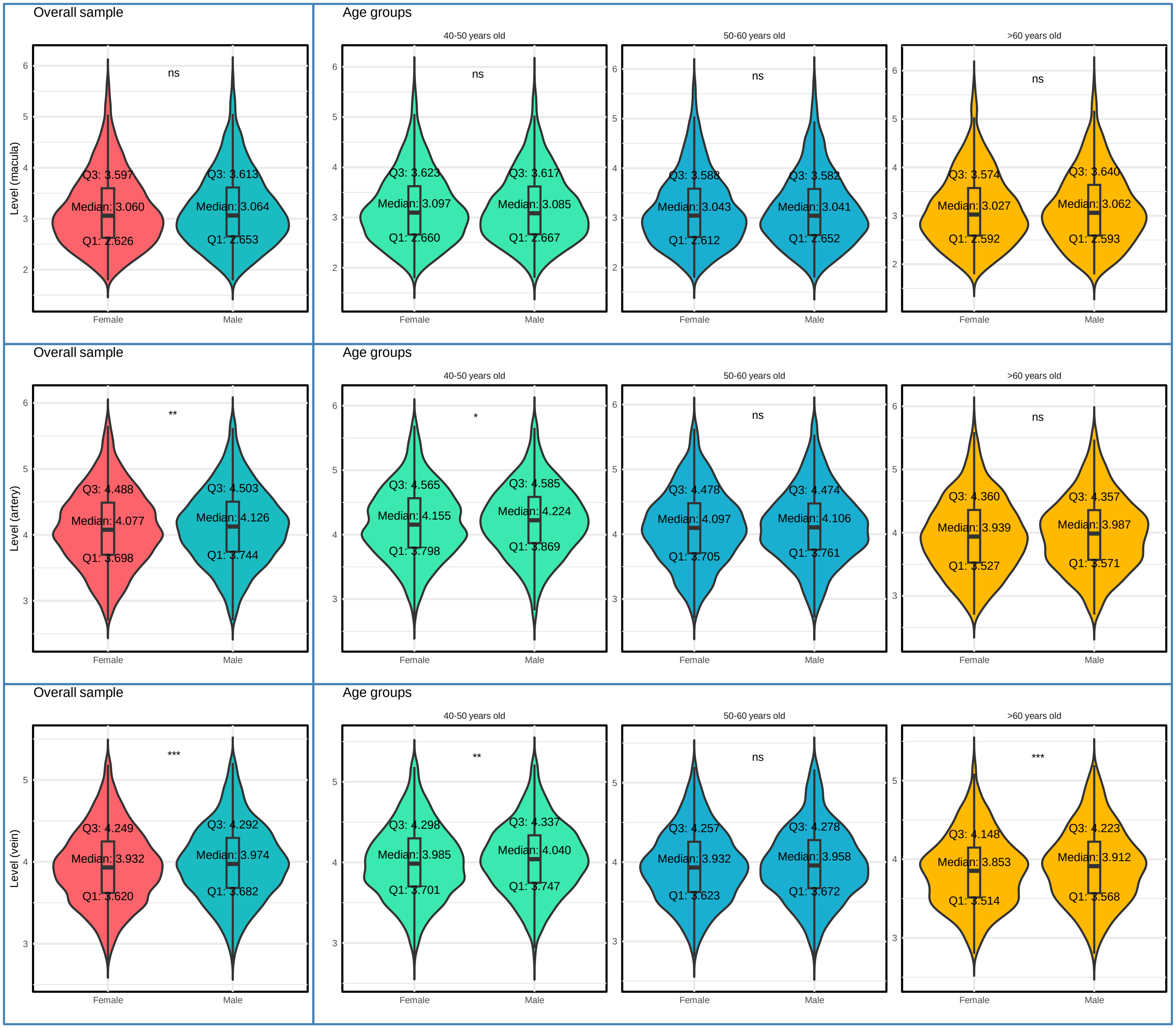
Overall sample
Age groups
40-50 years old
50-60 years old
>60 years old
Level (macula)
ns
Q3: 3.597
Median: 3.060
Q1: 2.626
Q3: 3.613
Median: 3.064
Q1: 2.653
Q3: 3.623
Median: 3.097
Q1: 2.660
Q3: 3.617
Median: 3.085
Q1: 2.667
Q3: 3.588
Median: 3.043
Q1: 2.612
Q3: 3.582
Median: 3.041
Q1: 2.652
Q3: 3.574
Median: 3.027
Q1: 2.592
Q3: 3.640
Median: 3.062
Q1: 2.593
Female
Male
Level (artery)
**
*
Q3: 4.488
Median: 4.077
Q1: 3.698
Q3: 4.503
Median: 4.126
Q1: 3.744
Q3: 4.565
Median: 4.155
Q1: 3.798
Q3: 4.585
Median: 4.224
Q1: 3.869
Q3: 4.478
Median: 4.097
Q1: 3.705
Q3: 4.474
Median: 4.106
Q1: 3.761
Q3: 4.360
Median: 3.939
Q1: 3.527
Q3: 4.357
Median: 3.987
Q1: 3.571
Level (vein)
***
Q3: 4.249
Median: 3.932
Q1: 3.620
Q3: 4.292
Median: 3.974
Q1: 3.682
Q3: 4.298
Median: 3.985
Q1: 3.701
Q3: 4.337
Median: 4.040
Q1: 3.747
Q3: 4.257
Median: 3.932
Q1: 3.623
Q3: 4.278
Median: 3.958
Q1: 3.672
Q3: 4.148
Median: 3.853
Q1: 3.514
Q3: 4.223
Median: 3.912
Q1: 3.568

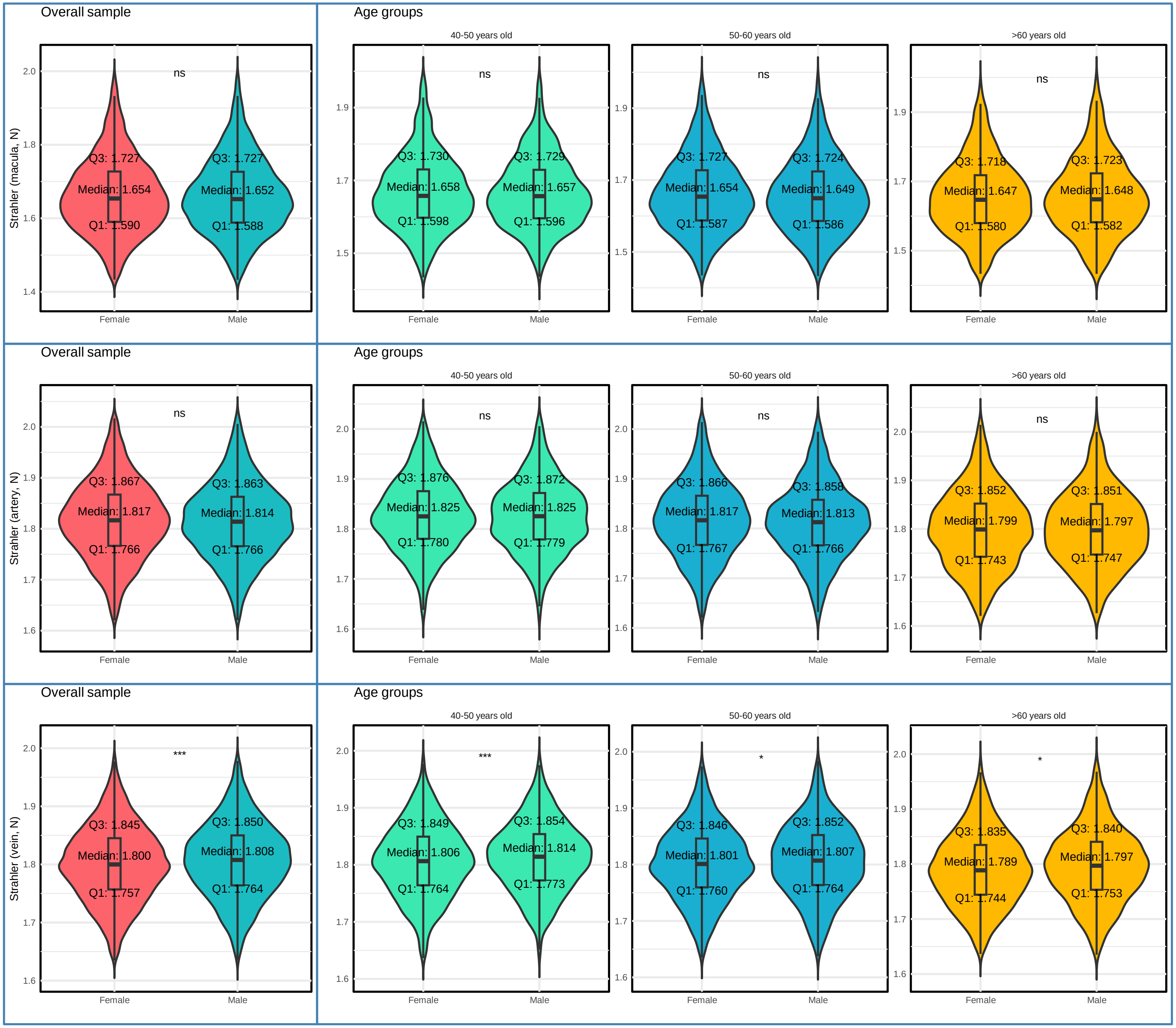
Overall sample
Age groups
40-50 years old
50-60 years old
>60 years old
Strahler (macula, N)
ns
Q3: 1.727
Median: 1.654
Q1: 1.590
Q3: 1.727
Median: 1.652
Q1: 1.588
Q3: 1.730
Median: 1.658
Q1: 1.598
Q3: 1.729
Median: 1.657
Q1: 1.596
Q3: 1.727
Median: 1.654
Q1: 1.587
Q3: 1.724
Median: 1.649
Q1: 1.586
Q3: 1.718
Median: 1.647
Q1: 1.580
Q3: 1.723
Median: 1.648
Q1: 1.582
Strahler (artery, N)
Q3: 1.867
Median: 1.817
Q1: 1.766
Q3: 1.863
Median: 1.814
Q1: 1.766
Q3: 1.876
Median: 1.825
Q1: 1.780
Q3: 1.872
Median: 1.825
Q1: 1.779
Q3: 1.866
Median: 1.817
Q1: 1.767
Q3: 1.858
Median: 1.813
Q1: 1.766
Q3: 1.852
Median: 1.799
Q1: 1.743
Q3: 1.851
Median: 1.797
Q1: 1.747
Strahler (vein, N)
***
*
Q3: 1.845
Median: 1.800
Q1: 1.757
Q3: 1.850
Median: 1.808
Q1: 1.764
Q3: 1.849
Median: 1.806
Q1: 1.764
Q3: 1.854
Median: 1.814
Q1: 1.773
Q3: 1.846
Median: 1.801
Q1: 1.760
Q3: 1.852
Median: 1.807
Q1: 1.764
Q3: 1.835
Median: 1.789
Q1: 1.744
Q3: 1.840
Median: 1.797
Q1: 1.753
Female
Male

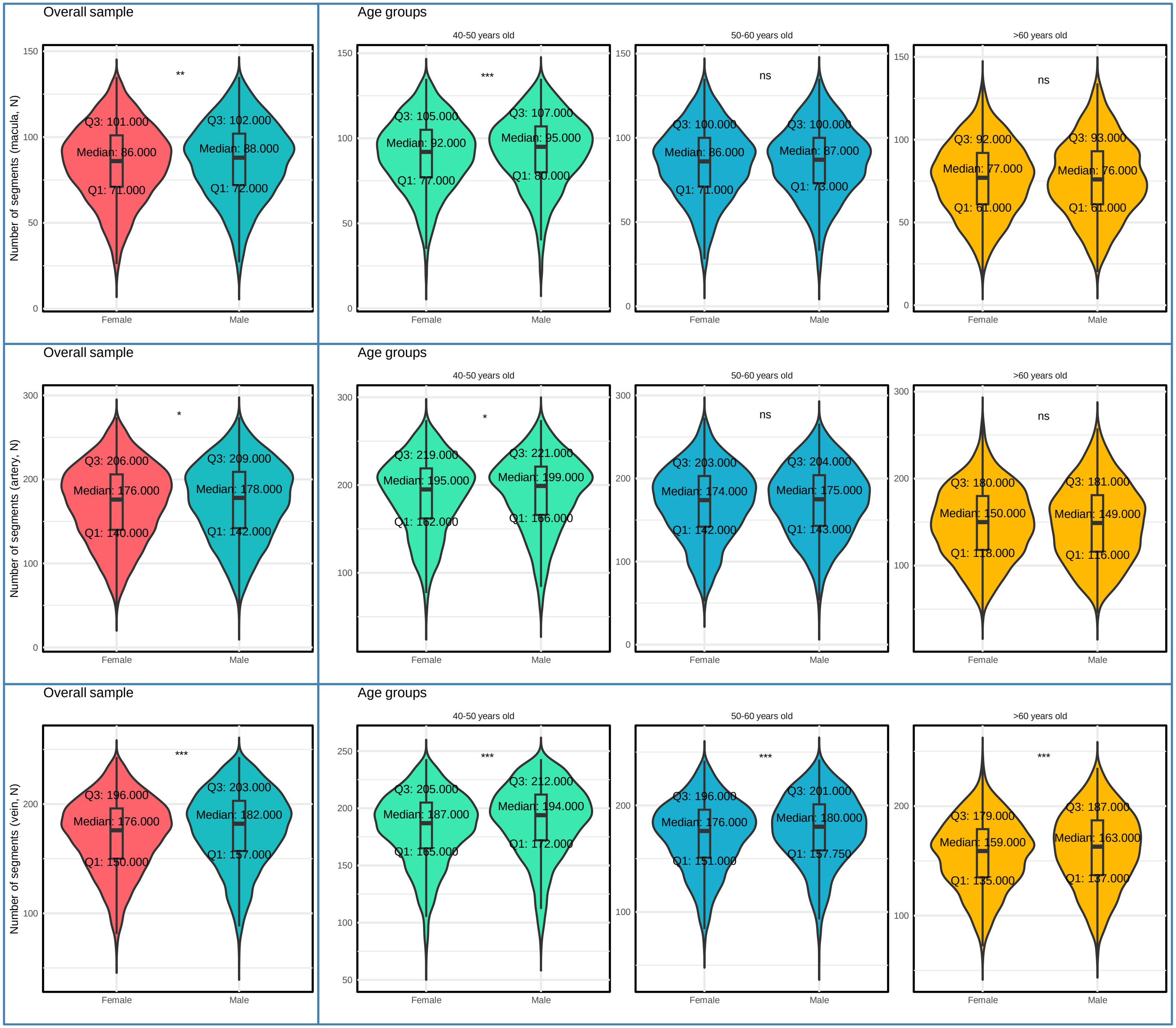
Overall sample
Age groups
40-50 years old
50-60 years old
>60 years old
Number of segments (macula, N)
Female
Male
Q3: 101.000
Median: 86.000
Q1: 71.000
Q3: 102.000
Median: 88.000
Q1: 72.000
**
Q3: 105.000
Median: 92.000
Q1: 77.000
Q3: 107.000
Median: 95.000
Q1: 80.000
***
Q3: 100.000
Median: 86.000
Q1: 71.000
Q3: 100.000
Median: 87.000
Q1: 73.000
ns
Q3: 92.000
Median: 77.000
Q1: 61.000
Q3: 93.000
Median: 76.000
Q1: 61.000
ns
Overall sample
Age groups
40-50 years old
50-60 years old
>60 years old
Number of segments (artery, N)
Female
Male
Q3: 206.000
Median: 176.000
Q1: 140.000
Q3: 209.000
Median: 178.000
Q1: 142.000
*
Q3: 219.000
Median: 195.000
Q1: 162.000
Q3: 221.000
Median: 199.000
Q1: 166.000
*
Q3: 203.000
Median: 174.000
Q1: 142.000
Q3: 204.000
Median: 175.000
Q1: 143.000
ns
Q3: 180.000
Median: 150.000
Q1: 118.000
Q3: 181.000
Median: 149.000
Q1: 116.000
ns
Overall sample
Age groups
40-50 years old
50-60 years old
>60 years old
Number of segments (vein, N)
Female
Male
Q3: 196.000
Median: 176.000
Q1: 150.000
Q3: 203.000
Median: 182.000
Q1: 157.000
***
Q3: 205.000
Median: 187.000
Q1: 165.000
Q3: 212.000
Median: 194.000
Q1: 172.000
***
Q3: 196.000
Median: 176.000
Q1: 151.000
Q3: 201.000
Median: 180.000
Q1: 157.750
***
Q3: 179.000
Median: 159.000
Q1: 135.000
Q3: 187.000
Median: 163.000
Q1: 137.000
***

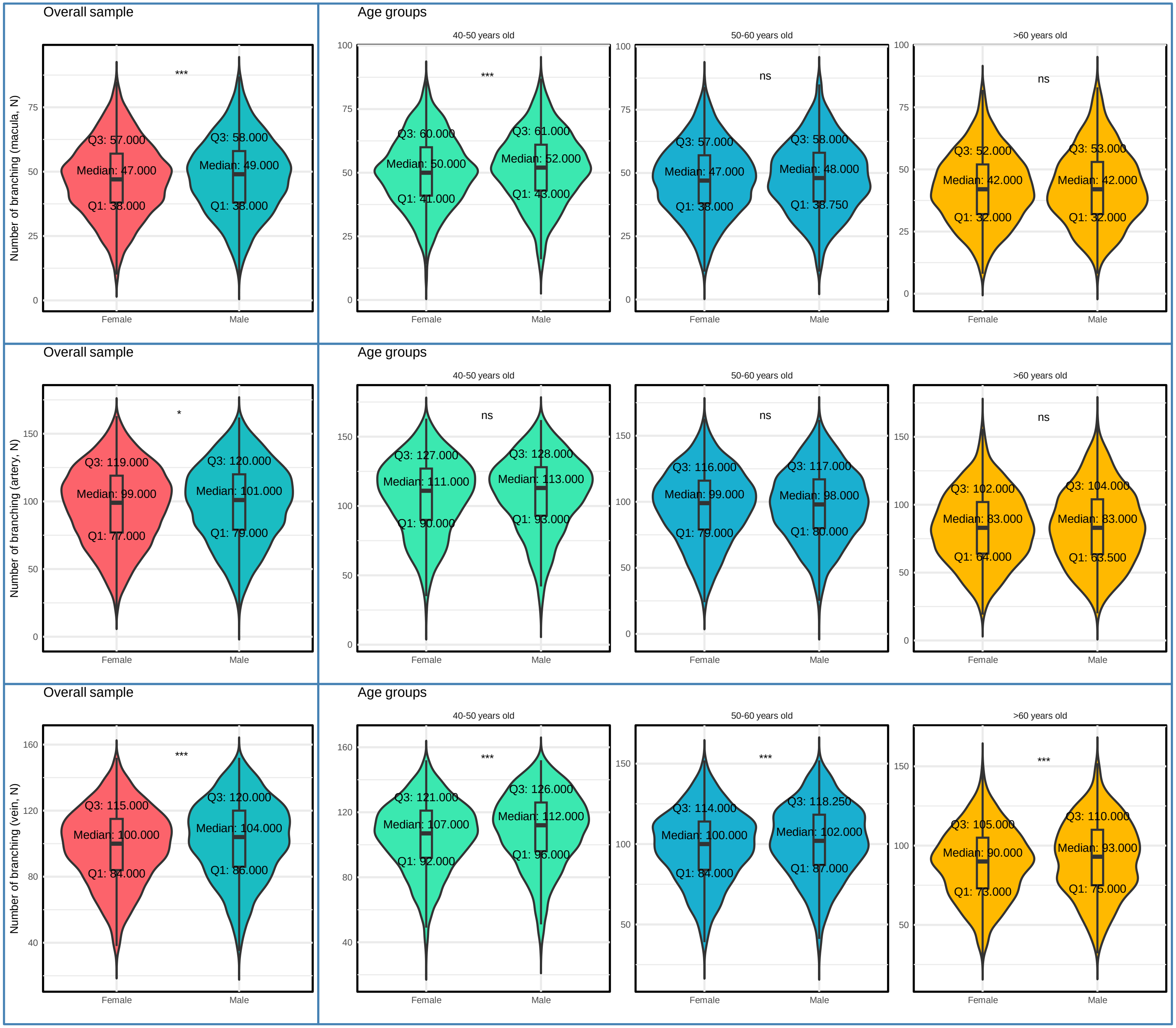
Overall sample
Age groups
40-50 years old
50-60 years old
>60 years old
Number of branching (macula, N)
Number of branching (artery, N)
Number of branching (vein, N)
Female
Male
***
*
ns
Q3: 57.000
Median: 47.000
Q1: 38.000
Q3: 58.000
Median: 49.000
Q1: 38.000
Q3: 60.000
Median: 50.000
Q1: 41.000
Q3: 61.000
Median: 52.000
Q1: 43.000
Q3: 57.000
Median: 47.000
Q1: 38.000
Q3: 58.000
Median: 48.000
Q1: 38.750
Q3: 52.000
Median: 42.000
Q1: 32.000
Q3: 53.000
Median: 42.000
Q1: 32.000
Q3: 119.000
Median: 99.000
Q1: 77.000
Q3: 120.000
Median: 101.000
Q1: 79.000
Q3: 127.000
Median: 111.000
Q1: 90.000
Q3: 128.000
Median: 113.000
Q1: 93.000
Q3: 116.000
Median: 99.000
Q1: 79.000
Q3: 117.000
Median: 98.000
Q1: 80.000
Q3: 102.000
Median: 83.000
Q1: 64.000
Q3: 104.000
Median: 83.000
Q1: 63.500
Q3: 115.000
Median: 100.000
Q1: 84.000
Q3: 120.000
Median: 104.000
Q1: 86.000
Q3: 121.000
Median: 107.000
Q1: 92.000
Q3: 126.000
Median: 112.000
Q1: 96.000
Q3: 114.000
Median: 100.000
Q1: 84.000
Q3: 118.250
Median: 102.000
Q1: 87.000
Q3: 105.000
Median: 90.000
Q1: 73.000
Q3: 110.000
Median: 93.000
Q1: 75.000

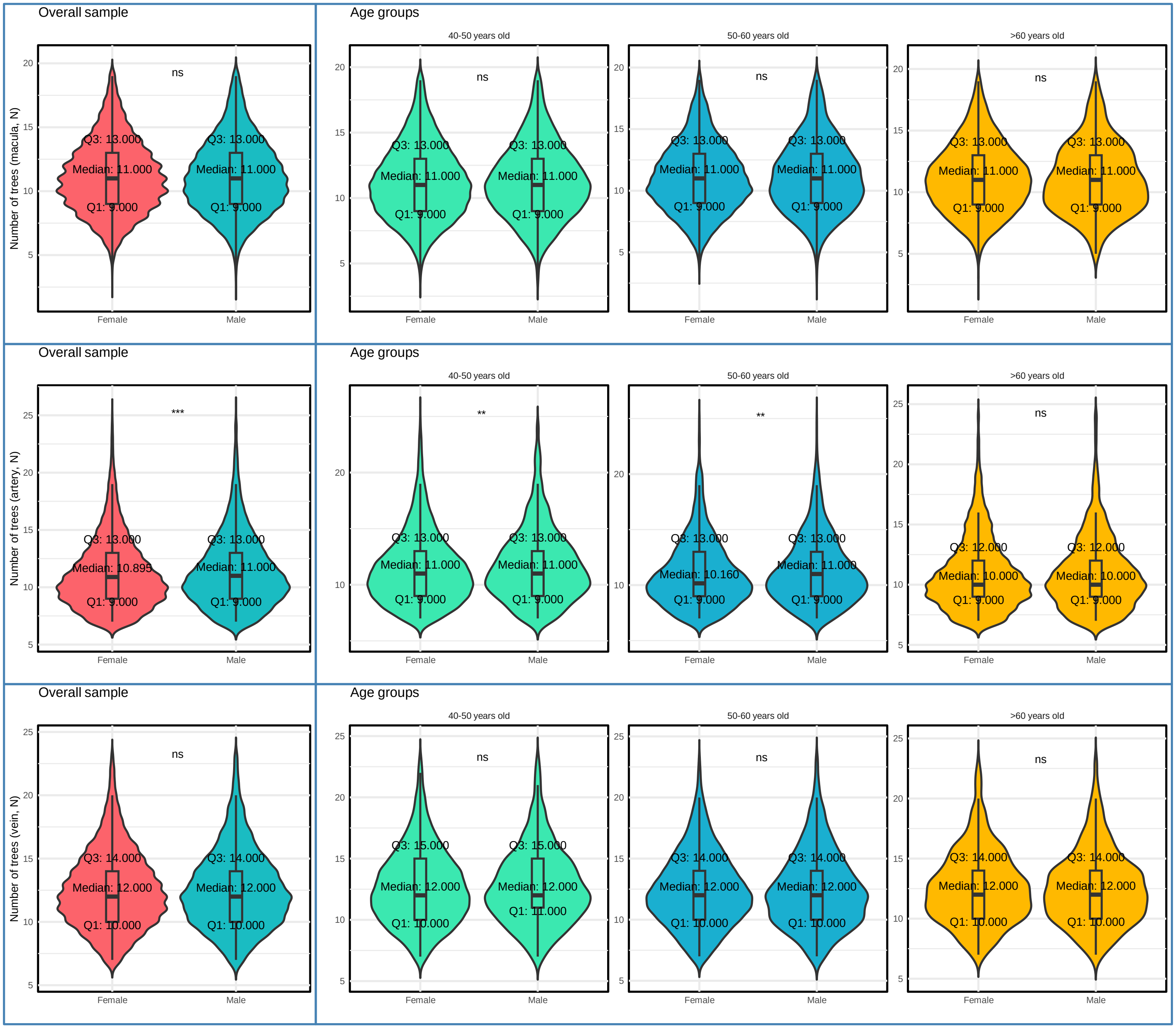

Overall sample
Age groups
40-50 years old
50-60 years old
>60 years old
Number of trees (macula, N)
Number of trees (artery, N)
Number of trees (vein, N)
Female
Male
ns
***
**
Q3: 13.000
Median: 11.000
Q1: 9.000
Median: 10.895
Median: 10.160
Q3: 12.000
Median: 10.000
Q3: 14.000
Median: 12.000
Q1: 10.000
Q3: 15.000
Q1: 11.000

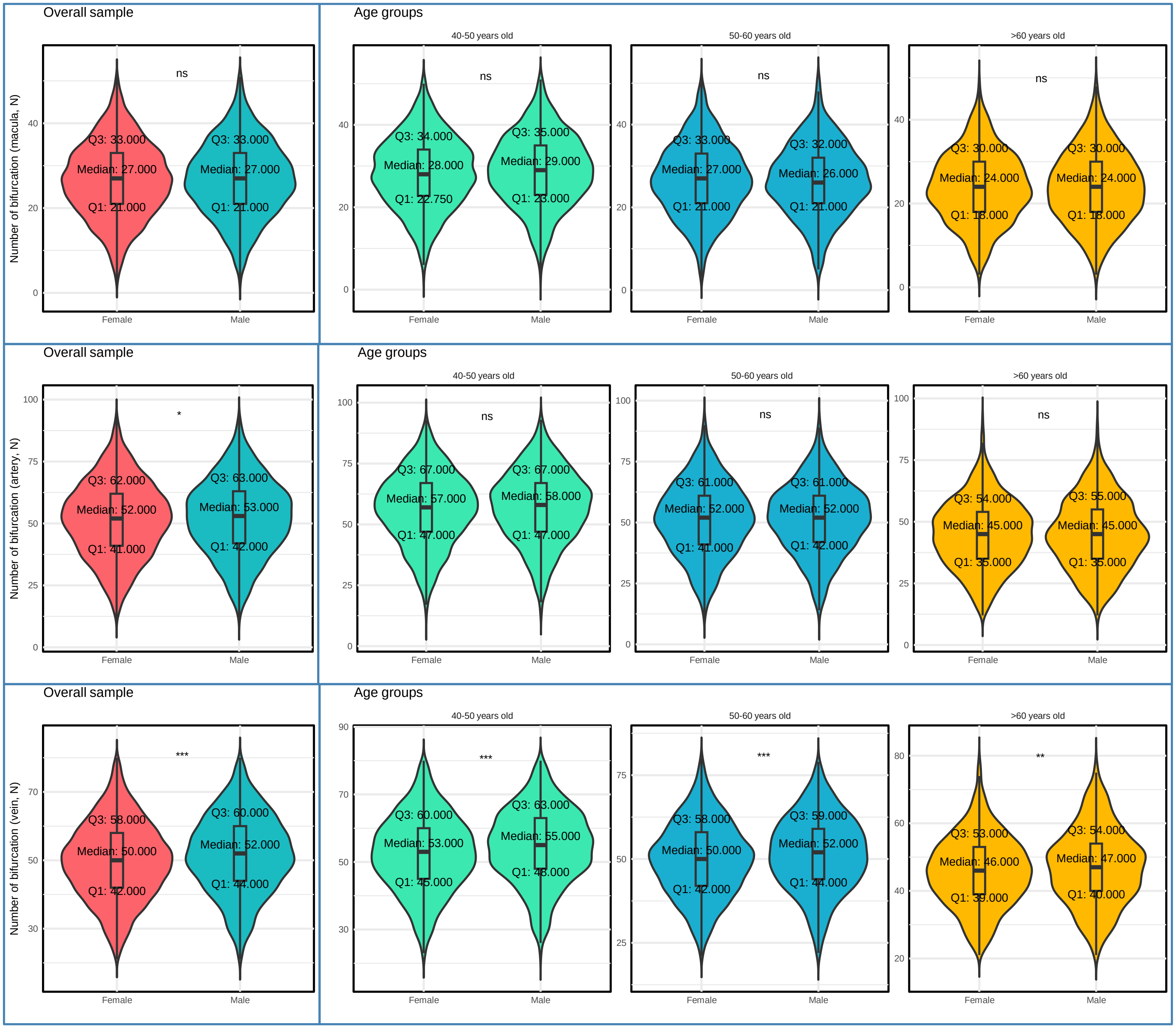
Overall sample
Age groups
40-50 years old
50-60 years old
>60 years old
Number of bifurcation (macula, N)
Number of bifurcation (artery, N)
Number of bifurcation (vein, N)
Female
Male
ns
*
**
***
Q3: 33.000
Median: 27.000
Q1: 21.000
Q3: 34.000
Median: 28.000
Q1: 22.750
Q3: 35.000
Median: 29.000
Q1: 23.000
Q3: 32.000
Median: 26.000
Q3: 30.000
Median: 24.000
Q1: 18.000
Q3: 62.000
Median: 52.000
Q1: 41.000
Q3: 63.000
Median: 53.000
Q1: 42.000
Q3: 67.000
Median: 57.000
Q1: 47.000
Median: 58.000
Q3: 61.000
Q3: 54.000
Median: 45.000
Q1: 35.000
Q3: 55.000
Q3: 58.000
Median: 50.000
Q1: 42.000
Q3: 60.000
Median: 52.000
Q1: 44.000
Median: 53.000
Q1: 45.000
Q3: 63.000
Median: 55.000
Q1: 48.000
Q3: 59.000
Q3: 53.000
Median: 46.000
Q1: 39.000
Median: 47.000
Q1: 40.000

### Supplementary Figure 3. Distribution of Density Measurements

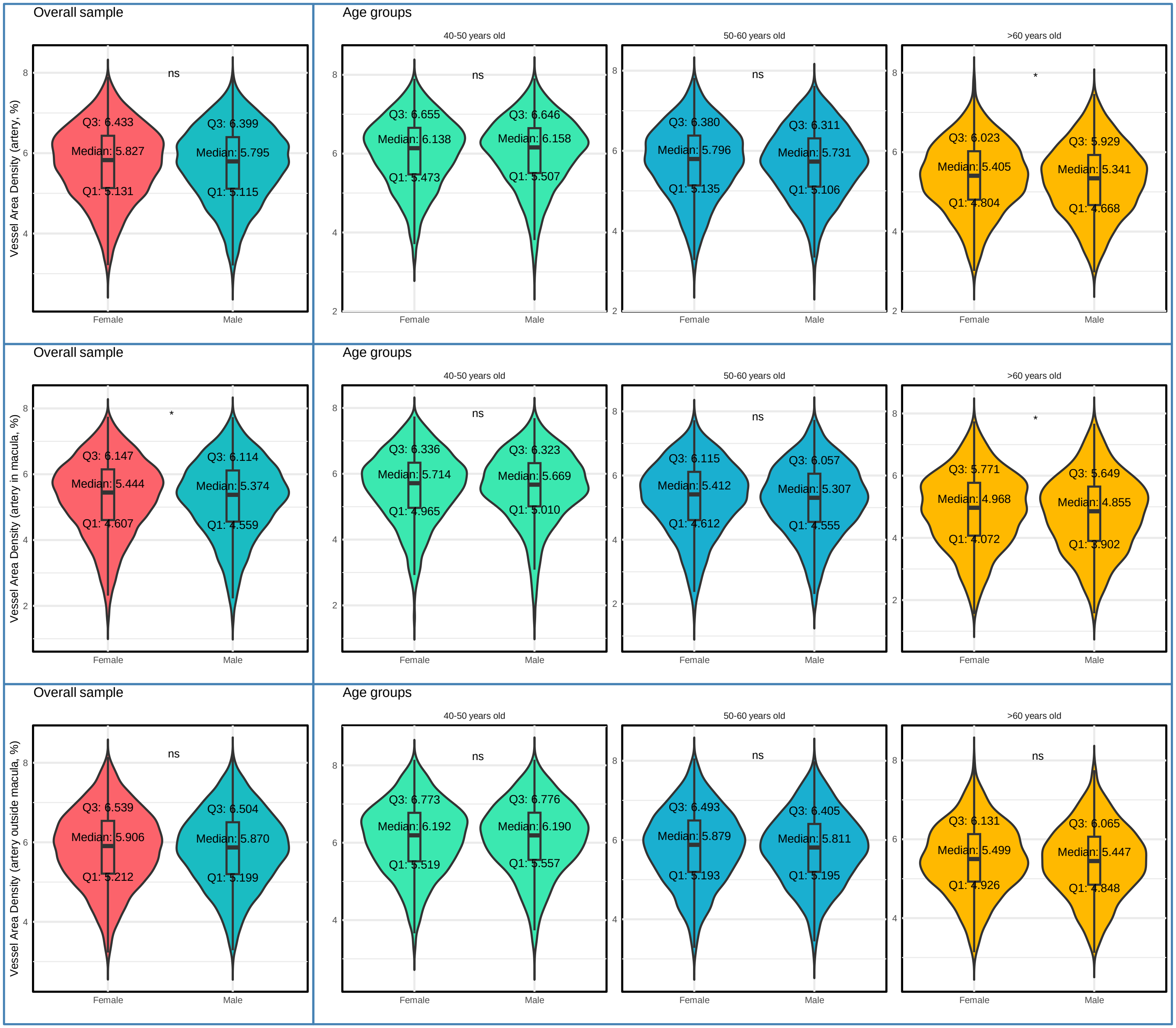

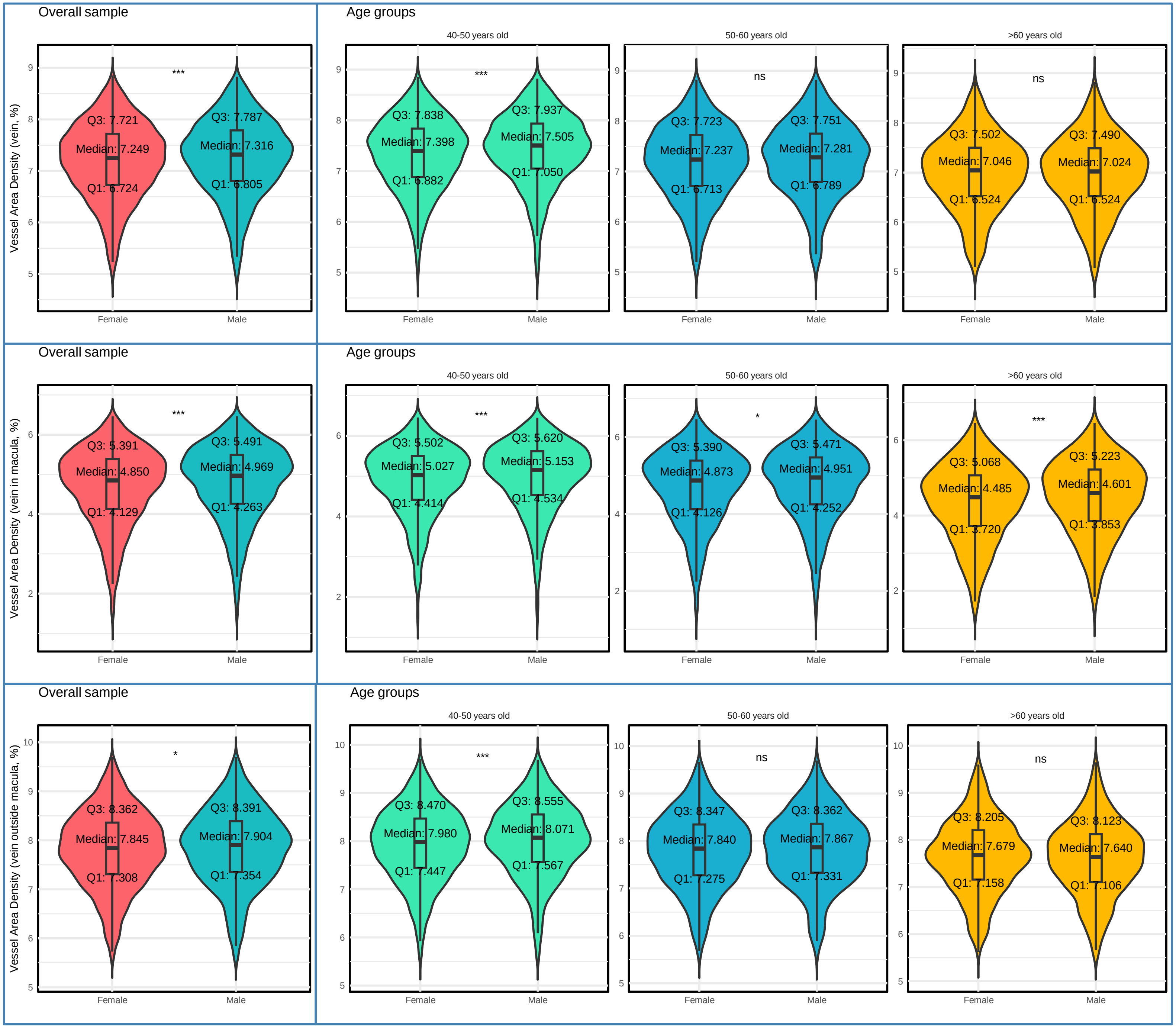

Overall sample
Age groups
40-50 years old
50-60 years old
>60 years old
Vessel Area Density (vein, %)
***
ns
ns
Q3: 7.721
Median: 7.249
Q1: 6.724
Q3: 7.787
Median: 7.316
Q1: 6.805
Q3: 7.838
Median: 7.398
Q1: 6.882
Q3: 7.937
Median: 7.505
Q1: 7.050
Q3: 7.723
Median: 7.237
Q1: 6.713
Q3: 7.751
Median: 7.281
Q1: 6.789
Q3: 7.502
Median: 7.046
Q1: 6.524
Q3: 7.490
Median: 7.024
Q1: 6.524
Female
Male
Vessel Area Density (vein in macula, %)
*
Q3: 5.391
Median: 4.850
Q1: 4.129
Q3: 5.491
Median: 4.969
Q1: 4.263
Q3: 5.502
Median: 5.027
Q1: 4.414
Q3: 5.620
Median: 5.153
Q1: 4.534
Q3: 5.390
Median: 4.873
Q1: 4.126
Q3: 5.471
Median: 4.951
Q1: 4.252
Q3: 5.068
Median: 4.485
Q1: 3.720
Q3: 5.223
Median: 4.601
Q1: 3.853
Vessel Area Density (vein outside macula, %)
Q3: 8.362
Median: 7.845
Q1: 7.308
Q3: 8.391
Median: 7.904
Q1: 7.354
Q3: 8.470
Median: 7.980
Q1: 7.447
Q3: 8.555
Median: 8.071
Q1: 7.567
Q3: 8.347
Median: 7.840
Q1: 7.275
Q3: 8.362
Median: 7.867
Q1: 7.331
Q3: 8.205
Median: 7.679
Q1: 7.158
Q3: 8.123
Median: 7.640
Q1: 7.106

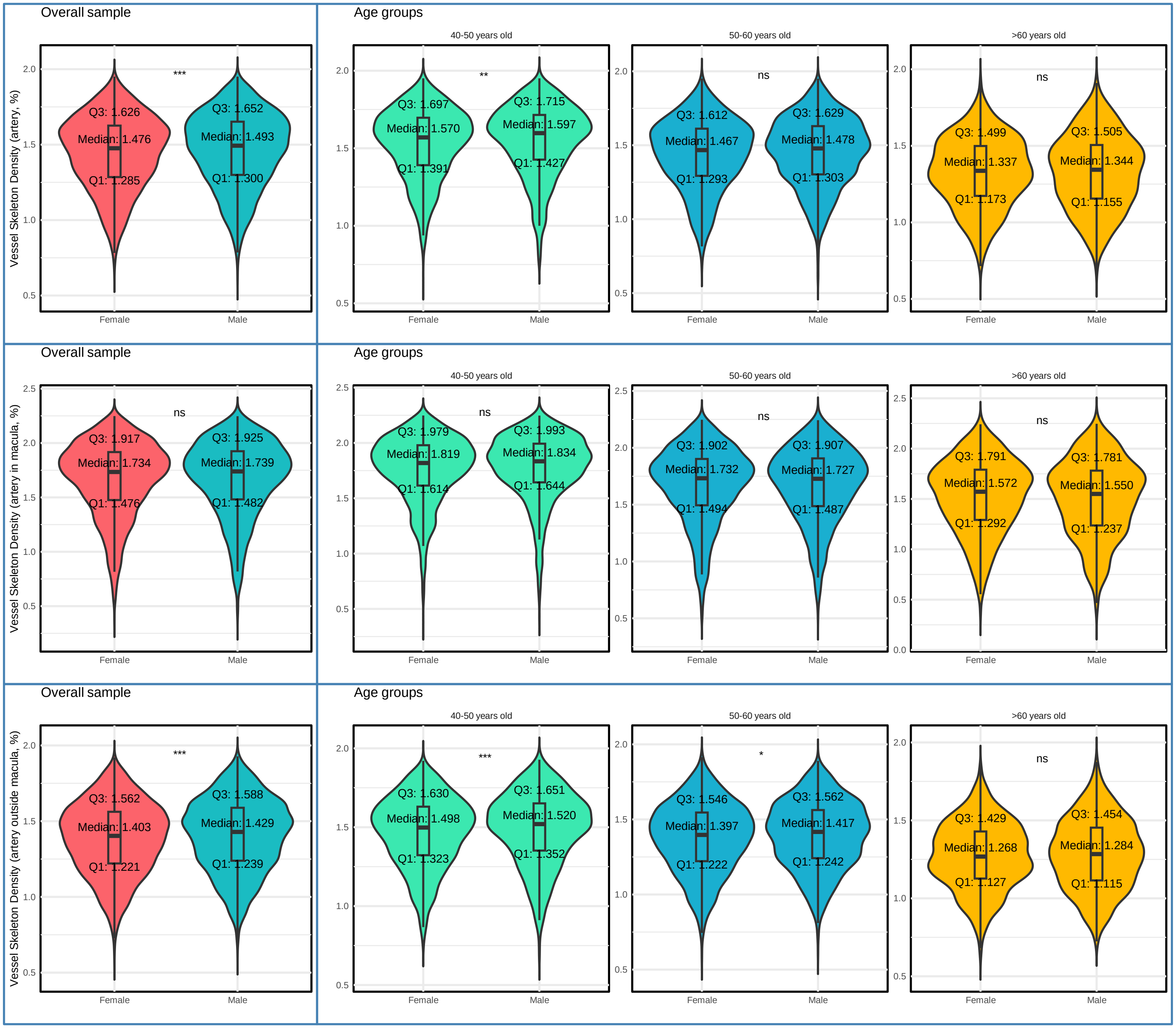
Overall sample
Age groups
40-50 years old
50-60 years old
>60 years old
Vessel Skeleton Density (artery, %)
***
Q3: 1.626
Median: 1.476
Q1: 1.285
Q3: 1.652
Median: 1.493
Q1: 1.300
**
Q3: 1.697
Median: 1.570
Q1: 1.391
Q3: 1.715
Median: 1.597
Q1: 1.427
ns
Q3: 1.612
Median: 1.467
Q1: 1.293
Q3: 1.629
Median: 1.478
Q1: 1.303
ns
Q3: 1.499
Median: 1.337
Q1: 1.173
Q3: 1.505
Median: 1.344
Q1: 1.155
Vessel Skeleton Density (artery in macula, %)
ns
Q3: 1.917
Median: 1.734
Q1: 1.476
Q3: 1.925
Median: 1.739
Q1: 1.482
ns
Q3: 1.979
Median: 1.819
Q1: 1.614
Q3: 1.993
Median: 1.834
Q1: 1.644
ns
Q3: 1.902
Median: 1.732
Q1: 1.494
Q3: 1.907
Median: 1.727
Q1: 1.487
ns
Q3: 1.791
Median: 1.572
Q1: 1.292
Q3: 1.781
Median: 1.550
Q1: 1.237
Vessel Skeleton Density (artery outside macula, %)
***
Q3: 1.562
Median: 1.403
Q1: 1.221
Q3: 1.588
Median: 1.429
Q1: 1.239
***
Q3: 1.630
Median: 1.498
Q1: 1.323
Q3: 1.651
Median: 1.520
Q1: 1.352
*
Q3: 1.546
Median: 1.397
Q1: 1.222
Q3: 1.562
Median: 1.417
Q1: 1.242
ns
Q3: 1.429
Median: 1.268
Q1: 1.127
Q3: 1.454
Median: 1.284
Q1: 1.115
Female
Male

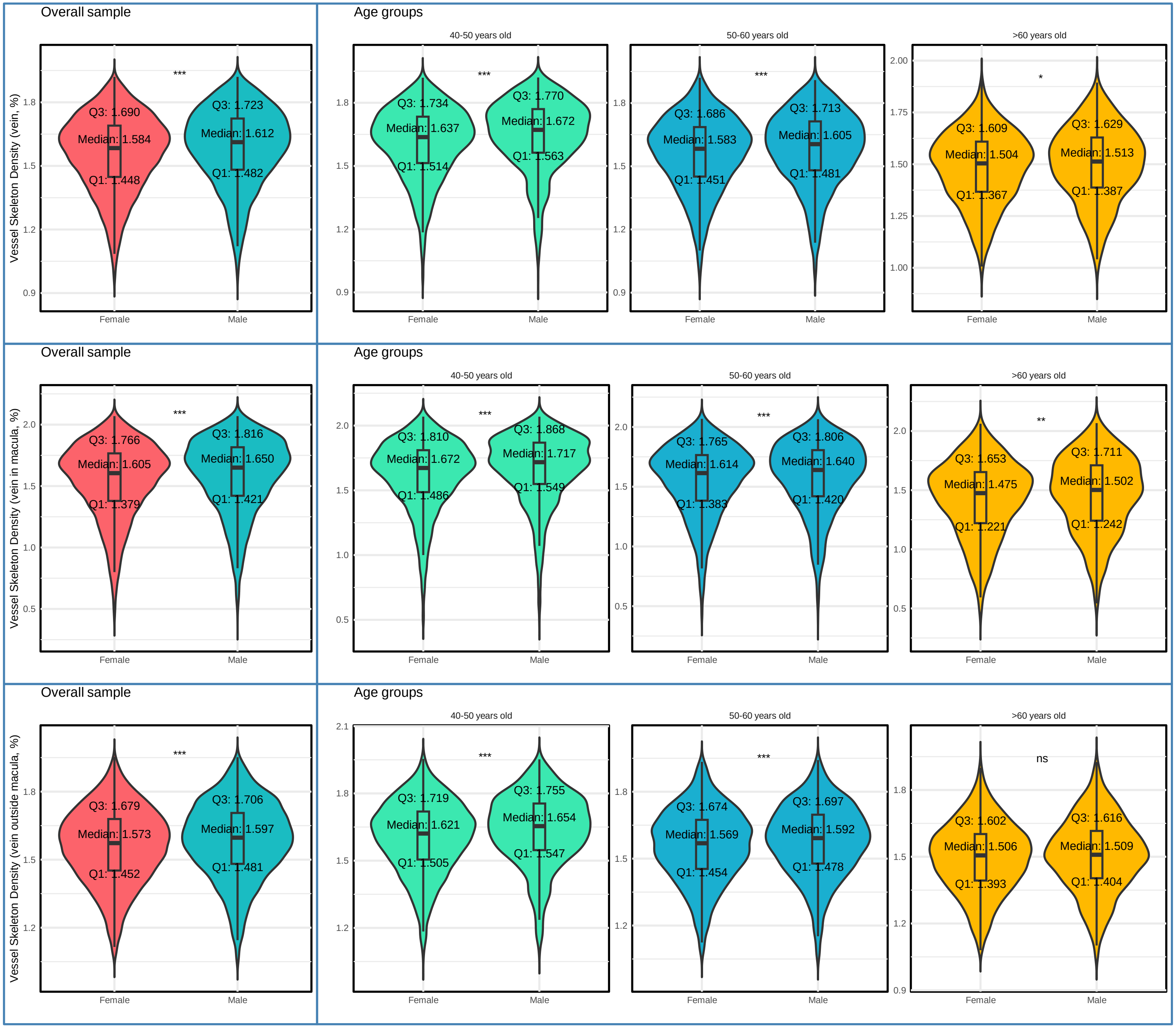
Overall sample
Age groups
40-50 years old
50-60 years old
>60 years old
Vessel Skeleton Density (vein, %)
Vessel Skeleton Density (vein in macula, %)
Vessel Skeleton Density (vein outside macula, %)
Female
Male
***
*
**
ns
Q3: 1.690
Median: 1.584
Q1: 1.448
Q3: 1.723
Median: 1.612
Q1: 1.482
Q3: 1.734
Median: 1.637
Q1: 1.514
Q3: 1.770
Median: 1.672
Q1: 1.563
Q3: 1.686
Median: 1.583
Q1: 1.451
Q3: 1.713
Median: 1.605
Q1: 1.481
Q3: 1.609
Median: 1.504
Q1: 1.367
Q3: 1.629
Median: 1.513
Q1: 1.387
Q3: 1.766
Median: 1.605
Q1: 1.379
Q3: 1.816
Median: 1.650
Q1: 1.421
Q3: 1.810
Median: 1.672
Q1: 1.486
Q3: 1.868
Median: 1.717
Q1: 1.549
Q3: 1.765
Median: 1.614
Q1: 1.383
Q3: 1.806
Median: 1.640
Q1: 1.420
Q3: 1.653
Median: 1.475
Q1: 1.221
Q3: 1.711
Median: 1.502
Q1: 1.242
Q3: 1.679
Median: 1.573
Q1: 1.452
Q3: 1.706
Median: 1.597
Q1: 1.481
Q3: 1.719
Median: 1.621
Q1: 1.505
Q3: 1.755
Median: 1.654
Q1: 1.547
Q3: 1.674
Median: 1.569
Q1: 1.454
Q3: 1.697
Median: 1.592
Q1: 1.478
Q3: 1.602
Median: 1.506
Q1: 1.393
Q3: 1.616
Median: 1.509
Q1: 1.404

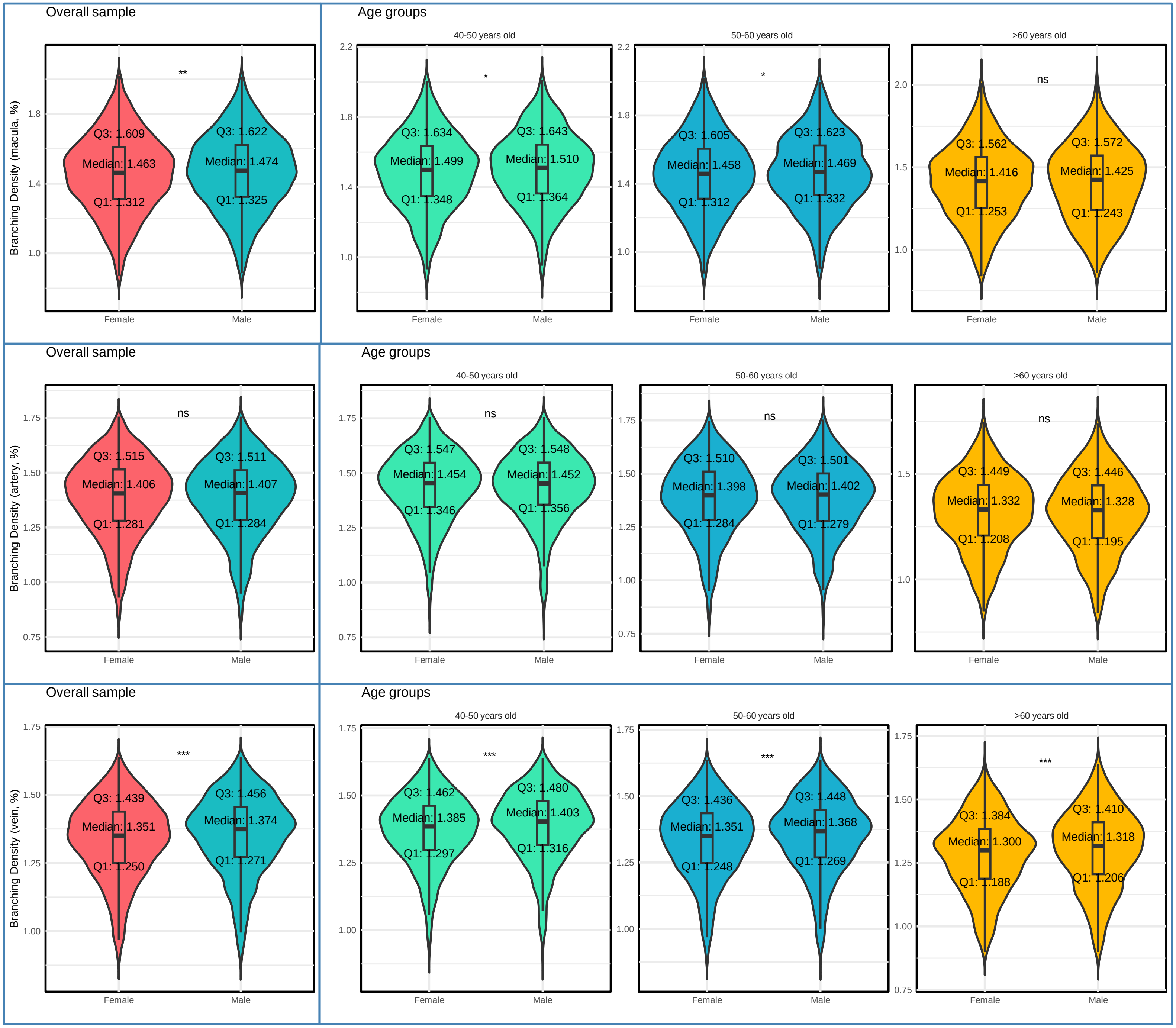
Overall sample
Age groups
40-50 years old
50-60 years old
>60 years old
Branching Density (macula, %)
**
Q3: 1.609
Median: 1.463
Q1: 1.312
Q3: 1.622
Median: 1.474
Q1: 1.325
*
Q3: 1.634
Median: 1.499
Q1: 1.348
Q3: 1.643
Median: 1.510
Q1: 1.364
*
Q3: 1.605
Median: 1.458
Q1: 1.312
Q3: 1.623
Median: 1.469
Q1: 1.332
ns
Q3: 1.562
Median: 1.416
Q1: 1.253
Q3: 1.572
Median: 1.425
Q1: 1.243
Overall sample
Age groups
40-50 years old
50-60 years old
>60 years old
Branching Density (artery, %)
ns
Q3: 1.515
Median: 1.406
Q1: 1.281
Q3: 1.511
Median: 1.407
Q1: 1.284
ns
Q3: 1.547
Median: 1.454
Q1: 1.346
Q3: 1.548
Median: 1.452
Q1: 1.356
ns
Q3: 1.510
Median: 1.398
Q1: 1.284
Q3: 1.501
Median: 1.402
Q1: 1.279
ns
Q3: 1.449
Median: 1.332
Q1: 1.208
Q3: 1.446
Median: 1.328
Q1: 1.195
Overall sample
Age groups
40-50 years old
50-60 years old
>60 years old
Branching Density (vein, %)
***
Q3: 1.439
Median: 1.351
Q1: 1.250
Q3: 1.456
Median: 1.374
Q1: 1.271
***
Q3: 1.462
Median: 1.385
Q1: 1.297
Q3: 1.480
Median: 1.403
Q1: 1.316
***
Q3: 1.436
Median: 1.351
Q1: 1.248
Q3: 1.448
Median: 1.368
Q1: 1.269
***
Q3: 1.384
Median: 1.300
Q1: 1.188
Q3: 1.410
Median: 1.318
Q1: 1.206
Female
Male

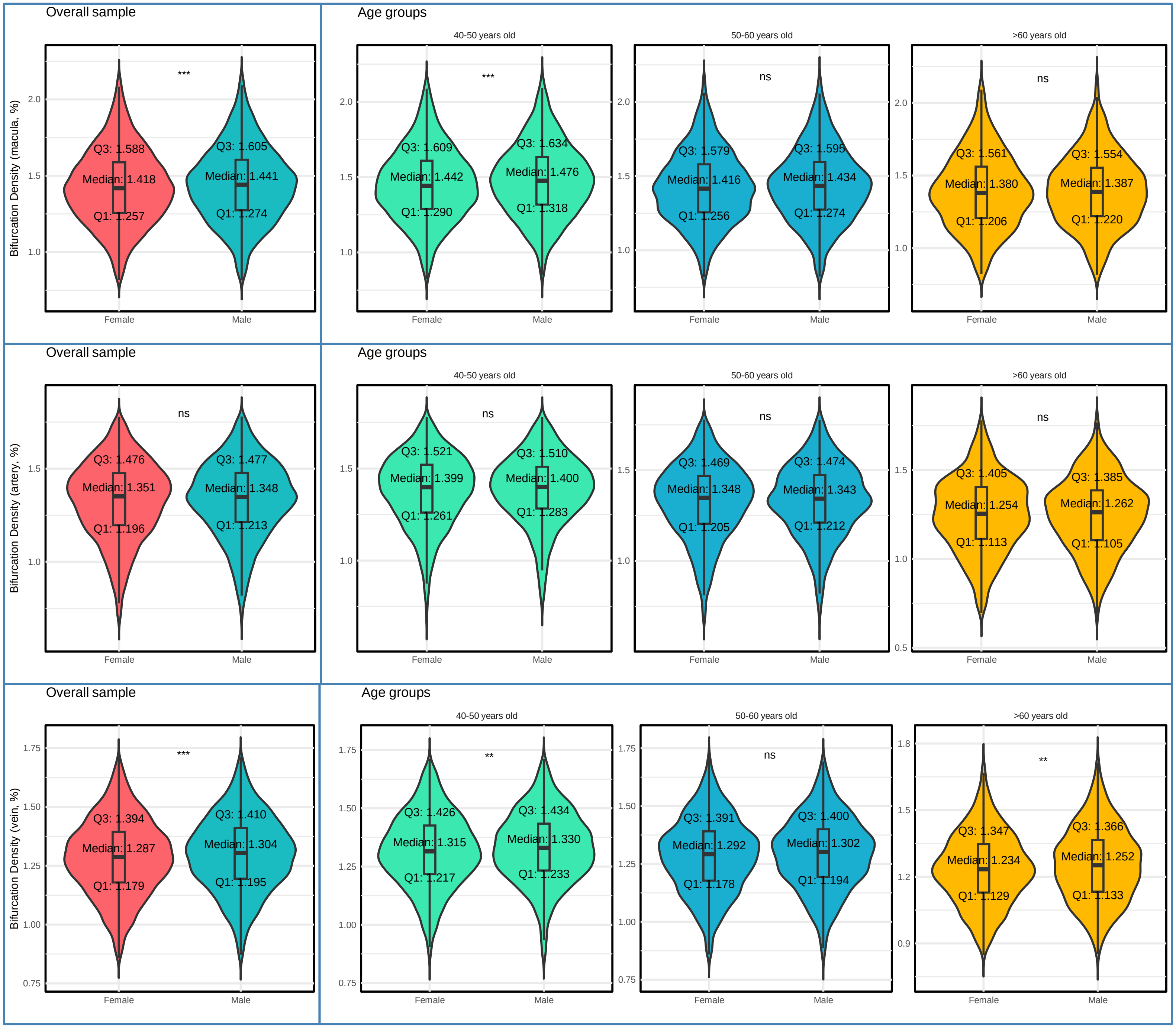

Overall sample
Age groups
40-50 years old
50-60 years old
>60 years old
Bifurcation Density (macula, %)
***
Q3: 1.588
Median: 1.418
Q1: 1.257
Q3: 1.605
Median: 1.441
Q1: 1.274
***
Q3: 1.609
Median: 1.442
Q1: 1.290
Q3: 1.634
Median: 1.476
Q1: 1.318
ns
Q3: 1.579
Median: 1.416
Q1: 1.256
Q3: 1.595
Median: 1.434
Q1: 1.274
ns
Q3: 1.561
Median: 1.380
Q1: 1.206
Q3: 1.554
Median: 1.387
Q1: 1.220
Female
Male
Overall sample
Age groups
40-50 years old
50-60 years old
>60 years old
Bifurcation Density (artery, %)
ns
Q3: 1.476
Median: 1.351
Q1: 1.196
Q3: 1.477
Median: 1.348
Q1: 1.213
ns
Q3: 1.521
Median: 1.399
Q1: 1.261
Q3: 1.510
Median: 1.400
Q1: 1.283
ns
Q3: 1.469
Median: 1.348
Q1: 1.205
Q3: 1.474
Median: 1.343
Q1: 1.212
ns
Q3: 1.405
Median: 1.254
Q1: 1.113
Q3: 1.385
Median: 1.262
Q1: 1.105
Female
Male
Overall sample
Age groups
40-50 years old
50-60 years old
>60 years old
Bifurcation Density (vein, %)
***
Q3: 1.394
Median: 1.287
Q1: 1.179
Q3: 1.410
Median: 1.304
Q1: 1.195
**
Q3: 1.426
Median: 1.315
Q1: 1.217
Q3: 1.434
Median: 1.330
Q1: 1.233
ns
Q3: 1.391
Median: 1.292
Q1: 1.178
Q3: 1.400
Median: 1.302
Q1: 1.194
**
Q3: 1.347
Median: 1.234
Q1: 1.129
Q3: 1.366
Median: 1.252
Q1: 1.133
Female
Male

### Supplementary Figure 4. Distribution of Branching Angle Measurements

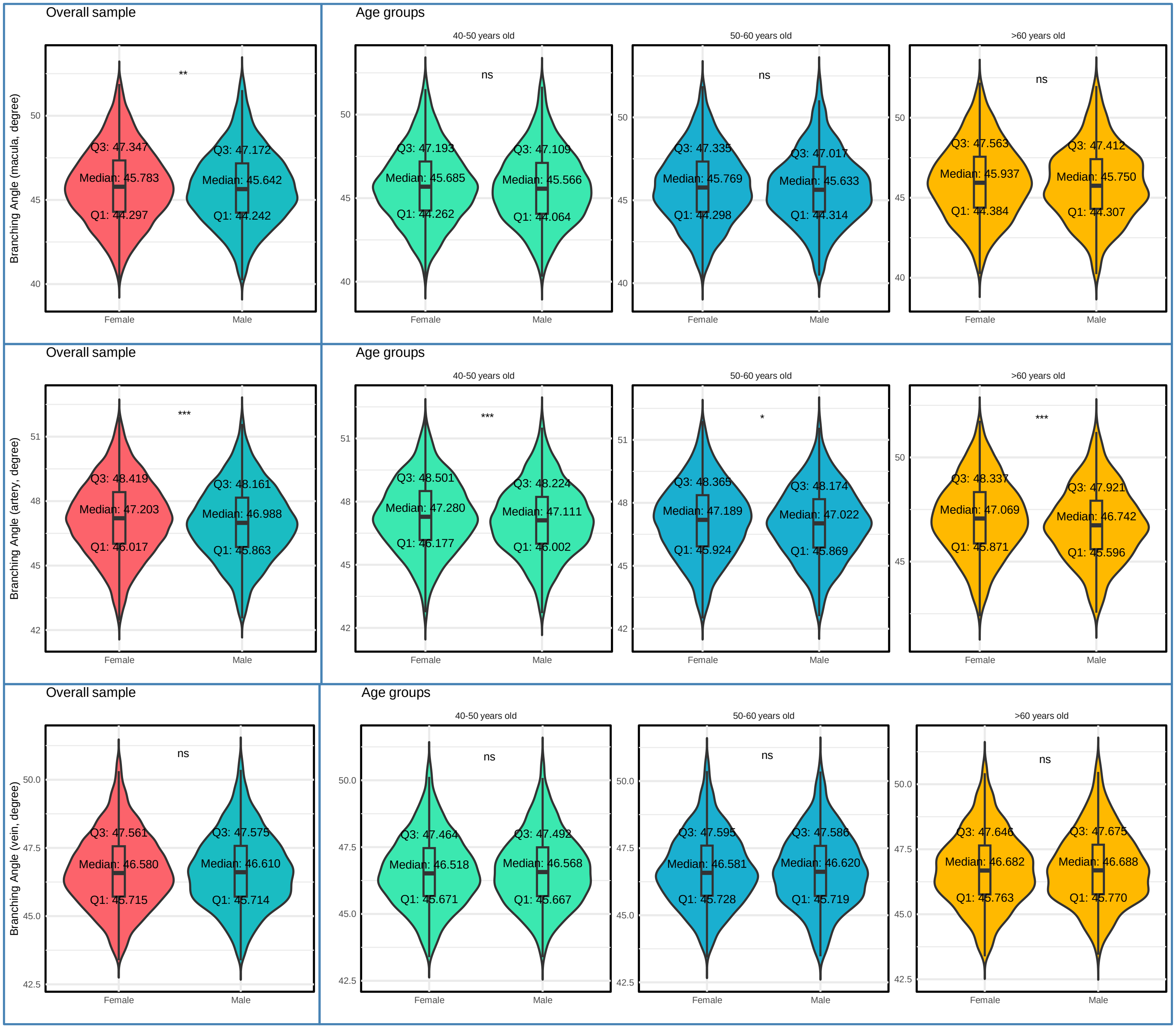

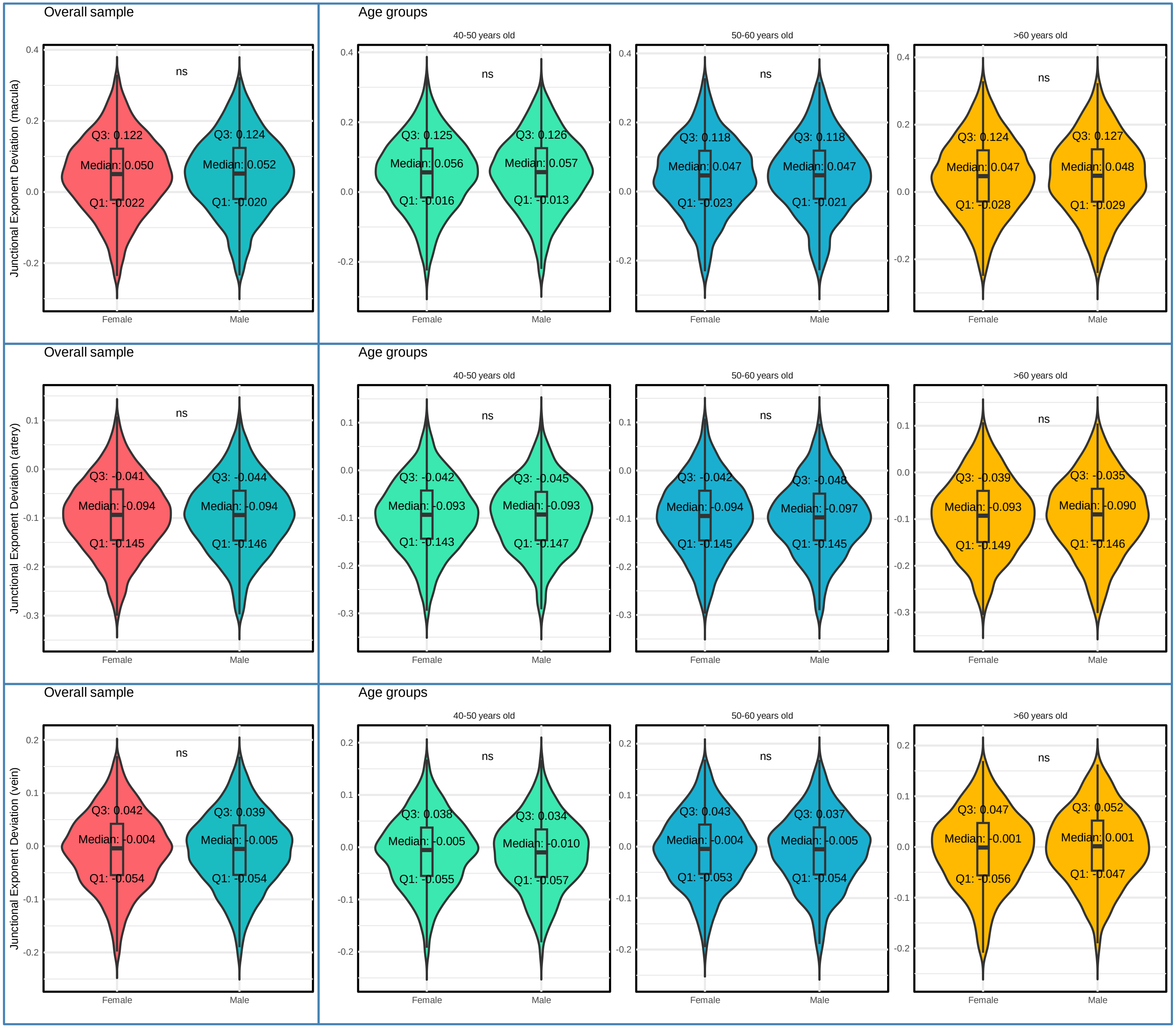

Overall sample
Age groups
40-50 years old
50-60 years old
>60 years old
Junctional Exponent Deviation (macula)
ns
Q3: 0.122
Median: 0.050
Q1: -0.022
Q3: 0.124
Median: 0.052
Q1: -0.020
Q3: 0.125
Median: 0.056
Q1: -0.016
Q3: 0.126
Median: 0.057
Q1: -0.013
Q3: 0.118
Median: 0.047
Q1: -0.023
Q3: 0.118
Median: 0.047
Q1: -0.021
Q3: 0.124
Median: 0.047
Q1: -0.028
Q3: 0.127
Median: 0.048
Q1: -0.029
Female
Male
Overall sample
Age groups
40-50 years old
50-60 years old
>60 years old
Junctional Exponent Deviation (artery)
ns
Q3: -0.041
Median: -0.094
Q1: -0.145
Q3: -0.044
Median: -0.094
Q1: -0.146
Q3: -0.042
Median: -0.093
Q1: -0.143
Q3: -0.045
Median: -0.093
Q1: -0.147
Q3: -0.042
Median: -0.094
Q1: -0.145
Q3: -0.048
Median: -0.097
Q1: -0.145
Q3: -0.039
Median: -0.093
Q1: -0.149
Q3: -0.035
Median: -0.090
Q1: -0.146
Female
Male
Overall sample
Age groups
40-50 years old
50-60 years old
>60 years old
Junctional Exponent Deviation (vein)
ns
Q3: 0.042
Median: -0.004
Q1: -0.054
Q3: 0.039
Median: -0.005
Q1: -0.054
Q3: 0.038
Median: -0.005
Q1: -0.055
Q3: 0.034
Median: -0.010
Q1: -0.057
Q3: 0.043
Median: -0.004
Q1: -0.053
Q3: 0.037
Median: -0.005
Q1: -0.054
Q3: 0.047
Median: -0.001
Q1: -0.056
Q3: 0.052
Median: 0.001
Q1: -0.047
Female
Male

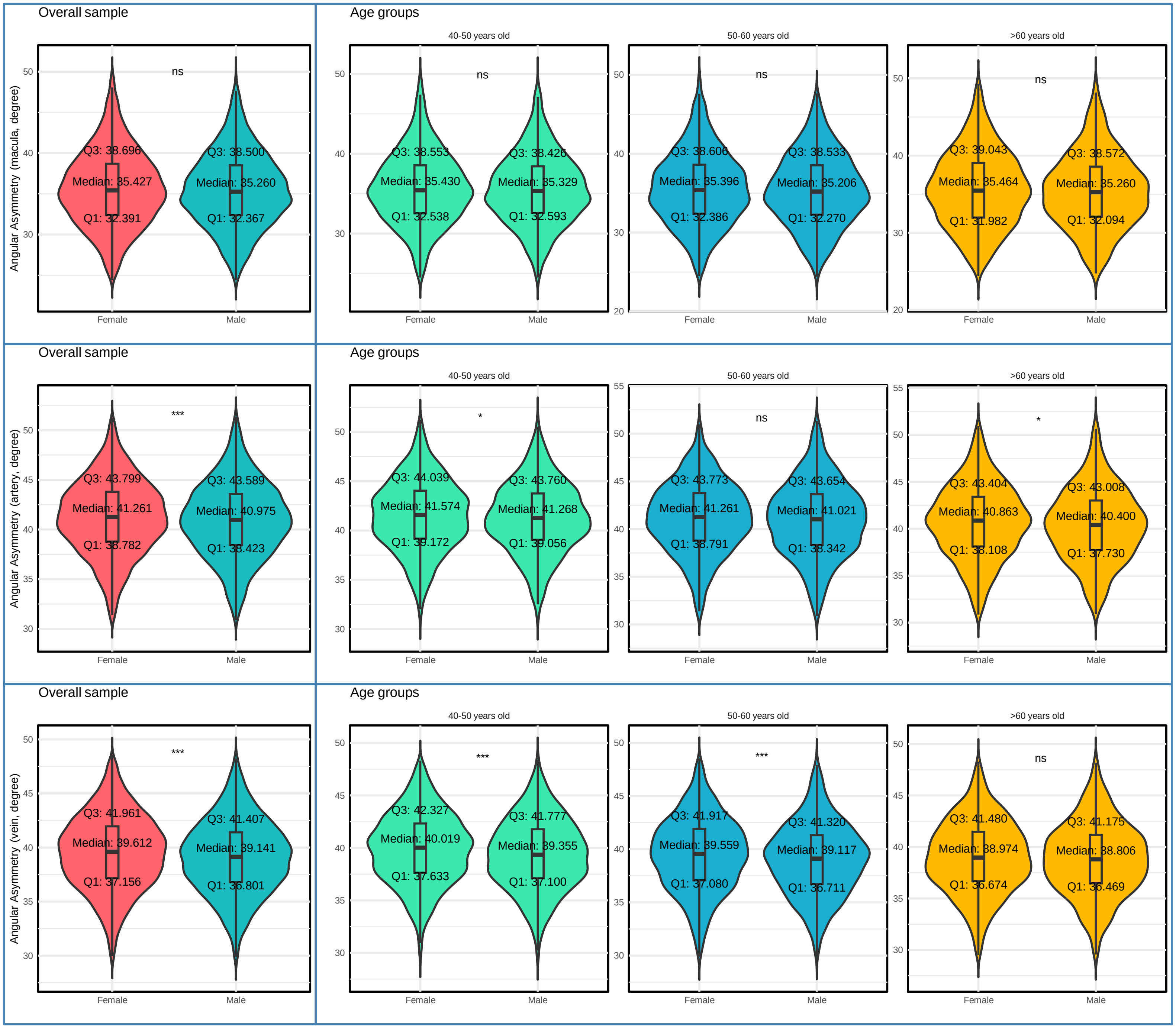

Overall sample
Age groups
40-50 years old
50-60 years old
>60 years old
Angular Asymmetry (macula, degree)
ns
Q3: 38.696
Median: 35.427
Q1: 32.391
Q3: 38.500
Median: 35.260
Q1: 32.367
Q3: 38.553
Median: 35.430
Q1: 32.538
Q3: 38.426
Median: 35.329
Q1: 32.593
Q3: 38.606
Median: 35.396
Q1: 32.386
Q3: 38.533
Median: 35.206
Q1: 32.270
Q3: 39.043
Median: 35.464
Q1: 31.982
Q3: 38.572
Median: 35.260
Q1: 32.094
Angular Asymmetry (artery, degree)
***
*
Q3: 43.799
Median: 41.261
Q1: 38.782
Q3: 43.589
Median: 40.975
Q1: 38.423
Q3: 44.039
Median: 41.574
Q1: 39.172
Q3: 43.760
Median: 41.268
Q1: 39.056
Q3: 43.773
Median: 41.261
Q1: 38.791
Q3: 43.654
Median: 41.021
Q1: 38.342
Q3: 43.404
Median: 40.863
Q1: 38.108
Q3: 43.008
Median: 40.400
Q1: 37.730
Angular Asymmetry (vein, degree)
Q3: 41.961
Median: 39.612
Q1: 37.156
Q3: 41.407
Median: 39.141
Q1: 36.801
Q3: 42.327
Median: 40.019
Q1: 37.633
Q3: 41.777
Median: 39.355
Q1: 37.100
Q3: 41.917
Median: 39.559
Q1: 37.080
Q3: 41.320
Median: 39.117
Q1: 36.711
Q3: 41.480
Median: 38.974
Q1: 36.674
Q3: 41.175
Median: 38.806
Q1: 36.469
Female
Male

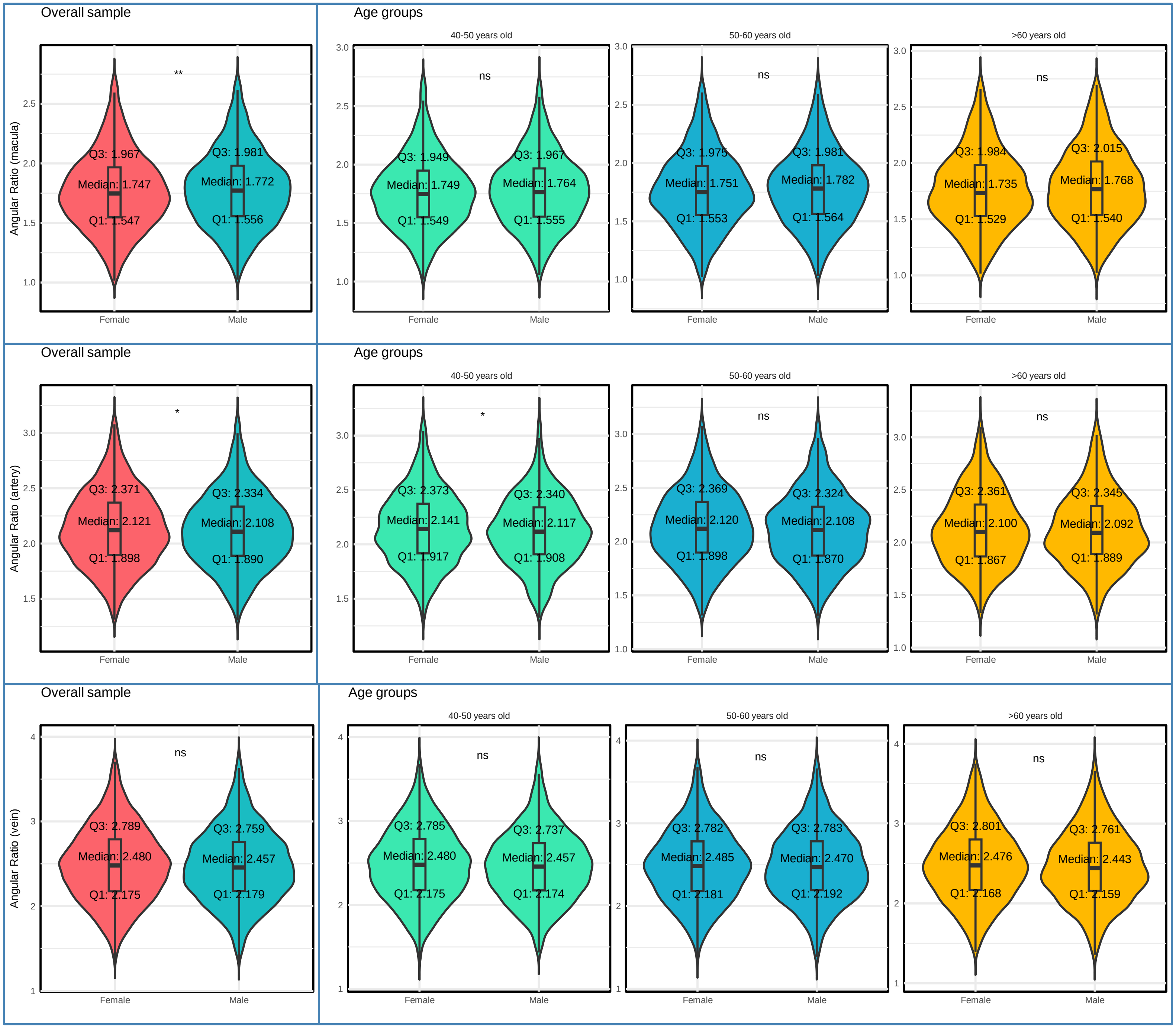

Overall sample
Angular Ratio (macula)
**
Q3: 1.967
Median: 1.747
Q1: 1.547
Q3: 1.981
Median: 1.772
Q1: 1.556
Female
Male
Age groups
40-50 years old
ns
Q3: 1.949
Median: 1.749
Q1: 1.549
Q3: 1.967
Median: 1.764
Q1: 1.555
50-60 years old
ns
Q3: 1.975
Median: 1.751
Q1: 1.553
Q3: 1.981
Median: 1.782
Q1: 1.564
>60 years old
ns
Q3: 1.984
Median: 1.735
Q1: 1.529
Q3: 2.015
Median: 1.768
Q1: 1.540
Overall sample
Angular Ratio (artery)
*
Q3: 2.371
Median: 2.121
Q1: 1.898
Q3: 2.334
Median: 2.108
Q1: 1.890
Age groups
40-50 years old
*
Q3: 2.373
Median: 2.141
Q1: 1.917
Q3: 2.340
Median: 2.117
Q1: 1.908
50-60 years old
ns
Q3: 2.369
Median: 2.120
Q1: 1.898
Q3: 2.324
Median: 2.108
Q1: 1.870
>60 years old
ns
Q3: 2.361
Median: 2.100
Q1: 1.867
Q3: 2.345
Median: 2.092
Q1: 1.889
Overall sample
Angular Ratio (vein)
ns
Q3: 2.789
Median: 2.480
Q1: 2.175
Q3: 2.759
Median: 2.457
Q1: 2.179
Age groups
40-50 years old
ns
Q3: 2.785
Median: 2.480
Q1: 2.175
Q3: 2.737
Median: 2.457
Q1: 2.174
50-60 years old
ns
Q3: 2.782
Median: 2.485
Q1: 2.181
Q3: 2.783
Median: 2.470
Q1: 2.192
>60 years old
ns
Q3: 2.801
Median: 2.476
Q1: 2.168
Q3: 2.761
Median: 2.443
Q1: 2.159

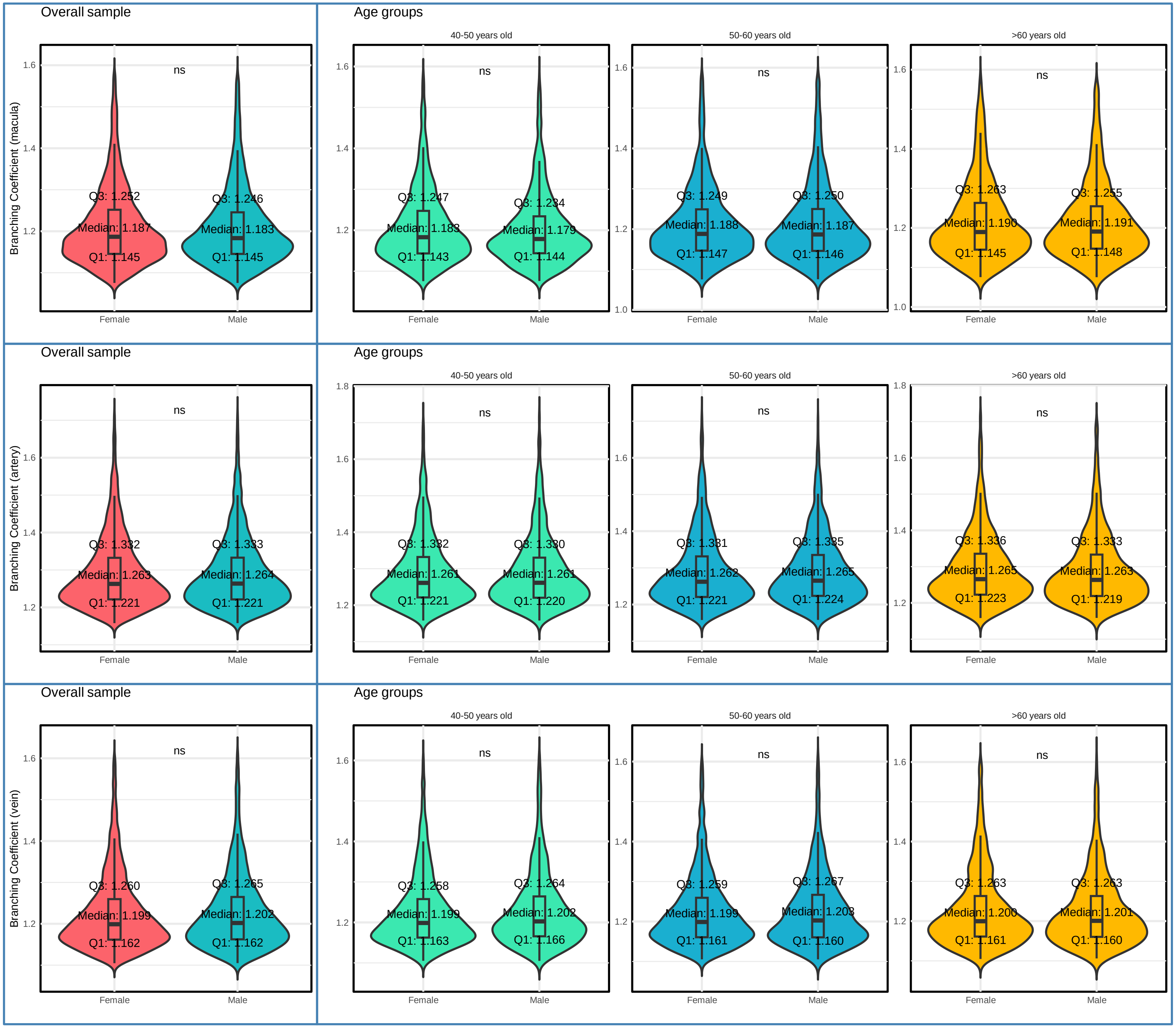

Overall sample
Age groups
40-50 years old
50-60 years old
>60 years old
Branching Coefficient (macula)
ns
Q3: 1.252
Median: 1.187
Q1: 1.145
Q3: 1.246
Median: 1.183
Q1: 1.145
Q3: 1.247
Median: 1.183
Q1: 1.143
Q3: 1.234
Median: 1.179
Q1: 1.144
Q3: 1.249
Median: 1.188
Q1: 1.147
Q3: 1.250
Median: 1.187
Q1: 1.146
Q3: 1.263
Median: 1.190
Q1: 1.145
Q3: 1.255
Median: 1.191
Q1: 1.148
Branching Coefficient (artery)
Q3: 1.332
Median: 1.263
Q1: 1.221
Q3: 1.333
Median: 1.264
Q1: 1.221
Q3: 1.332
Median: 1.261
Q1: 1.221
Q3: 1.330
Median: 1.261
Q1: 1.220
Q3: 1.331
Median: 1.262
Q1: 1.221
Q3: 1.335
Median: 1.265
Q1: 1.224
Q3: 1.336
Median: 1.265
Q1: 1.223
Q3: 1.333
Median: 1.263
Q1: 1.219
Branching Coefficient (vein)
Q3: 1.260
Median: 1.199
Q1: 1.162
Q3: 1.265
Median: 1.202
Q1: 1.162
Q3: 1.258
Median: 1.199
Q1: 1.163
Q3: 1.264
Median: 1.202
Q1: 1.166
Q3: 1.259
Median: 1.199
Q1: 1.161
Q3: 1.267
Median: 1.203
Q1: 1.160
Q3: 1.263
Median: 1.200
Q1: 1.161
Q3: 1.263
Median: 1.201
Q1: 1.160
Female
Male

### Supplementary Figure 5. Distribution of Tortuosity Measurements

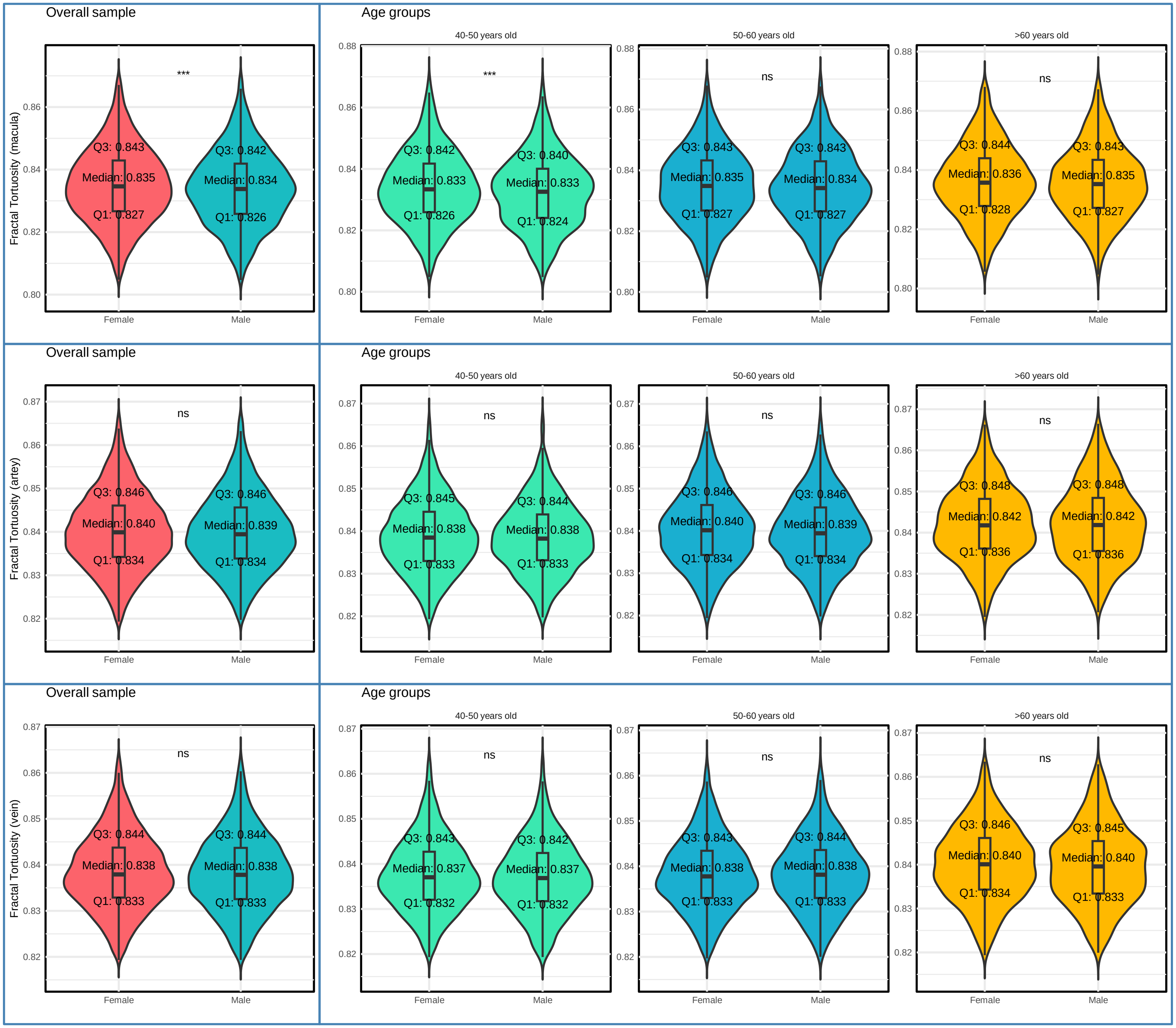

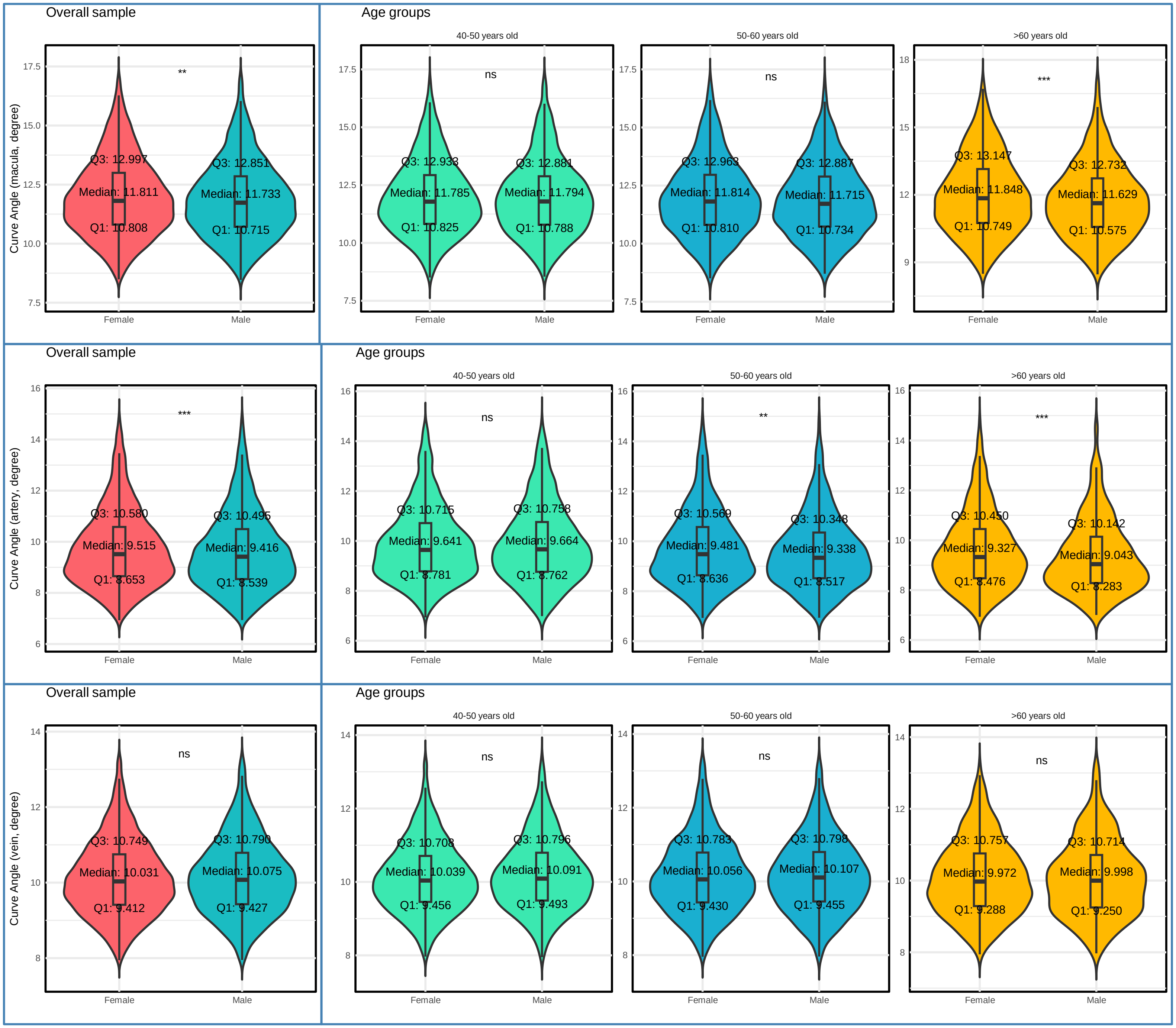
Overall sample
Age groups
40-50 years old
50-60 years old
>60 years old
Curve Angle (macula, degree)
**
ns
ns
***
Q3: 12.997
Median: 11.811
Q1: 10.808
Q3: 12.851
Median: 11.733
Q1: 10.715
Q3: 12.933
Median: 11.785
Q1: 10.825
Q3: 12.881
Median: 11.794
Q1: 10.788
Q3: 12.963
Median: 11.814
Q1: 10.810
Q3: 12.887
Median: 11.715
Q1: 10.734
Q3: 13.147
Median: 11.848
Q1: 10.749
Q3: 12.732
Median: 11.629
Q1: 10.575
Female
Male
Overall sample
Age groups
40-50 years old
50-60 years old
>60 years old
Curve Angle (artery, degree)
***
ns
**
***
Q3: 10.580
Median: 9.515
Q1: 8.653
Q3: 10.495
Median: 9.416
Q1: 8.539
Q3: 10.715
Median: 9.641
Q1: 8.781
Q3: 10.758
Median: 9.664
Q1: 8.762
Q3: 10.569
Median: 9.481
Q1: 8.636
Q3: 10.348
Median: 9.338
Q1: 8.517
Q3: 10.450
Median: 9.327
Q1: 8.476
Q3: 10.142
Median: 9.043
Q1: 8.283
Female
Male
Overall sample
Age groups
40-50 years old
50-60 years old
>60 years old
Curve Angle (vein, degree)
ns
ns
ns
ns
Q3: 10.749
Median: 10.031
Q1: 9.412
Q3: 10.790
Median: 10.075
Q1: 9.427
Q3: 10.708
Median: 10.039
Q1: 9.456
Q3: 10.796
Median: 10.091
Q1: 9.493
Q3: 10.783
Median: 10.056
Q1: 9.430
Q3: 10.798
Median: 10.107
Q1: 9.455
Q3: 10.757
Median: 9.972
Q1: 9.288
Q3: 10.714
Median: 9.998
Q1: 9.250
Female
Male

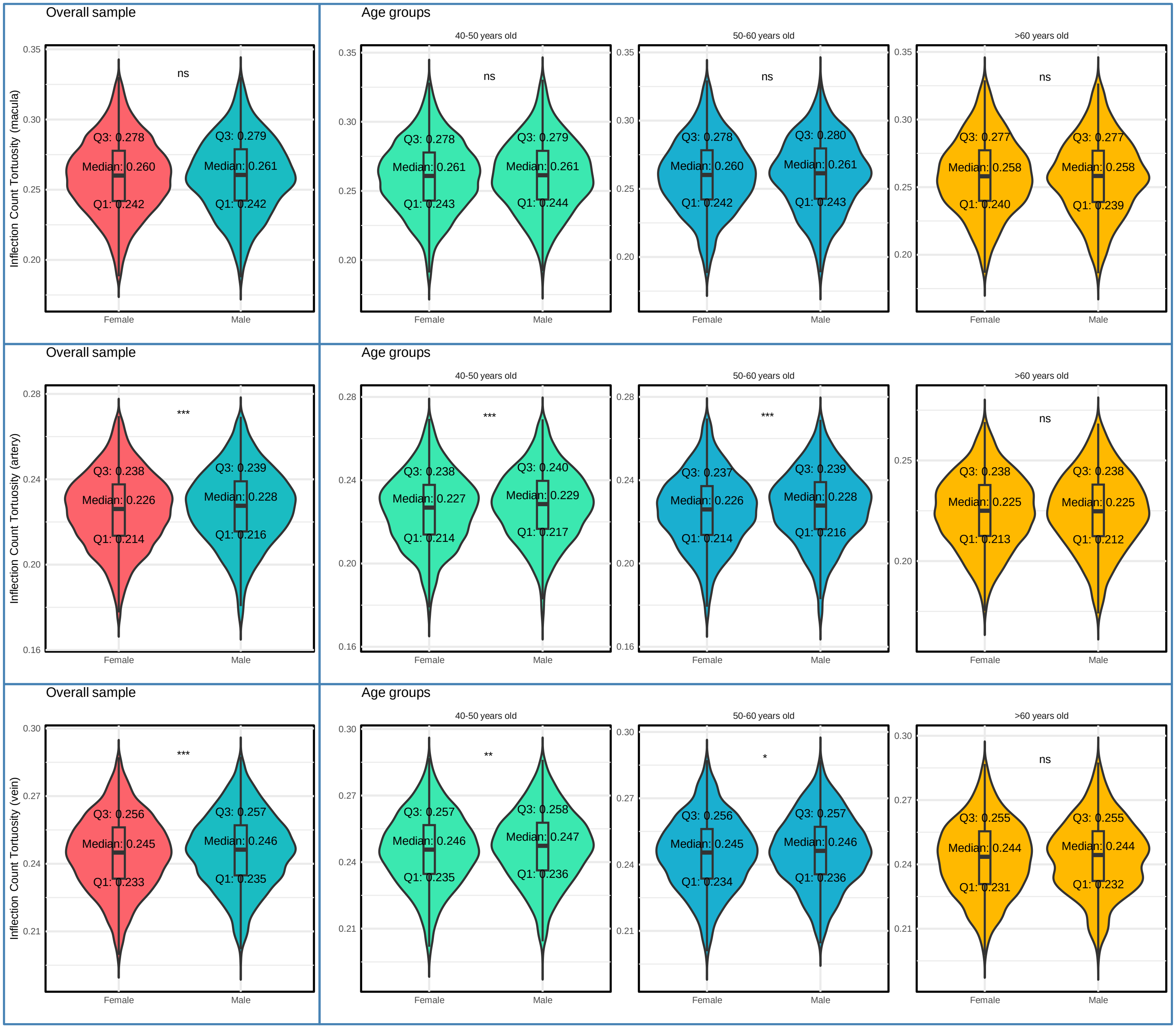
Overall sample
Age groups
40-50 years old
50-60 years old
>60 years old
Inflection Count Tortuosity (macula)
Inflection Count Tortuosity (artery)
Inflection Count Tortuosity (vein)
Female
Male
ns
***
**
*
Q3: 0.278
Median: 0.260
Q1: 0.242
Q3: 0.279
Median: 0.261
Q1: 0.242
Q3: 0.278
Median: 0.261
Q1: 0.243
Q3: 0.279
Median: 0.261
Q1: 0.244
Q3: 0.278
Median: 0.260
Q1: 0.242
Q3: 0.280
Median: 0.261
Q1: 0.243
Q3: 0.277
Median: 0.258
Q1: 0.240
Q3: 0.277
Median: 0.258
Q1: 0.239
Q3: 0.238
Median: 0.226
Q1: 0.214
Q3: 0.239
Median: 0.228
Q1: 0.216
Q3: 0.238
Median: 0.227
Q1: 0.214
Q3: 0.240
Median: 0.229
Q1: 0.217
Q3: 0.237
Median: 0.226
Q1: 0.214
Q3: 0.239
Median: 0.228
Q1: 0.216
Q3: 0.238
Median: 0.225
Q1: 0.213
Q3: 0.238
Median: 0.225
Q1: 0.212
Q3: 0.256
Median: 0.245
Q1: 0.233
Q3: 0.257
Median: 0.246
Q1: 0.235
Q3: 0.257
Median: 0.246
Q1: 0.235
Q3: 0.258
Median: 0.247
Q1: 0.236
Q3: 0.256
Median: 0.245
Q1: 0.234
Q3: 0.257
Median: 0.246
Q1: 0.236
Q3: 0.255
Median: 0.244
Q1: 0.231
Q3: 0.255
Median: 0.244
Q1: 0.232

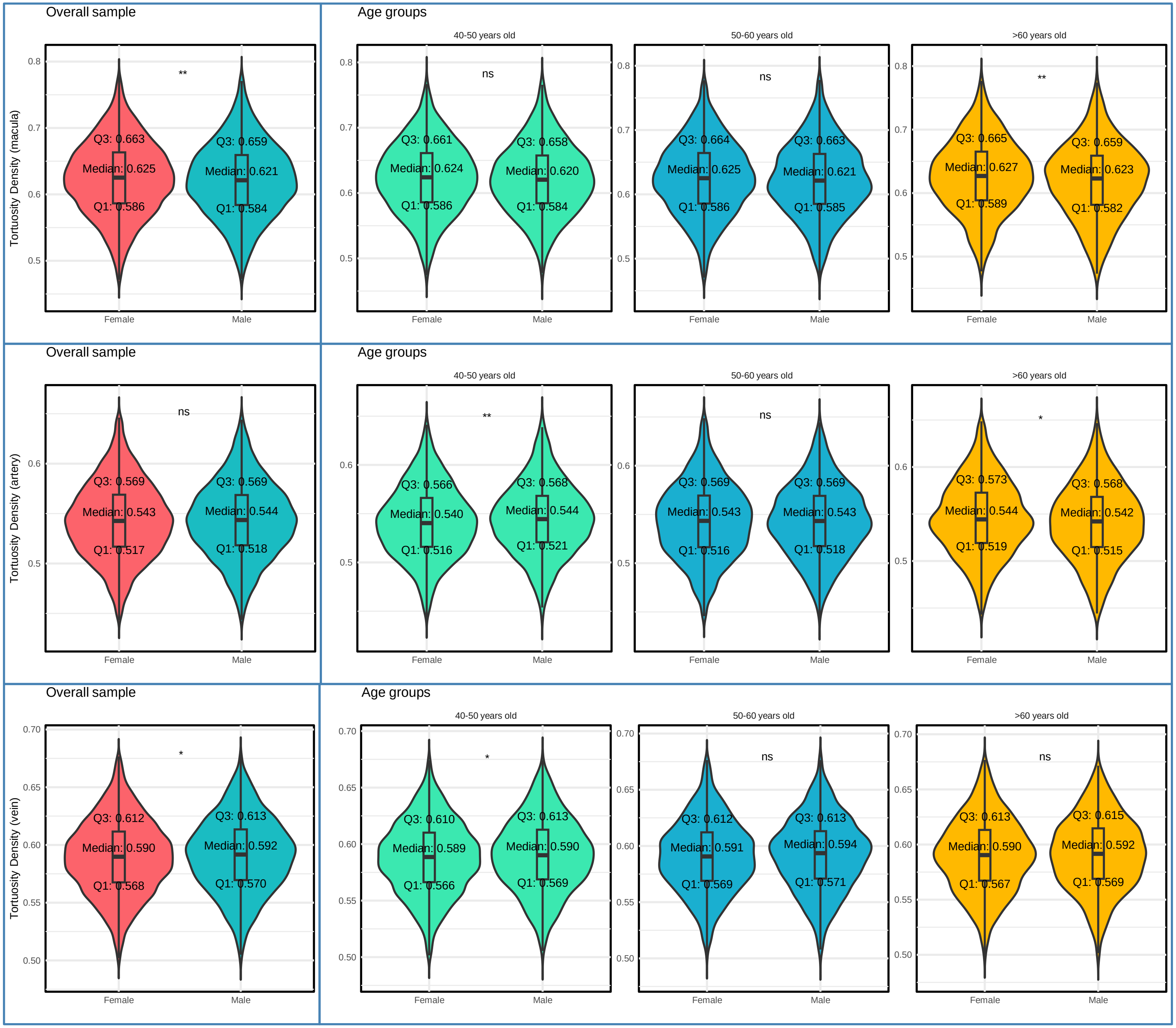
Overall sample
Age groups
40-50 years old
50-60 years old
>60 years old
Tortuosity Density (macula)
**
Q3: 0.663
Median: 0.625
Q1: 0.586
Q3: 0.659
Median: 0.621
Q1: 0.584
ns
Q3: 0.661
Median: 0.624
Q1: 0.586
Q3: 0.658
Median: 0.620
Q1: 0.584
ns
Q3: 0.664
Median: 0.625
Q1: 0.586
Q3: 0.663
Median: 0.621
Q1: 0.585
**
Q3: 0.665
Median: 0.627
Q1: 0.589
Q3: 0.659
Median: 0.623
Q1: 0.582
Tortuosity Density (artery)
ns
Q3: 0.569
Median: 0.543
Q1: 0.517
Q3: 0.569
Median: 0.544
Q1: 0.518
**
Q3: 0.566
Median: 0.540
Q1: 0.516
Q3: 0.568
Median: 0.544
Q1: 0.521
ns
Q3: 0.569
Median: 0.543
Q1: 0.516
Q3: 0.569
Median: 0.543
Q1: 0.518
*
Q3: 0.573
Median: 0.544
Q1: 0.519
Q3: 0.568
Median: 0.542
Q1: 0.515
Tortuosity Density (vein)
*
Q3: 0.612
Median: 0.590
Q1: 0.568
Q3: 0.613
Median: 0.592
Q1: 0.570
*
Q3: 0.610
Median: 0.589
Q1: 0.566
Q3: 0.613
Median: 0.590
Q1: 0.569
ns
Q3: 0.612
Median: 0.591
Q1: 0.569
Q3: 0.613
Median: 0.594
Q1: 0.571
ns
Q3: 0.613
Median: 0.590
Q1: 0.567
Q3: 0.615
Median: 0.592
Q1: 0.569
Female
Male

Supplementary Figure 6. Associations of Selected Retinal Vascular Measurements with Age and SBP

A. Associations of Selected Retinal Vascular Measurements with Age

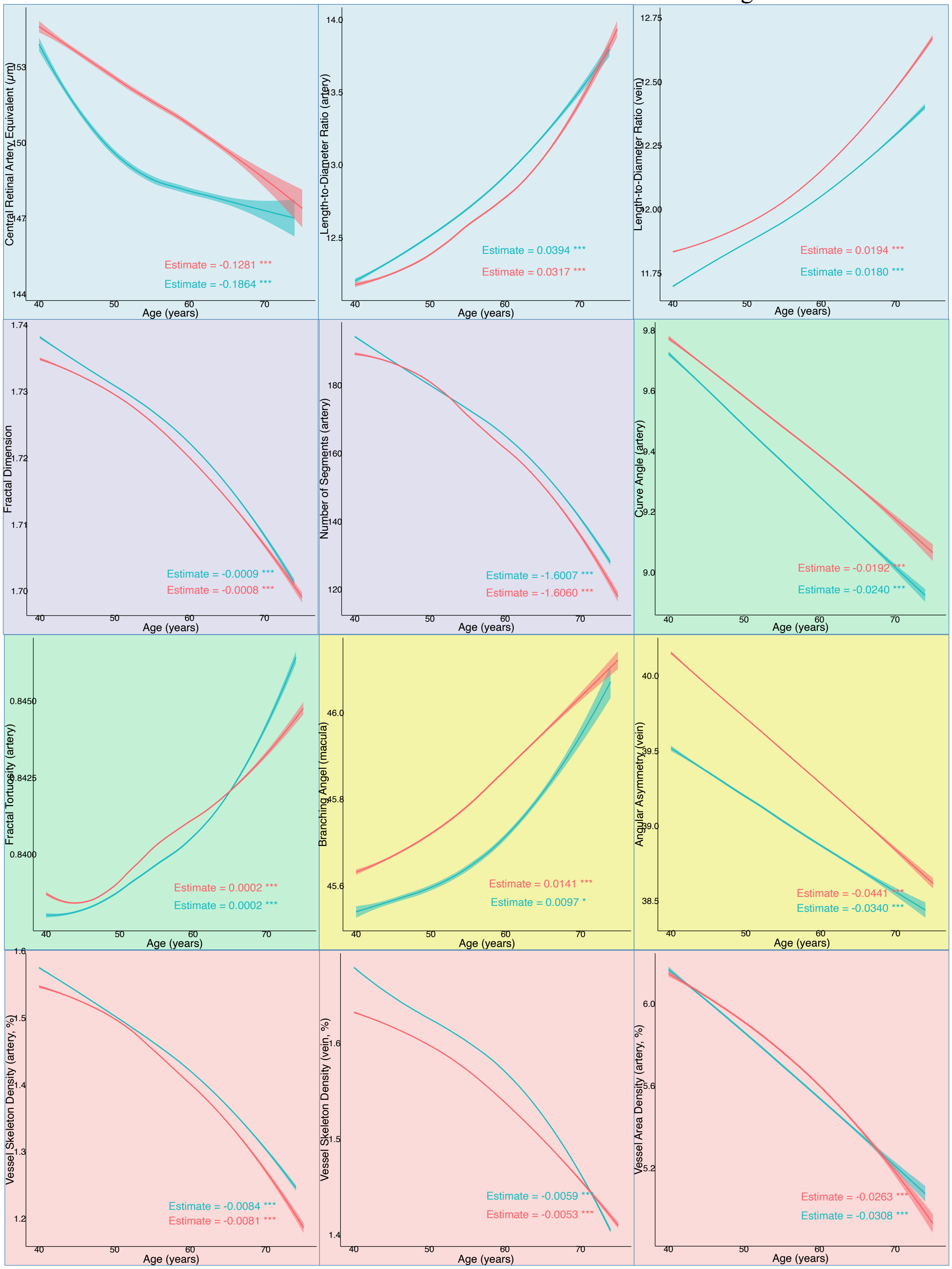

### B. Associations of Selected Retinal Vascular Measurements with SBP

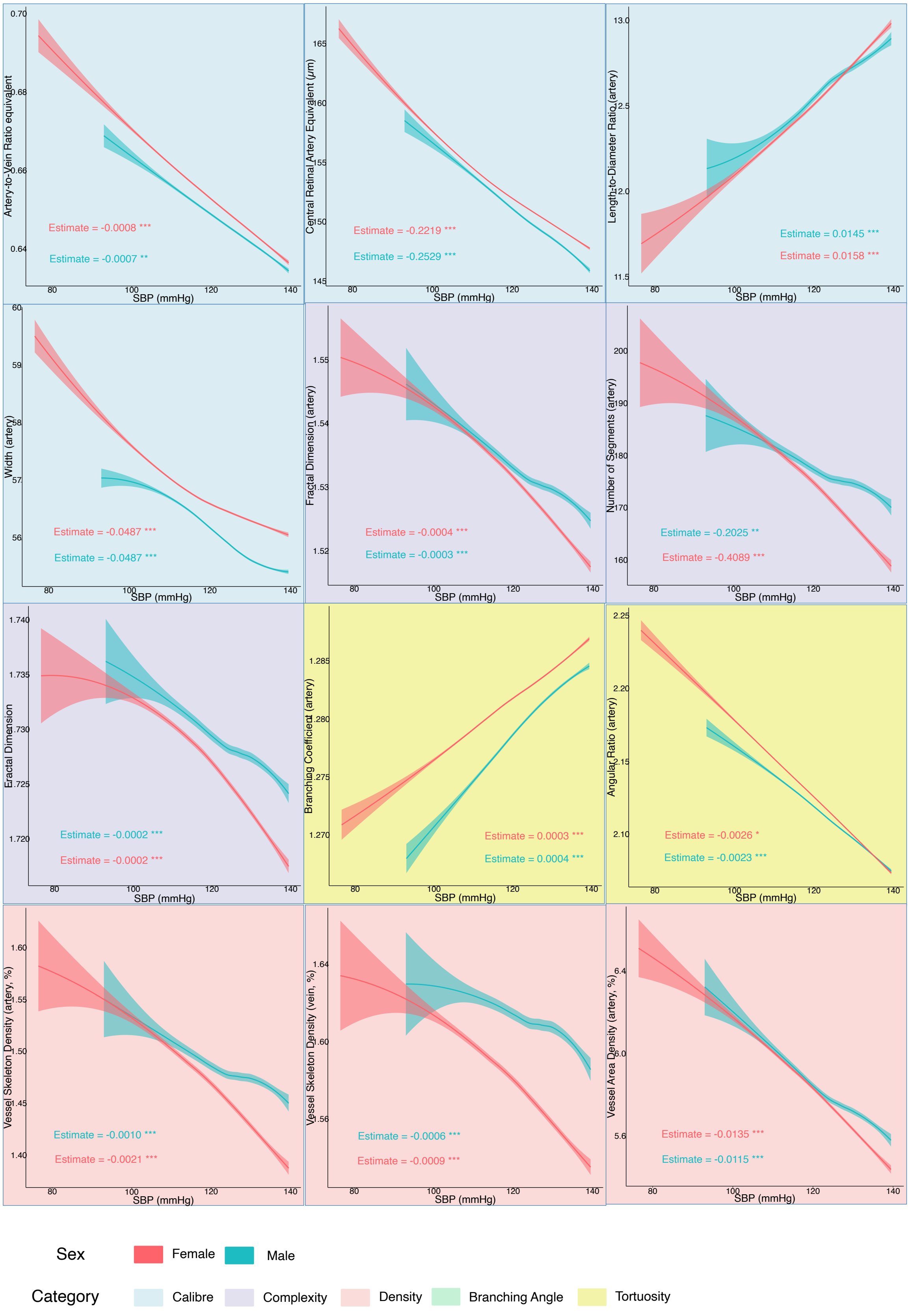

### Supplementary Figure 7. Correlations among retinal vascular measurements

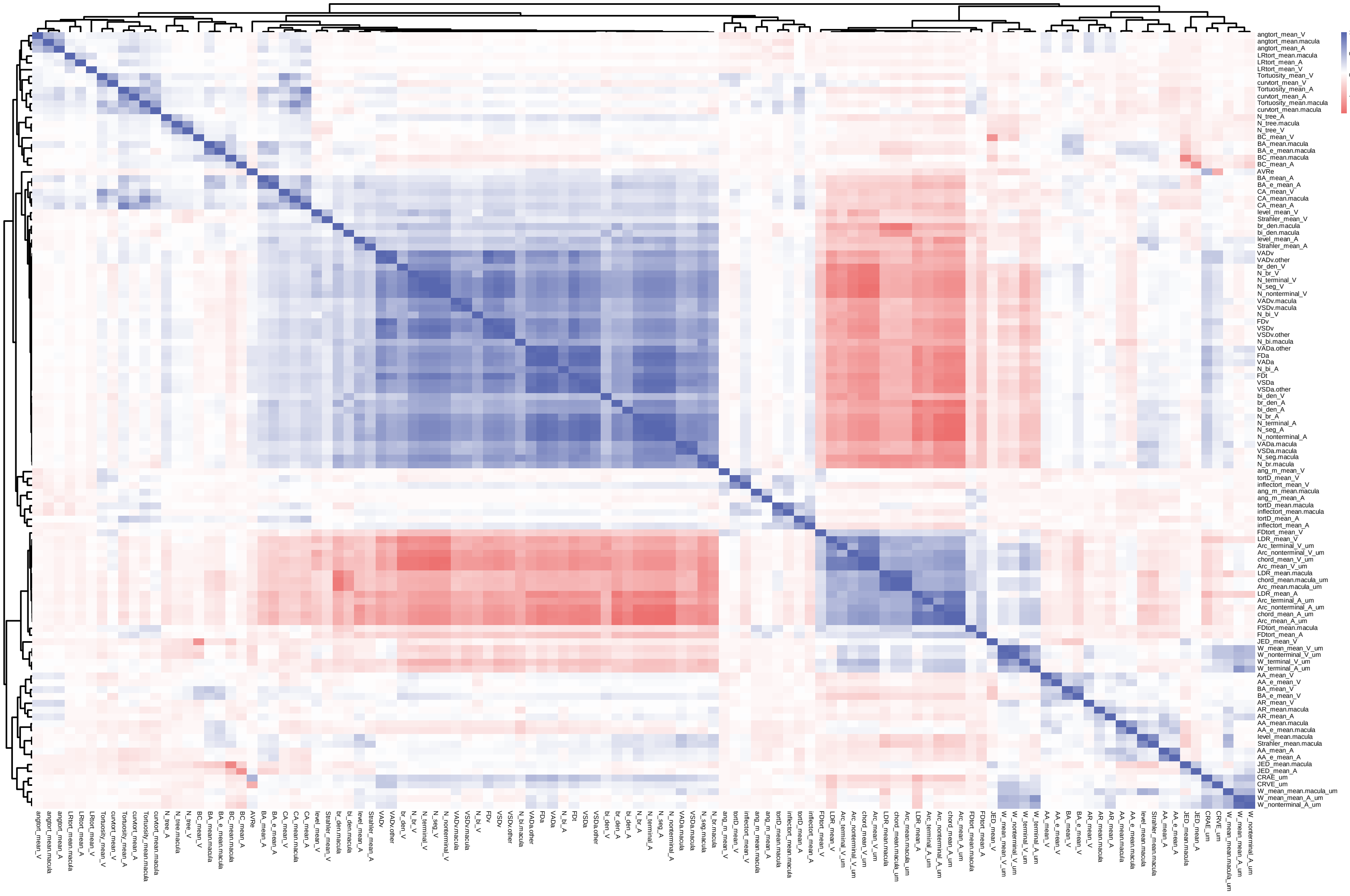

Supplementary Figure 8. Deviation of retinal vascular measurements in excluded participants

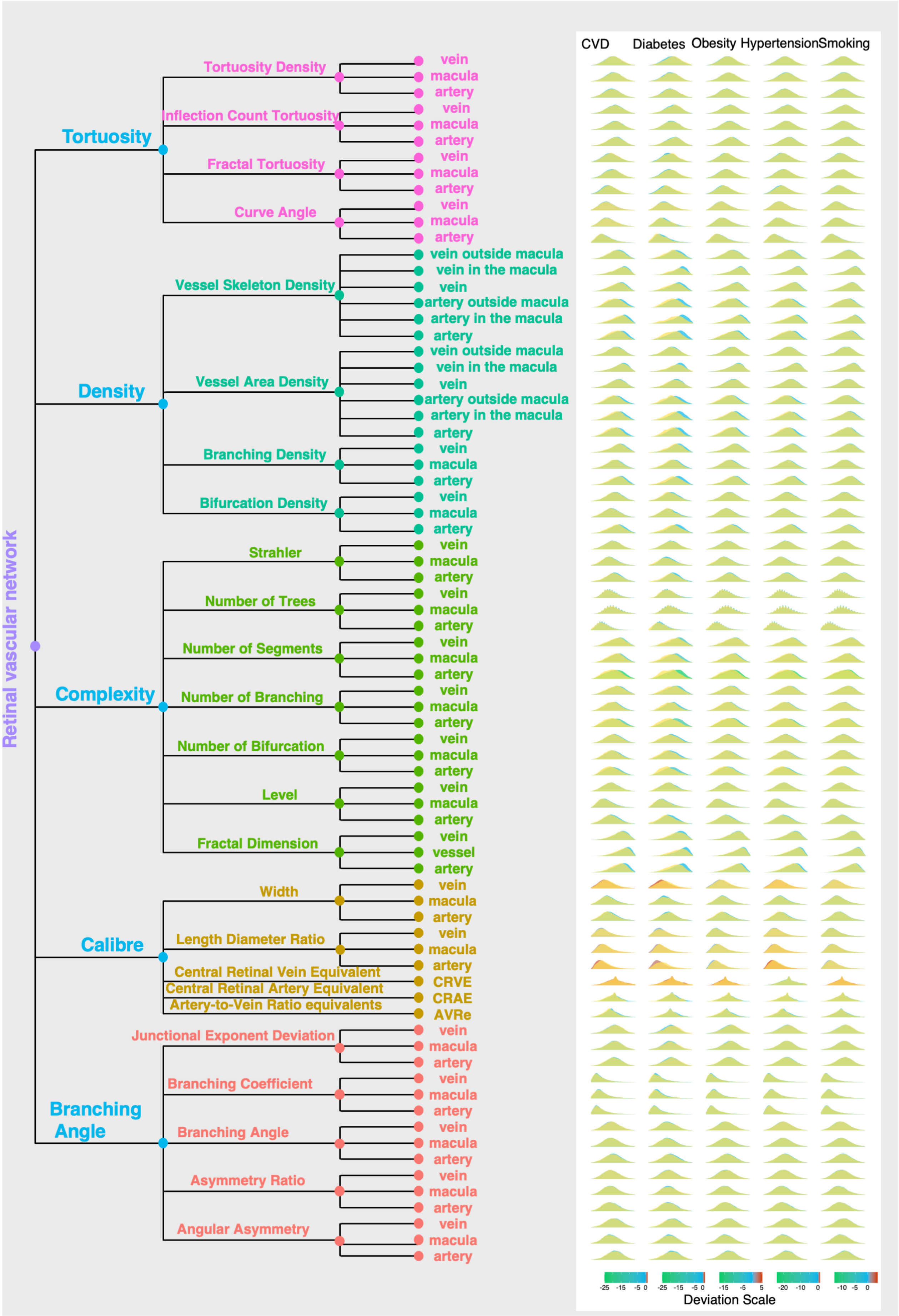

Supplementary Table 1. Mean (standard deviation) of retinal vascular measurements by age groups

| Retinal Vascular Measurements | Overall (n=10151) | 40-50 years (n=3888) | 50-60 years (n=3803) | >60 years (n=2460) | P value |
|---|---|---|---|---|---|
| **Calibre** | | | | | |
| **Artery-to-Vein Ratio equivalent** | 0.653 (0.068) | 0.656 (0.069) | 0.652 (0.068) | 0.647 (0.067) | <0.001 |
| **Central Retinal Artery Equivalent (µm)** | 152.022 (14.902) | 153.580 (14.919) | 151.698 (14.962) | 150.060 (14.520) | <0.001 |
| **Central Retinal Vein Equivalent (µm)** | 233.232 (21.527) | 234.394 (21.613) | 232.805 (21.412) | 232.055 (21.487) | 0.001 |
| **Length-to-Diameter Ratio** | | | | | |
| Vessels in macular region | 13.570 (1.861) | 13.288 (1.735) | 13.589 (1.841) | 13.987 (2.000) | <0.001 |
| Arteries | 12.839 (1.523) | 12.457 (1.349) | 12.866 (1.493) | 13.403 (1.642) | <0.001 |
| Veins | 12.154 (1.058) | 11.953 (0.950) | 12.154 (1.047) | 12.470 (1.155) | <0.001 |
| **Width (µm)** | | | | | |
| Vessels in macular region | 48.366 (2.923) | 48.258 (2.815) | 48.313 (2.954) | 48.621 (3.026) | <0.001 |
| Arteries | 56.598 (2.955) | 56.364 (2.811) | 56.525 (2.961) | 57.079 (3.109) | <0.001 |
| Veins | 63.254 (3.525) | 62.660 (3.271) | 63.214 (3.507) | 64.255 (3.716) | <0.001 |
| Terminal arteries | 39.750 (1.966) | 39.347 (1.753) | 39.719 (1.928) | 40.435 (2.151) | <0.001 |
| Terminal veins | 42.084 (2.242) | 41.608 (2.051) | 42.088 (2.232) | 42.832 (2.341) | <0.001 |
| Nonterminal arteries | 83.445 (5.322) | 82.661 (5.002) | 83.354 (5.264) | 84.825 (5.622) | <0.001 |
| Nonterminal veins | 72.428 (4.377) | 72.272 (4.235) | 72.313 (4.392) | 72.854 (4.545) | <0.001 |
| **Complexity** | | | | | |
| **Fractal Dimension** | | | | | |
| All vessels | 1.722 (0.032) | 1.732 (0.028) | 1.721 (0.031) | 1.706 (0.033) | <0.001 |
| Arteries | 1.526 (0.039) | 1.539 (0.035) | 1.525 (0.038) | 1.508 (0.039) | <0.001 |
| Veins | 1.563 (0.025) | 1.569 (0.023) | 1.562 (0.025) | 1.553 (0.026) | <0.001 |
| **Level (N)** | | | | | |
| Vessels in macular region | 3.164 (0.717) | 3.186 (0.695) | 3.159 (0.726) | 3.137 (0.736) | 0.004 |
| Arteries | 4.112 (0.570) | 4.198 (0.550) | 4.114 (0.565) | 3.972 (0.580) | <0.001 |
| Veins | 3.962 (0.455) | 4.018 (0.442) | 3.957 (0.454) | 3.879 (0.461) | <0.001 |
| **Strahler (N)** | | | | | |

| Retinal Vascular Measurements | Overall (n=10151) | 40-50 years (n=3888) | 50-60 years (n=3803) | >60 years (n=2460) | P value |
|---|---|---|---|---|---|
| Vessels in macular region | 1.662 (0.102) | 1.667 (0.099) | 1.661 (0.102) | 1.656 (0.106) | <0.001 |
| Arteries | 1.816 (0.074) | 1.827 (0.071) | 1.816 (0.074) | 1.800 (0.077) | <0.001 |
| Veins | 1.803 (0.064) | 1.809 (0.062) | 1.804 (0.064) | 1.793 (0.065) | <0.001 |
| **Number of segments (N)** | | | | | |
| Vessels in macular region | 85.457 (21.963) | 91.271 (20.627) | 85.306 (21.229) | 76.501 (22.093) | <0.001 |
| Arteries | 172.877 (45.228) | 189.666 (42.100) | 171.335 (43.014) | 148.726 (41.814) | <0.001 |
| Veins | 174.303 (33.598) | 185.200 (31.034) | 173.813 (32.395) | 157.838 (32.483) | <0.001 |
| **Number of trees (N)** | | | | | |
| Vessels in macular region | 11.188 (2.780) | 11.264 (2.786) | 11.123 (2.782) | 11.169 (2.767) | 0.095 |
| Arteries | 11.218 (3.035) | 11.534 (3.136) | 11.116 (2.964) | 10.876 (2.934) | <0.001 |
| Veins | 12.583 (3.037) | 12.703 (3.061) | 12.462 (3.017) | 12.580 (3.023) | 0.003 |
| **Number of branching** | | | | | |
| Vessels in macular region | 47.639 (14.234) | 50.889 (13.633) | 47.570 (13.868) | 42.609 (14.252) | <0.001 |
| Arteries | 97.886 (28.484) | 107.928 (26.695) | 96.968 (27.224) | 83.431 (26.584) | <0.001 |
| Veins | 100.043 (23.069) | 106.901 (21.675) | 99.608 (22.355) | 89.877 (22.433) | <0.001 |
| **Number of bifurcation (N)** | | | | | |
| Vessels in macular region | 26.725 (8.691) | 28.624 (8.562) | 26.600 (8.531) | 23.915 (8.353) | <0.001 |
| Arteries | 51.870 (15.048) | 56.713 (14.569) | 51.420 (14.483) | 44.914 (13.749) | <0.001 |
| Veins | 50.511 (11.251) | 53.271 (11.042) | 50.470 (10.995) | 46.213 (10.599) | <0.001 |
| **Number of Terminal point** | | | | | |
| Arteries | 83.447 (21.267) | 91.320 (19.756) | 82.707 (20.258) | 72.147 (19.720) | <0.001 |
| Veins | 84.971 (16.033) | 90.131 (14.932) | 84.720 (15.450) | 77.206 (15.395) | <0.001 |
| **Number of non-terminal point** | | | | | |
| Arteries | 89.381 (24.332) | 98.254 (22.727) | 88.596 (23.183) | 76.569 (22.511) | <0.001 |
| Veins | 89.279 (17.947) | 94.981 (16.501) | 89.048 (17.371) | 80.624 (17.495) | <0.001 |
| **Density** | | | | | |
| **Vessel Area Density (%)** | | | | | |
| Arteries | 5.747 (0.915) | 6.031 (0.863) | 5.713 (0.889) | 5.349 (0.878) | <0.001 |

| Retinal Vascular Measurements | Overall (n=10151) | 40-50 years (n=3888) | 50-60 years (n=3803) | >60 years (n=2460) | P value |
|---|---|---|---|---|---|
| Arteries in macular region | 5.297 (1.148) | 5.599 (1.047) | 5.276 (1.113) | 4.852 (1.203) | <0.001 |
| Arteries outside macular region | 5.847 (0.942) | 6.123 (0.909) | 5.809 (0.918) | 5.471 (0.888) | <0.001 |
| Veins | 7.216 (0.735) | 7.372 (0.698) | 7.202 (0.731) | 6.989 (0.737) | <0.001 |
| Veins in macular region | 4.733 (0.943) | 4.933 (0.855) | 4.737 (0.936) | 4.409 (0.997) | <0.001 |
| Veins outside macular region | 7.828 (0.776) | 7.964 (0.753) | 7.809 (0.773) | 7.642 (0.776) | <0.001 |
| **Vessel Skeleton Density (%)** | | | | | |
| Arteries | 1.453 (0.243) | 1.539 (0.223) | 1.445 (0.232) | 1.327 (0.231) | <0.001 |
| Arteries in macular region | 1.672 (0.339) | 1.770 (0.296) | 1.672 (0.325) | 1.520 (0.366) | <0.001 |
| Arteries outside macular region | 1.393 (0.237) | 1.476 (0.224) | 1.384 (0.228) | 1.276 (0.218) | <0.001 |
| Veins | 1.571 (0.178) | 1.625 (0.166) | 1.568 (0.173) | 1.489 (0.174) | <0.001 |
| Veins in macular region | 1.563 (0.299) | 1.639 (0.266) | 1.565 (0.293) | 1.440 (0.315) | <0.001 |
| Veins outside macular region | 1.570 (0.166) | 1.617 (0.158) | 1.566 (0.161) | 1.500 (0.157) | <0.001 |
| **Branching Density (%)** | | | | | |
| Vessels in macular region | 1.461 (0.218) | 1.494 (0.209) | 1.459 (0.216) | 1.410 (0.225) | <0.001 |
| Arteries | 1.389 (0.172) | 1.438 (0.156) | 1.385 (0.169) | 1.318 (0.178) | <0.001 |
| Veins | 1.344 (0.142) | 1.378 (0.128) | 1.342 (0.140) | 1.291 (0.149) | <0.001 |
| **Bifurcation Density (%)** | | | | | |
| Vessels in macular region | 1.433 (0.244) | 1.463 (0.239) | 1.429 (0.241) | 1.393 (0.250) | <0.001 |
| Arteries | 1.331 (0.204) | 1.383 (0.188) | 1.330 (0.200) | 1.251 (0.207) | <0.001 |
| Veins | 1.291 (0.159) | 1.323 (0.153) | 1.290 (0.157) | 1.244 (0.161) | <0.001 |
| **Chord Length (μm)** | | | | | |
| Vessels in macular region | 538.461 (73.699) | 525.107 (66.748) | 538.809 (72.157) | 559.026 (81.375) | <0.001 |
| Arteries | 583.966 (82.342) | 559.264 (69.269) | 585.034 (80.354) | 621.356 (89.744) | <0.001 |
| Veins | 595.689 (63.133) | 578.780 (53.989) | 596.049 (61.894) | 621.857 (69.167) | <0.001 |
| **Arc Length (μm)** | | | | | |
| Vessels in macular region | 589.742 (81.484) | 574.878 (74.186) | 590.193 (79.595) | 612.538 (89.676) | <0.001 |
| Arteries | 633.910 (89.126) | 607.080 (74.785) | 634.996 (87.098) | 674.633 (97.053) | <0.001 |

| Retinal Vascular Measurements | Overall (n=10151) | 40-50 years (n=3888) | 50-60 years (n=3803) | >60 years (n=2460) | P value |
|---|---|---|---|---|---|
| Veins | 646.410 (68.883) | 627.823 (58.947) | 646.813 (67.360) | 675.165 (75.522) | <0.001 |
| Nonterminal arteries | 668.744 (107.807) | 636.044 (89.917) | 670.318 (104.814) | 717.991 (118.686) | <0.001 |
| Nonterminal veins | 687.284 (82.002) | 664.948 (70.595) | 688.317 (80.117) | 720.989 (89.592) | <0.001 |
| Terminal arteries | 596.283 (81.650) | 576.463 (71.772) | 597.227 (80.590) | 626.148 (88.425) | <0.001 |
| Terminal veins | 603.737 (70.781) | 589.500 (63.106) | 603.400 (70.231) | 626.758 (76.859) | <0.001 |
| **Angular Asymmetry (Degree)** | | | | | |
| Vessels in macular region | 35.596 (4.588) | 35.655 (4.437) | 35.550 (4.571) | 35.572 (4.840) | 0.521 |
| Arteries | 41.132 (3.736) | 41.448 (3.581) | 41.134 (3.753) | 40.631 (3.892) | <0.001 |
| Veins | 39.397 (3.434) | 39.733 (3.365) | 39.346 (3.422) | 38.947 (3.504) | <0.001 |
| **Junctional Exponent Deviation** | | | | | |
| Vessels in macular region | 0.049 (0.107) | 0.054 (0.103) | 0.046 (0.107) | 0.047 (0.112) | 0.003 |
| Arteries | -0.095 (0.076) | -0.095 (0.074) | -0.096 (0.076) | -0.093 (0.080) | 0.396 |
| Veins | -0.007 (0.070) | -0.010 (0.068) | -0.007 (0.069) | -0.003 (0.074) | <0.001 |
| **Angular Asymmetry at edge (Degree)** | | | | | |
| Vessels in macular region | 39.853 (5.192) | 39.681 (4.853) | 39.790 (5.128) | 40.221 (5.762) | 0.013 |
| Arteries | 42.874 (3.974) | 43.091 (3.786) | 42.857 (3.940) | 42.560 (4.284) | <0.001 |
| Veins | 43.259 (3.543) | 43.443 (3.390) | 43.218 (3.533) | 43.034 (3.772) | <0.001 |
| **Asymmetry Ratio** | | | | | |
| Vessels in macular region | 1.773 (0.317) | 1.767 (0.303) | 1.776 (0.315) | 1.776 (0.340) | 0.45 |
| Arteries | 2.133 (0.334) | 2.142 (0.325) | 2.129 (0.337) | 2.126 (0.345) | 0.022 |
| Veins | 2.486 (0.434) | 2.479 (0.423) | 2.494 (0.431) | 2.485 (0.455) | 0.367 |
| **Branching Angle (Degree)** | | | | | |
| Vessels in macular region | 45.809 (2.206) | 45.725 (2.173) | 45.785 (2.177) | 45.979 (2.290) | <0.001 |
| Arteries | 47.133 (1.733) | 47.246 (1.681) | 47.108 (1.745) | 46.996 (1.782) | <0.001 |
| Veins | 46.665 (1.355) | 46.606 (1.331) | 46.676 (1.350) | 46.742 (1.398) | <0.001 |
| **Branching Angle at edge (Degree)** | | | | | |
| Vessels in macular region | 41.974 (3.046) | 41.935 (2.899) | 41.897 (2.951) | 42.154 (3.391) | 0.046 |
| Arteries | 43.546 (2.466) | 43.775 (2.351) | 43.504 (2.471) | 43.249 (2.598) | <0.001 |

| Retinal Vascular Measurements | Overall (n=10151) | 40-50 years (n=3888) | 50-60 years (n=3803) | >60 years (n=2460) | P value |
|---|---|---|---|---|---|
| Veins | 42.512 (1.978) | 42.605 (1.872) | 42.495 (1.974) | 42.392 (2.134) | <0.001 |
| **Branching Coefficient** | | | | | |
| Vessels in macular region | 1.209 (0.092) | 1.204 (0.088) | 1.210 (0.091) | 1.215 (0.098) | <0.001 |
| Arteries | 1.289 (0.094) | 1.288 (0.094) | 1.289 (0.094) | 1.289 (0.095) | 0.721 |
| Veins | 1.224 (0.089) | 1.225 (0.088) | 1.223 (0.088) | 1.225 (0.092) | 0.54 |
| **Tortuosity** | | | | | |
| **Tortuosity Density** | | | | | |
| Vessels in macular region | 0.624 (0.055) | 0.623 (0.054) | 0.625 (0.056) | 0.625 (0.057) | 0.219 |
| Arteries | 0.543 (0.037) | 0.543 (0.036) | 0.544 (0.038) | 0.544 (0.039) | 0.317 |
| Veins | 0.590 (0.032) | 0.589 (0.032) | 0.591 (0.032) | 0.591 (0.033) | 0.002 |
| **Curve Angle (Degree)** | | | | | |
| Vessels in macular region | 11.940 (1.610) | 11.959 (1.581) | 11.934 (1.598) | 11.918 (1.672) | 0.002 |
| Arteries | 9.705 (1.487) | 9.881 (1.498) | 9.656 (1.459) | 9.503 (1.482) | <0.001 |
| Veins | 10.134 (0.992) | 10.155 (0.957) | 10.151 (0.993) | 10.075 (1.041) | <0.001 |
| **Fractal Tortuosity** | | | | | |
| Vessels in macular region | 0.835 (0.012) | 0.833 (0.012) | 0.835 (0.012) | 0.836 (0.012) | <0.001 |
| Arteries | 0.840 (0.009) | 0.839 (0.008) | 0.840 (0.009) | 0.842 (0.009) | <0.001 |
| Veins | 0.838 (0.008) | 0.837 (0.008) | 0.838 (0.008) | 0.840 (0.009) | <0.001 |
| **Inflection Count Tortuosity** | | | | | |
| Vessels in macular region | 0.260 (0.026) | 0.261 (0.025) | 0.260 (0.026) | 0.258 (0.028) | 0.507 |
| Arteries | 0.226 (0.017) | 0.227 (0.017) | 0.226 (0.017) | 0.225 (0.018) | <0.001 |
| Veins | 0.245 (0.016) | 0.246 (0.016) | 0.245 (0.016) | 0.243 (0.017) | <0.001 |
| **Distance-based Tortuosity** | | | | | |
| Vessels in macular region | 1.084 (0.008) | 1.085 (0.009) | 1.084 (0.008) | 1.084 (0.009) | 0.002 |
| Arteries | 1.094 (0.009) | 1.093 (0.008) | 1.094 (0.009) | 1.094 (0.009) | 0.002 |
| Veins | 1.084 (0.005) | 1.084 (0.005) | 1.084 (0.005) | 1.084 (0.005) | 0.014 |
| **Linear Regression Tortuosity** | | | | | |
| Vessels in macular region | 3.742 (2.629) | 3.689 (2.572) | 3.726 (2.632) | 3.849 (2.711) | 0.008 |

| Retinal Vascular Measurements | Overall (n=10151) | 40-50 years (n=3888) | 50-60 years (n=3803) | >60 years (n=2460) | P value |
|---|---|---|---|---|---|
| Arteries | 3.992 (2.695) | 3.968 (2.609) | 3.973 (2.712) | 4.059 (2.800) | 0.445 |
| Veins | 3.256 (1.722) | 3.233 (1.699) | 3.241 (1.719) | 3.316 (1.760) | 0.026 |
| **Angle-based Tortuosity** | | | | | |
| Vessels in macular region | 0.075 (0.047) | 0.073 (0.046) | 0.075 (0.047) | 0.077 (0.050) | 0.169 |
| Arteries | 0.073 (0.036) | 0.073 (0.036) | 0.073 (0.036) | 0.073 (0.038) | 0.489 |
| Veins | 0.065 (0.031) | 0.064 (0.030) | 0.065 (0.031) | 0.066 (0.032) | 0.204 |
| **Squared Curvature Tortuosity** | | | | | |
| Vessels in macular region | 0.129 (0.016) | 0.128 (0.016) | 0.129 (0.016) | 0.130 (0.016) | 0.011 |
| Arteries | 0.117 (0.012) | 0.116 (0.012) | 0.117 (0.012) | 0.117 (0.012) | 0.609 |
| Veins | 0.116 (0.010) | 0.116 (0.009) | 0.116 (0.010) | 0.117 (0.010) | <0.001 |
| **Turning Angles** | | | | | |
| Vessels in macular region | 0.368 (0.017) | 0.368 (0.017) | 0.368 (0.017) | 0.368 (0.017) | 0.984 |
| Arteries | 0.376 (0.013) | 0.376 (0.012) | 0.376 (0.013) | 0.376 (0.014) | 0.348 |
| Veins | 0.370 (0.012) | 0.370 (0.012) | 0.370 (0.012) | 0.370 (0.012) | 0.661 |

Notes: Kolmogorov-Smirnov was performed to test distribution normality. For normally distributed measurements, the Analysis of Variance was performed to test the intergroup difference, otherwise, the Kruskal-Wallis H test was conducted. P value <0.05 was set for the significance of intergroup differences.

Supplementary Table 2. Median (1st quartile to 3rd quartile) of retinal vascular measurements by age groups

| **Retinal Vascular Measurements** | **Overall** (n=10151) | **40-50 years** (n=3888) | **50-60 years** (n=3803) | **>60 years** (n=2460) |
|---|---|---|---|---|
| **Calibre** | | | | |
| **Artery-to-Vein Ratio equivalent** | 0.647 [0.609, 0.695] | 0.651 [0.611, 0.700] | 0.647 [0.610, 0.694] | 0.641 [0.606, 0.687] |
| **Central Retinal Artery Equivalent (μm)** | 151.181 [141.992, 161.318] | 152.491 [143.585, 162.979] | 151.003 [141.667, 161.178] | 150.004 [140.289, 158.919] |
| **Central Retinal Vein Equivalent (μm)** | 233.703 [218.691, 247.217] | 234.203 [219.376, 248.712] | 233.433 [218.335, 246.719] | 233.467 [217.941, 245.971] |
| **Length-to-Diameter Ratio** | | | | |
| Vessels in macular region | 13.277 [12.205, 14.626] | 13.019 [12.053, 14.230] | 13.317 [12.257, 14.590] | 13.655 [12.443, 15.209] |
| Arteries | 12.549 [11.730, 13.628] | 12.173 [11.503, 13.092] | 12.609 [11.793, 13.629] | 13.128 [12.185, 14.389] |
| Veins | 11.970 [11.386, 12.718] | 11.810 [11.271, 12.431] | 11.981 [11.410, 12.709] | 12.293 [11.615, 13.209] |
| **Width (μm)** | | | | |
| Vessels in macular region | 48.038 [46.302, 50.142] | 47.946 [46.273, 49.978] | 48.013 [46.223, 50.075] | 48.288 [46.481, 50.557] |
| Arteries | 56.376 [54.526, 58.428] | 56.150 [54.385, 58.083] | 56.313 [54.502, 58.335] | 56.861 [54.849, 59.103] |
| Veins | 62.815 [60.759, 65.266] | 62.281 [60.327, 64.456] | 62.758 [60.747, 65.286] | 63.886 [61.666, 66.610] |
| Terminal arteries | 39.426 [38.322, 40.822] | 39.110 [38.111, 40.310] | 39.407 [38.308, 40.764] | 40.123 [38.839, 41.700] |
| Terminal veins | 41.775 [40.456, 43.372] | 41.325 [40.140, 42.707] | 41.781 [40.494, 43.381] | 42.588 [41.085, 44.287] |
| Nonterminal arteries | 72.222 [69.391, 75.277] | 72.063 [69.393, 74.996] | 72.164 [69.313, 75.158] | 72.703 [69.568, 75.966] |
| Nonterminal veins | 82.943 [79.652, 86.690] | 82.157 [79.094, 85.620] | 82.880 [79.599, 86.571] | 84.337 [80.890, 88.404] |
| **Complexity** | | | | |
| **Fractal Dimension** | | | | |
| All vessels | 1.729 [1.704, 1.746] | 1.739 [1.718, 1.752] | 1.728 [1.705, 1.744] | 1.711 [1.685, 1.732] |
| Arteries | 1.532 [1.503, 1.555] | 1.546 [1.519, 1.564] | 1.531 [1.503, 1.553] | 1.512 [1.484, 1.537] |
| Veins | 1.566 [1.548, 1.581] | 1.573 [1.556, 1.586] | 1.566 [1.548, 1.580] | 1.556 [1.537, 1.571] |
| **Level (N)** | | | | |
| Vessels in macular region | 3.062 [2.634, 3.602] | 3.093 [2.663, 3.622] | 3.043 [2.621, 3.586] | 3.037 [2.592, 3.591] |
| Arteries | 4.096 [3.715, 4.493] | 4.187 [3.821, 4.576] | 4.100 [3.728, 4.477] | 3.952 [3.544, 4.360] |
| Veins | 3.949 [3.645, 4.265] | 4.006 [3.717, 4.312] | 3.941 [3.643, 4.263] | 3.871 [3.530, 4.183] |
| **Strahler (N)** | | | | |

| | | | | |
|---|---|---|---|---|
| Vessels in macular region | 1.653 [1.589, 1.727] | 1.657 [1.597, 1.730] | 1.652 [1.587, 1.727] | 1.648 [1.581, 1.720] |
| Arteries | 1.816 [1.766, 1.866] | 1.825 [1.779, 1.874] | 1.815 [1.767, 1.864] | 1.798 [1.744, 1.852] |
| Veins | 1.803 [1.760, 1.847] | 1.810 [1.767, 1.851] | 1.803 [1.761, 1.848] | 1.792 [1.747, 1.837] |
| **Number of segments (N)** | | | | |
| Vessels in macular region | 87.000 [71.000, 101.000] | 93.000 [78.000, 106.000] | 87.000 [72.000, 100.000] | 77.000 [61.000, 93.000] |
| Arteries | 177.000 [141.000, 207.000] | 197.000 [164.000, 219.250] | 175.000 [142.000, 203.000] | 149.500 [118.000, 180.000] |
| Veins | 178.000 [153.000, 199.000] | 189.000 [167.000, 208.000] | 177.000 [154.000, 197.000] | 160.000 [136.000, 182.000] |
| **Number of trees (N)** | | | | |
| Vessels in macular region | 11.000 [9.000, 13.000] | 11.000 [9.000, 13.000] | 11.000 [9.000, 13.000] | 11.000 [9.000, 13.000] |
| Arteries | 11.000 [9.000, 13.000] | 11.000 [9.000, 13.000] | 10.710 [9.000, 13.000] | 10.000 [9.000, 12.000] |
| Veins | 12.000 [10.000, 14.000] | 12.000 [10.160, 15.000] | 12.000 [10.000, 14.000] | 12.000 [10.000, 14.000] |
| **Number of branching** | | | | |
| Vessels in macular region | 48.000 [38.000, 58.000] | 51.000 [42.000, 60.000] | 48.000 [38.000, 57.000] | 42.000 [32.000, 52.000] |
| Arteries | 100.000 [78.000, 119.000] | 111.000 [91.000, 127.000] | 98.000 [79.000, 117.000] | 83.000 [64.000, 103.000] |
| Veins | 102.000 [85.000, 117.000] | 109.000 [93.000, 122.000] | 101.000 [85.000, 116.000] | 91.000 [74.000, 106.000] |
| **Number of bifurcation (N)** | | | | |
| Vessels in macular region | 27.000 [21.000, 33.000] | 29.000 [23.000, 35.000] | 26.000 [21.000, 33.000] | 24.000 [18.000, 30.000] |
| Arteries | 52.000 [41.000, 62.000] | 57.000 [47.000, 67.000] | 52.000 [41.000, 61.000] | 45.000 [35.000, 54.000] |
| Veins | 50.000 [43.000, 58.000] | 53.500 [46.000, 61.000] | 50.000 [43.000, 58.000] | 46.000 [39.000, 53.000] |
| **Number of Terminal point** | | | | |
| Arteries | 86.000 [68.000, 100.000] | 95.000 [79.000, 106.000] | 85.000 [69.000, 98.000] | 72.000 [58.000, 87.000] |
| Veins | 87.000 [75.000, 97.000] | 92.000 [81.000, 101.000] | 86.000 [75.000, 96.000] | 78.000 [67.000, 88.000] |
| **Number of non-terminal point** | | | | |
| Arteries | 91.000 [72.000, 107.000] | 102.000 [84.000, 115.000] | 90.000 [73.000, 105.000] | 77.000 [60.000, 93.000] |
| Veins | 91.000 [78.000, 102.000] | 97.000 [85.000, 107.000] | 91.000 [78.000, 102.000] | 82.000 [69.000, 93.250] |
| **Density** | | | | |
| **Vessel Area Density (%)** | | | | |
| Arteries | 5.813 [5.127, 6.420] | 6.150 [5.487, 6.653] | 5.771 [5.129, 6.360] | 5.381 [4.757, 5.989] |
| Arteries in macular region | 5.416 [4.583, 6.135] | 5.697 [4.990, 6.332] | 5.372 [4.585, 6.097] | 4.935 [4.034, 5.740] |
| Arteries outside macular region | 5.895 [5.208, 6.530] | 6.191 [5.532, 6.773] | 5.854 [5.194, 6.463] | 5.482 [4.901, 6.109] |

| | | | | |
|---|---|---|---|---|
| Veins | 7.276 [6.749, 7.744] | 7.441 [6.945, 7.877] | 7.252 [6.733, 7.737] | 7.039 [6.524, 7.495] |
| Veins in macular region | 4.893 [4.176, 5.424] | 5.080 [4.479, 5.553] | 4.900 [4.173, 5.415] | 4.523 [3.787, 5.134] |
| Veins outside macular region | 7.866 [7.325, 8.372] | 8.009 [7.490, 8.503] | 7.846 [7.294, 8.352] | 7.664 [7.135, 8.183] |
| **Vessel Skeleton Density (%)** | | | | |
| Arteries | 1.482 [1.290, 1.636] | 1.580 [1.403, 1.702] | 1.472 [1.300, 1.617] | 1.338 [1.165, 1.502] |
| Arteries in macular region | 1.736 [1.478, 1.920] | 1.824 [1.627, 1.984] | 1.730 [1.491, 1.904] | 1.565 [1.278, 1.786] |
| Arteries outside macular region | 1.413 [1.227, 1.571] | 1.506 [1.330, 1.639] | 1.403 [1.230, 1.555] | 1.274 [1.124, 1.437] |
| Veins | 1.594 [1.461, 1.702] | 1.649 [1.531, 1.747] | 1.590 [1.463, 1.694] | 1.507 [1.374, 1.616] |
| Veins in macular region | 1.621 [1.396, 1.783] | 1.690 [1.506, 1.833] | 1.624 [1.397, 1.780] | 1.483 [1.232, 1.669] |
| Veins outside macular region | 1.581 [1.462, 1.690] | 1.633 [1.522, 1.734] | 1.577 [1.462, 1.682] | 1.508 [1.396, 1.607] |
| **Branching Density (%)** | | | | |
| Vessels in macular region | 1.467 [1.317, 1.614] | 1.502 [1.354, 1.638] | 1.462 [1.317, 1.614] | 1.420 [1.247, 1.566] |
| Arteries | 1.407 [1.282, 1.513] | 1.454 [1.349, 1.548] | 1.400 [1.282, 1.506] | 1.330 [1.204, 1.448] |
| Veins | 1.359 [1.258, 1.445] | 1.392 [1.305, 1.469] | 1.358 [1.257, 1.441] | 1.305 [1.192, 1.395] |
| **Bifurcation Density (%)** | | | | |
| Vessels in macular region | 1.425 [1.264, 1.595] | 1.458 [1.299, 1.620] | 1.421 [1.261, 1.584] | 1.383 [1.212, 1.559] |
| Arteries | 1.350 [1.202, 1.477] | 1.400 [1.269, 1.516] | 1.346 [1.206, 1.471] | 1.256 [1.110, 1.397] |
| Veins | 1.294 [1.185, 1.401] | 1.321 [1.223, 1.429] | 1.296 [1.184, 1.395] | 1.238 [1.131, 1.355] |
| **Chord Length (µm)** | | | | |
| Vessels in macular region | 526.026 [484.851, 578.556] | 513.462 [476.809, 560.880] | 528.260 [486.763, 577.661] | 544.874 [498.169, 607.928] |
| Arteries | 564.722 [525.068, 622.040] | 542.064 [512.340, 587.903] | 567.443 [528.017, 621.097] | 603.303 [555.420, 670.831] |
| Veins | 583.348 [550.548, 626.176] | 568.737 [541.747, 603.654] | 584.407 [552.086, 625.551] | 608.536 [572.273, 661.219] |
| **Arc Length (µm)** | | | | |
| Vessels in macular region | 526.026 [484.851, 578.556] | 513.462 [476.809, 560.880] | 528.260 [486.763, 577.661] | 544.874 [498.169, 607.928] |
| Arteries | 564.722 [525.068, 622.040] | 542.064 [512.340, 587.903] | 567.443 [528.017, 621.097] | 603.303 [555.420, 670.831] |
| Veins | 583.348 [550.548, 626.176] | 568.737 [541.747, 603.654] | 584.407 [552.086, 625.551] | 608.536 [572.273, 661.219] |
| Nonterminal arteries | 526.026 [484.851, 578.556] | 513.462 [476.809, 560.880] | 528.260 [486.763, 577.661] | 544.874 [498.169, 607.928] |
| Nonterminal veins | 564.722 [525.068, 622.040] | 542.064 [512.340, 587.903] | 567.443 [528.017, 621.097] | 603.303 [555.420, 670.831] |
| Terminal arteries | 583.348 [550.548, 626.176] | 568.737 [541.747, 603.654] | 584.407 [552.086, 625.551] | 608.536 [572.273, 661.219] |
| Terminal veins | 526.026 [484.851, 578.556] | 513.462 [476.809, 560.880] | 528.260 [486.763, 577.661] | 544.874 [498.169, 607.928] |

| | | | | |
|---|---|---|---|---|
| **Angular Asymmetry (Degree)** | | | | |
| Vessels in macular region | 35.381 [32.381, 38.620] | 35.401 [32.550, 38.506] | 35.327 [32.346, 38.572] | 35.414 [32.029, 38.904] |
| Arteries | 41.155 [38.638, 43.708] | 41.481 [39.133, 43.920] | 41.166 [38.599, 43.724] | 40.714 [37.938, 43.284] |
| Veins | 39.443 [37.037, 41.783] | 39.758 [37.410, 42.088] | 39.430 [36.985, 41.704] | 38.893 [36.615, 41.355] |
| **Junctional Exponent Deviation** | | | | |
| Vessels in macular region | 0.051 [-0.021, 0.123] | 0.057 [-0.014, 0.125] | 0.047 [-0.022, 0.118] | 0.047 [-0.029, 0.125] |
| Arteries | -0.094 [-0.146, -0.042] | -0.093 [-0.145, -0.044] | -0.095 [-0.145, -0.045] | -0.092 [-0.148, -0.038] |
| Veins | -0.004 [-0.054, 0.041] | -0.007 [-0.056, 0.036] | -0.005 [-0.054, 0.041] | 0.000 [-0.053, 0.049] |
| **Angular Asymmetry at edge (Degree)** | | | | |
| Vessels in macular region | 39.683 [36.336, 43.034] | 39.522 [36.373, 42.727] | 39.666 [36.364, 42.889] | 39.915 [36.248, 43.822] |
| Arteries | 42.872 [40.291, 45.485] | 43.010 [40.616, 45.626] | 42.823 [40.265, 45.483] | 42.638 [39.791, 45.255] |
| Veins | 43.223 [40.924, 45.580] | 43.365 [41.153, 45.713] | 43.230 [40.845, 45.467] | 43.024 [40.540, 45.566] |
| **Asymmetry Ratio** | | | | |
| Vessels in macular region | 1.755 [1.550, 1.972] | 1.754 [1.551, 1.958] | 1.761 [1.559, 1.977] | 1.747 [1.533, 1.996] |
| Arteries | 2.117 [1.895, 2.357] | 2.129 [1.913, 2.360] | 2.117 [1.888, 2.357] | 2.098 [1.876, 2.355] |
| Veins | 2.473 [2.176, 2.778] | 2.473 [2.175, 2.762] | 2.480 [2.186, 2.783] | 2.462 [2.164, 2.788] |
| **Branching Angle (Degree)** | | | | |
| Vessels in macular region | 45.730 [44.281, 47.286] | 45.644 [44.212, 47.162] | 45.705 [44.306, 47.213] | 45.903 [44.362, 47.490] |
| Arteries | 47.129 [45.946, 48.330] | 47.211 [46.101, 48.423] | 47.128 [45.902, 48.296] | 46.941 [45.779, 48.222] |
| Veins | 46.592 [45.714, 47.567] | 46.539 [45.669, 47.475] | 46.592 [45.724, 47.592] | 46.686 [45.766, 47.655] |
| **Branching Angle at edge (Degree)** | | | | |
| Vessels in macular region | 41.898 [39.972, 43.776] | 41.886 [40.077, 43.726] | 41.851 [39.889, 43.656] | 42.037 [39.934, 44.082] |
| Arteries | 43.579 [41.946, 45.171] | 43.770 [42.252, 45.289] | 43.533 [41.907, 45.134] | 43.305 [41.528, 45.027] |
| Veins | 42.493 [41.228, 43.743] | 42.588 [41.373, 43.783] | 42.467 [41.166, 43.771] | 42.389 [41.080, 43.648] |
| **Branching Coefficient** | | | | |
| Vessels in macular region | 1.186 [1.145, 1.250] | 1.181 [1.143, 1.242] | 1.188 [1.147, 1.249] | 1.190 [1.146, 1.260] |
| Arteries | 1.263 [1.221, 1.332] | 1.261 [1.221, 1.331] | 1.263 [1.222, 1.333] | 1.265 [1.222, 1.335] |
| Veins | 1.200 [1.162, 1.261] | 1.200 [1.163, 1.260] | 1.200 [1.160, 1.262] | 1.200 [1.161, 1.263] |
| **Tortuosity** | | | | |
| **Tortuosity Density** | | | | |

| | | | | |
|---|---|---|---|---|
| Vessels in macular region | 0.624 [0.586, 0.662] | 0.623 [0.585, 0.660] | 0.624 [0.585, 0.664] | 0.626 [0.586, 0.663] |
| Arteries | 0.543 [0.517, 0.569] | 0.542 [0.518, 0.567] | 0.543 [0.517, 0.569] | 0.544 [0.518, 0.571] |
| Veins | 0.591 [0.568, 0.612] | 0.589 [0.567, 0.611] | 0.592 [0.570, 0.613] | 0.591 [0.568, 0.614] |
| **Curve Angle (Degree)** | | | | |
| Vessels in macular region | 11.782 [10.780, 12.942] | 11.790 [10.813, 12.912] | 11.785 [10.783, 12.936] | 11.764 [10.699, 13.006] |
| Arteries | 9.477 [8.614, 10.548] | 9.651 [8.773, 10.730] | 9.434 [8.592, 10.494] | 9.240 [8.393, 10.366] |
| Veins | 10.048 [9.417, 10.762] | 10.056 [9.476, 10.741] | 10.070 [9.444, 10.792] | 9.981 [9.273, 10.749] |
| **Fractal Tortuosity** | | | | |
| Vessels in macular region | 0.834 [0.826, 0.843] | 0.833 [0.825, 0.841] | 0.835 [0.827, 0.843] | 0.836 [0.828, 0.844] |
| Arteries | 0.840 [0.834, 0.846] | 0.838 [0.833, 0.844] | 0.840 [0.834, 0.846] | 0.842 [0.836, 0.848] |
| Veins | 0.838 [0.833, 0.844] | 0.837 [0.832, 0.843] | 0.838 [0.833, 0.843] | 0.840 [0.834, 0.846] |
| **Inflection Count Tortuosity** | | | | |
| Vessels in macular region | 0.260 [0.242, 0.278] | 0.261 [0.243, 0.278] | 0.261 [0.242, 0.279] | 0.258 [0.240, 0.277] |
| Arteries | 0.227 [0.214, 0.238] | 0.228 [0.215, 0.239] | 0.227 [0.215, 0.238] | 0.225 [0.212, 0.238] |
| Veins | 0.246 [0.234, 0.256] | 0.246 [0.235, 0.257] | 0.246 [0.234, 0.256] | 0.244 [0.231, 0.255] |
| **Distance-based Tortuosity** | | | | |
| Vessels in macular region | 1.083 [1.078, 1.089] | 1.083 [1.078, 1.089] | 1.082 [1.078, 1.088] | 1.082 [1.078, 1.088] |
| Arteries | 1.092 [1.087, 1.099] | 1.092 [1.087, 1.098] | 1.093 [1.087, 1.099] | 1.093 [1.087, 1.100] |
| Veins | 1.083 [1.080, 1.087] | 1.083 [1.080, 1.087] | 1.083 [1.080, 1.087] | 1.083 [1.080, 1.087] |
| **Linear Regression Tortuosity** | | | | |
| Vessels in macular region | 2.808 [2.138, 4.263] | 2.782 [2.141, 4.198] | 2.797 [2.115, 4.260] | 2.899 [2.188, 4.370] |
| Arteries | 3.086 [2.288, 4.580] | 3.099 [2.329, 4.550] | 3.038 [2.280, 4.560] | 3.143 [2.255, 4.639] |
| Veins | 2.706 [2.145, 3.692] | 2.699 [2.146, 3.646] | 2.666 [2.138, 3.654] | 2.783 [2.157, 3.804] |
| **Angle-based Tortuosity** | | | | |
| Vessels in macular region | 0.069 [0.039, 0.104] | 0.067 [0.039, 0.102] | 0.069 [0.040, 0.105] | 0.070 [0.039, 0.109] |
| Arteries | 0.070 [0.046, 0.097] | 0.070 [0.046, 0.096] | 0.070 [0.046, 0.097] | 0.069 [0.044, 0.097] |
| Veins | 0.062 [0.042, 0.085] | 0.061 [0.042, 0.083] | 0.063 [0.042, 0.085] | 0.063 [0.042, 0.088] |
| **Squared Curvature Tortuosity** | | | | |
| Vessels in macular region | 0.128 [0.118, 0.140] | 0.128 [0.117, 0.139] | 0.128 [0.118, 0.139] | 0.129 [0.118, 0.141] |
| Arteries | 0.116 [0.108, 0.124] | 0.115 [0.108, 0.124] | 0.116 [0.108, 0.124] | 0.116 [0.108, 0.125] |

| | | | | |
|---|---|---|---|---|
| Veins | 0.116 [0.110, 0.123] | 0.115 [0.109, 0.122] | 0.116 [0.110, 0.123] | 0.117 [0.110, 0.124] |
| **Turning Angles** | | | | |
| Vessels in macular region | 0.368 [0.356, 0.379] | 0.368 [0.356, 0.379] | 0.368 [0.356, 0.379] | 0.368 [0.356, 0.379] |
| Arteries | 0.376 [0.367, 0.385] | 0.376 [0.368, 0.385] | 0.376 [0.367, 0.385] | 0.376 [0.367, 0.386] |
| Veins | 0.370 [0.362, 0.378] | 0.370 [0.362, 0.379] | 0.370 [0.362, 0.378] | 0.370 [0.362, 0.378] |

Supplementary Table 3. Normative data of retinal vascular parameters by sex and age groups

A. Normative data in females

| Retinal Vascular Measurements | Female | | | | P value |
|---|---|---|---|---|---|
| | Overall (n=6476) | 40-50 years (n=2444) | 50-60 years (n=2455) | >60 years (n=1577) | |
| **Calibre** | | | | | |
| **Artery-to-Vein Ratio equivalent** | 0.655 (0.068) | 0.659 (0.069) | 0.655 (0.068) | 0.650 (0.065) | <0.001 |
| **Central Retinal Artery Equivalent (μm)** | 152.812 (14.777) | 154.244 (14.779) | 152.722 (14.770) | 150.731 (14.537) | <0.001 |
| **Central Retinal Vein Equivalent (μm)** | 233.429 (21.283) | 234.328 (21.261) | 233.328 (21.143) | 232.193 (21.479) | 0.028 |
| **Length-to-Diameter Ratio** | | | | | |
| Vessels in macular region | 13.587 (1.870) | 13.326 (1.786) | 13.607 (1.837) | 13.962 (1.981) | <0.001 |
| Arteries | 12.807 (1.540) | 12.432 (1.376) | 12.830 (1.510) | 13.351 (1.655) | <0.001 |
| Veins | 12.187 (1.062) | 11.988 (0.959) | 12.185 (1.059) | 12.498 (1.144) | <0.001 |
| **Width (μm)** | | | | | |
| Vessels in macular region | 48.461 (2.920) | 48.359 (2.827) | 48.436 (2.950) | 48.660 (3.006) | 0.011 |
| Arteries | 56.851 (2.930) | 56.621 (2.835) | 56.793 (2.910) | 57.296 (3.055) | <0.001 |
| Veins | 63.402 (3.556) | 62.794 (3.331) | 63.349 (3.517) | 64.426 (3.722) | <0.001 |
| Terminal arteries | 39.884 (1.976) | 39.476 (1.786) | 39.835 (1.921) | 40.591 (2.141) | <0.001 |
| Terminal veins | 42.211 (2.271) | 41.709 (2.087) | 42.195 (2.259) | 43.012 (2.338) | <0.001 |
| Nonterminal arteries | 72.749 (4.338) | 72.613 (4.258) | 72.674 (4.327) | 73.078 (4.464) | 0.002 |
| Nonterminal veins | 83.632 (5.373) | 82.843 (5.080) | 83.541 (5.291) | 84.996 (5.672) | <0.001 |
| **Complexity** | | | | | |
| **Fractal Dimension** | | | | | |
| All vessels | 1.721 (0.032) | 1.731 (0.028) | 1.720 (0.032) | 1.706 (0.033) | <0.001 |
| Arteries | 1.526 (0.039) | 1.538 (0.034) | 1.525 (0.038) | 1.508 (0.038) | <0.001 |
| Veins | 1.562 (0.025) | 1.568 (0.023) | 1.561 (0.025) | 1.553 (0.025) | <0.001 |
| **Level (N)** | | | | | |
| Vessels in macular region | 3.161 (0.719) | 3.186 (0.700) | 3.157 (0.728) | 3.128 (0.731) | 0.012 |
| Arteries | 4.101 (0.576) | 4.184 (0.556) | 4.109 (0.571) | 3.961 (0.586) | <0.001 |
| Veins | 3.946 (0.455) | 4.003 (0.449) | 3.947 (0.454) | 3.855 (0.453) | <0.001 |
| **Strahler (N)** | | | | | |
| Vessels in macular region | 1.662 (0.102) | 1.667 (0.100) | 1.663 (0.102) | 1.655 (0.104) | <0.001 |
| Arteries | 1.817 (0.075) | 1.828 (0.071) | 1.817 (0.075) | 1.800 (0.078) | <0.001 |
| Veins | 1.801 (0.064) | 1.807 (0.063) | 1.802 (0.064) | 1.791 (0.065) | <0.001 |
| **Number of segments (N)** | | | | | |
| Vessels in macular region | 84.969 (21.860) | 90.485 (20.603) | 85.007 (21.396) | 76.361 (21.704) | <0.001 |
| Arteries | 172.061 (45.180) | 188.613 (42.304) | 170.699 (43.513) | 148.527 (41.015) | <0.001 |
| Veins | 172.124 (33.100) | 182.576 (30.610) | 171.983 (32.326) | 156.143 (31.570) | <0.001 |
| **Number of trees (N)** | | | | | |

| | | | | | |
|---|---|---|---|---|---|
| Vessels in macular region | 11.173 (2.763) | 11.246 (2.793) | 11.081 (2.738) | 11.203 (2.752) | 0.152 |
| Arteries | 11.141 (3.014) | 11.436 (3.104) | 11.029 (2.953) | 10.859 (2.929) | <0.001 |
| Veins | 12.585 (3.028) | 12.692 (3.064) | 12.465 (2.991) | 12.604 (3.026) | 0.058 |
| **Number of branching** | | | | | |
| Vessels in macular region | 47.238 (14.119) | 50.337 (13.562) | 47.238 (13.878) | 42.436 (14.002) | <0.001 |
| Arteries | 97.370 (28.389) | 107.318 (26.771) | 96.556 (27.408) | 83.220 (25.992) | <0.001 |
| Veins | 98.701 (22.660) | 105.267 (21.140) | 98.576 (22.289) | 88.721 (21.832) | <0.001 |
| **Number of bifurcation (N)** | | | | | |
| Vessels in macular region | 26.626 (8.686) | 28.419 (8.569) | 26.594 (8.598) | 23.897 (8.282) | <0.001 |
| Arteries | 51.618 (15.049) | 56.363 (14.592) | 51.213 (14.681) | 44.894 (13.579) | <0.001 |
| Veins | 49.830 (11.163) | 52.443 (11.065) | 49.884 (10.993) | 45.696 (10.312) | <0.001 |
| **Number of Terminal point** | | | | | |
| Arteries | 82.936 (21.183) | 90.728 (19.810) | 82.265 (20.409) | 71.904 (19.238) | <0.001 |
| Veins | 83.951 (15.762) | 88.901 (14.680) | 83.873 (15.396) | 76.401 (14.918) | <0.001 |
| **Number of non-terminal point** | | | | | |
| Arteries | 89.073 (24.365) | 97.800 (22.892) | 88.394 (23.503) | 76.604 (22.184) | <0.001 |
| Veins | 88.141 (17.755) | 93.630 (16.406) | 88.078 (17.377) | 79.732 (17.036) | <0.001 |
| **Density** | | | | | |
| **Vessel Area Density (%)** | | | | | |
| Arteries | 5.756 (0.917) | 6.032 (0.861) | 5.725 (0.906) | 5.375 (0.872) | <0.001 |
| Arteries in macular region | 5.314 (1.148) | 5.604 (1.053) | 5.296 (1.123) | 4.894 (1.196) | <0.001 |
| Arteries outside macular region | 5.854 (0.945) | 6.122 (0.907) | 5.820 (0.934) | 5.490 (0.886) | <0.001 |
| Veins | 7.194 (0.733) | 7.332 (0.698) | 7.186 (0.734) | 6.994 (0.737) | <0.001 |
| Veins in macular region | 4.693 (0.948) | 4.892 (0.856) | 4.709 (0.940) | 4.359 (1.004) | <0.001 |
| Veins outside macular region | 7.814 (0.775) | 7.929 (0.754) | 7.798 (0.779) | 7.660 (0.773) | <0.001 |
| **Vessel Skeleton Density (%)** | | | | | |
| Arteries | 1.446 (0.241) | 1.531 (0.222) | 1.439 (0.235) | 1.326 (0.226) | <0.001 |
| Arteries in macular region | 1.671 (0.338) | 1.764 (0.298) | 1.671 (0.329) | 1.527 (0.359) | <0.001 |
| Arteries outside macular region | 1.385 (0.236) | 1.467 (0.222) | 1.377 (0.230) | 1.271 (0.213) | <0.001 |
| Veins | 1.560 (0.177) | 1.611 (0.166) | 1.559 (0.173) | 1.483 (0.172) | <0.001 |
| Veins in macular region | 1.548 (0.299) | 1.621 (0.267) | 1.553 (0.293) | 1.426 (0.313) | <0.001 |
| Veins outside macular region | 1.561 (0.164) | 1.605 (0.158) | 1.558 (0.162) | 1.496 (0.155) | <0.001 |
| **Branching Density (%)** | | | | | |
| Vessels in macular region | 1.456 (0.219) | 1.488 (0.212) | 1.454 (0.218) | 1.410 (0.224) | <0.001 |
| Arteries | 1.389 (0.173) | 1.437 (0.156) | 1.385 (0.171) | 1.320 (0.177) | <0.001 |
| Veins | 1.337 (0.141) | 1.373 (0.127) | 1.336 (0.140) | 1.284 (0.146) | <0.001 |
| **Bifurcation Density (%)** | | | | | |
| Vessels in macular region | 1.427 (0.244) | 1.453 (0.240) | 1.424 (0.239) | 1.392 (0.254) | <0.001 |
| Arteries | 1.330 (0.206) | 1.381 (0.191) | 1.328 (0.203) | 1.251 (0.209) | <0.001 |

| | | | | | |
|---|---|---|---|---|---|
| Veins | 1.286 (0.160) | 1.318 (0.155) | 1.287 (0.159) | 1.238 (0.159) | <0.001 |
| **Chord Length (μm)** | | | | | |
| Vessels in macular region | 539.939 (73.776) | 527.746 (68.412) | 540.401 (72.235) | 558.118 (80.131) | <0.001 |
| Arteries | 584.350 (82.905) | 559.714 (69.654) | 585.582 (82.078) | 620.613 (89.269) | <0.001 |
| Veins | 598.607 (63.376) | 581.358 (53.804) | 598.676 (62.766) | 625.233 (68.624) | <0.001 |
| **Arc Length (μm)** | | | | | |
| Vessels in macular region | 591.592 (81.691) | 577.850 (76.098) | 592.203 (79.722) | 611.937 (88.527) | <0.001 |
| Arteries | 634.493 (89.771) | 607.567 (75.286) | 635.842 (88.878) | 674.121 (96.567) | <0.001 |
| Veins | 649.655 (69.104) | 630.691 (58.689) | 649.738 (68.216) | 678.917 (74.968) | <0.001 |
| Nonterminal arteries | 670.441 (108.539) | 637.823 (90.648) | 672.073 (106.813) | 718.452 (118.058) | <0.001 |
| Nonterminal veins | 691.363 (82.606) | 668.605 (70.618) | 691.823 (81.278) | 725.916 (89.558) | <0.001 |
| Terminal arteries | 595.623 (82.208) | 575.751 (72.539) | 596.901 (81.392) | 624.430 (88.595) | <0.001 |
| Terminal veins | 606.041 (71.314) | 591.453 (63.335) | 605.632 (71.660) | 629.288 (76.221) | <0.001 |
| **Branching Angle** | | | | | |
| **Angular Asymmetry (Degree)** | | | | | |
| Vessels in macular region | 35.658 (4.624) | 35.694 (4.470) | 35.621 (4.613) | 35.661 (4.873) | 0.81 |
| Arteries | 41.223 (3.720) | 41.548 (3.598) | 41.195 (3.689) | 40.760 (3.902) | <0.001 |
| Veins | 39.549 (3.447) | 39.935 (3.351) | 39.507 (3.459) | 39.017 (3.501) | <0.001 |
| **Junctional Exponent Deviation** | | | | | |
| Vessels in macular region | 0.049 (0.107) | 0.053 (0.103) | 0.046 (0.107) | 0.047 (0.112) | 0.05 |
| Arteries | -0.095 (0.076) | -0.094 (0.074) | -0.095 (0.076) | -0.094 (0.079) | 0.87 |
| Veins | -0.007 (0.070) | -0.009 (0.068) | -0.006 (0.069) | -0.005 (0.075) | 0.146 |
| **Angular Asymmetry at edge (Degree)** | | | | | |
| Vessels in macular region | 39.905 (5.231) | 39.705 (4.892) | 39.837 (5.192) | 40.321 (5.754) | 0.035 |
| Arteries | 42.991 (3.965) | 43.221 (3.780) | 42.954 (3.912) | 42.692 (4.295) | <0.001 |
| Veins | 43.376 (3.574) | 43.539 (3.362) | 43.362 (3.627) | 43.144 (3.794) | 0.003 |
| **Asymmetry Ratio** | | | | | |
| Vessels in macular region | 1.766 (0.317) | 1.761 (0.302) | 1.770 (0.317) | 1.766 (0.340) | 0.533 |
| Arteries | 2.140 (0.338) | 2.153 (0.330) | 2.137 (0.339) | 2.127 (0.347) | 0.022 |
| Veins | 2.491 (0.438) | 2.486 (0.430) | 2.494 (0.432) | 2.494 (0.459) | 0.875 |
| **Branching Angle (Degree)** | | | | | |
| Vessels in macular region | 45.852 (2.220) | 45.769 (2.183) | 45.822 (2.199) | 46.028 (2.299) | 0.001 |
| Arteries | 47.203 (1.748) | 47.317 (1.685) | 47.155 (1.770) | 47.101 (1.803) | <0.001 |
| Veins | 46.658 (1.356) | 46.595 (1.332) | 46.678 (1.344) | 46.722 (1.409) | 0.015 |
| **Branching Angle at edge (Degree)** | | | | | |
| Vessels in macular region | 42.015 (3.105) | 41.931 (2.905) | 41.950 (3.033) | 42.245 (3.484) | 0.046 |
| Arteries | 43.636 (2.505) | 43.839 (2.365) | 43.581 (2.534) | 43.407 (2.644) | <0.001 |
| Veins | 42.529 (1.969) | 42.601 (1.842) | 42.546 (1.953) | 42.388 (2.168) | 0.003 |
| **Branching Coefficient** | | | | | |

| | | | | | |
|---|---|---|---|---|---|
| Vessels in macular region | 1.210 (0.092) | 1.206 (0.090) | 1.210 (0.090) | 1.216 (0.099) | 0.014 |
| Arteries | 1.289 (0.095) | 1.288 (0.095) | 1.288 (0.096) | 1.291 (0.095) | 0.331 |
| Veins | 1.224 (0.089) | 1.223 (0.087) | 1.222 (0.088) | 1.227 (0.094) | 0.708 |
| **Tortuosity** | | | | | |
| **Tortuosity Density** | | | | | |
| Vessels in macular region | 0.625 (0.055) | 0.623 (0.054) | 0.625 (0.056) | 0.627 (0.057) | 0.112 |
| Arteries | 0.543 (0.038) | 0.542 (0.036) | 0.544 (0.038) | 0.546 (0.039) | 0.005 |
| Veins | 0.590 (0.032) | 0.588 (0.032) | 0.591 (0.032) | 0.590 (0.033) | 0.005 |
| **Curve Angle (Degree)** | | | | | |
| Vessels in macular region | 11.978 (1.621) | 11.967 (1.571) | 11.970 (1.619) | 12.009 (1.701) | 0.846 |
| Arteries | 9.743 (1.484) | 9.880 (1.472) | 9.708 (1.479) | 9.585 (1.493) | <0.001 |
| Veins | 10.121 (0.988) | 10.129 (0.946) | 10.142 (0.999) | 10.076 (1.034) | 0.049 |
| **Fractal Tortuosity** | | | | | |
| Vessels in macular region | 0.835 (0.012) | 0.834 (0.012) | 0.835 (0.012) | 0.836 (0.012) | <0.001 |
| Arteries | 0.840 (0.009) | 0.839 (0.008) | 0.840 (0.009) | 0.842 (0.009) | <0.001 |
| Veins | 0.838 (0.008) | 0.838 (0.008) | 0.838 (0.008) | 0.840 (0.009) | <0.001 |
| **Inflection Count Tortuosity** | | | | | |
| Vessels in macular region | 0.260 (0.026) | 0.260 (0.025) | 0.260 (0.026) | 0.259 (0.027) | 0.11 |
| Arteries | 0.225 (0.017) | 0.226 (0.017) | 0.225 (0.017) | 0.225 (0.018) | 0.079 |
| Veins | 0.245 (0.016) | 0.245 (0.016) | 0.245 (0.016) | 0.243 (0.017) | <0.001 |
| **Distance-based Tortuosity** | | | | | |
| Vessels in macular region | 1.094 (0.009) | 1.093 (0.008) | 1.094 (0.009) | 1.095 (0.009) | <0.001 |
| Arteries | 1.085 (0.009) | 1.085 (0.009) | 1.085 (0.008) | 1.085 (0.009) | 0.639 |
| Veins | 1.084 (0.005) | 1.084 (0.005) | 1.084 (0.005) | 1.084 (0.005) | 0.017 |
| **Linear Regression Tortuosity** | | | | | |
| Vessels in macular region | 3.726 (2.592) | 3.687 (2.517) | 3.684 (2.609) | 3.852 (2.677) | 0.008 |
| Arteries | 4.039 (2.733) | 3.969 (2.612) | 4.066 (2.788) | 4.108 (2.828) | 0.919 |
| Veins | 3.256 (1.716) | 3.215 (1.686) | 3.238 (1.733) | 3.348 (1.734) | 0.001 |
| **Angle-based Tortuosity** | | | | | |
| Vessels in macular region | 0.075 (0.048) | 0.074 (0.046) | 0.075 (0.047) | 0.077 (0.050) | 0.690 |
| Arteries | 0.074 (0.037) | 0.074 (0.036) | 0.074 (0.037) | 0.074 (0.039) | 0.880 |
| Veins | 0.065 (0.031) | 0.064 (0.030) | 0.065 (0.031) | 0.067 (0.033) | 0.169 |
| **Squared Curvature Tortuosity** | | | | | |
| Vessels in macular region | 0.129 (0.016) | 0.129 (0.016) | 0.129 (0.016) | 0.131 (0.017) | 0.001 |
| Arteries | 0.117 (0.012) | 0.117 (0.012) | 0.117 (0.012) | 0.117 (0.012) | 0.440 |
| Veins | 0.117 (0.010) | 0.116 (0.009) | 0.117 (0.010) | 0.118 (0.010) | <0.001 |
| **Turning Angles** | | | | | |
| Vessels in macular region | 0.368 (0.017) | 0.368 (0.016) | 0.368 (0.017) | 0.368 (0.017) | 0.727 |
| Arteries | 0.376 (0.013) | 0.376 (0.012) | 0.376 (0.013) | 0.376 (0.014) | 0.654 |

| | | | | | |
|---|---|---|---|---|---|
| Veins | 0.370 (0.012) | 0.370 (0.012) | 0.370 (0.012) | 0.370 (0.013) | 0.553 |

#### B. Normative data in males

| Retinal Vascular Measurements | Male | | | | P value |
|---|---|---|---|---|---|
| | Overall (n=3675) | 40-50 years (n=1444) | 50-60 years (n=1348) | >60 years (n=883) | |
| **Calibre** | | | | | |
| **Artery-to-Vein Ratio equivalent** | 0.648 (0.069) | 0.651 (0.069) | 0.647 (0.068) | 0.643 (0.070) | 0.006 |
| **Central Retinal Artery Equivalent (μm)** | 150.630 (15.021) | 152.454 (15.092) | 149.833 (15.134) | 148.862 (14.420) | <0.001 |
| **Central Retinal Vein Equivalent (μm)** | 232.885 (21.949) | 234.507 (22.204) | 231.851 (21.868) | 231.810 (21.510) | 0.014 |
| **Length-to-Diameter Ratio** | | | | | |
| Vessels in macular region | 13.540 (1.846) | 13.225 (1.643) | 13.557 (1.850) | 14.031 (2.036) | <0.001 |
| Arteries | 12.897 (1.491) | 12.499 (1.302) | 12.930 (1.460) | 13.497 (1.614) | <0.001 |
| Veins | 12.095 (1.048) | 11.894 (0.932) | 12.096 (1.025) | 12.420 (1.174) | <0.001 |
| **Width (μm)** | | | | | |
| Vessels in macular region | 48.200 (2.921) | 48.087 (2.788) | 48.090 (2.951) | 48.551 (3.062) | <0.001 |
| Arteries | 56.152 (2.946) | 55.928 (2.717) | 56.038 (2.990) | 56.690 (3.167) | <0.001 |
| Veins | 62.994 (3.456) | 62.434 (3.154) | 62.969 (3.477) | 63.949 (3.687) | <0.001 |
| Terminal arteries | 39.514 (1.927) | 39.129 (1.674) | 39.507 (1.923) | 40.155 (2.141) | <0.001 |
| Terminal veins | 41.862 (2.172) | 41.437 (1.977) | 41.892 (2.168) | 42.509 (2.314) | <0.001 |
| Nonterminal arteries | 71.862 (4.387) | 71.694 (4.132) | 71.655 (4.435) | 72.454 (4.662) | <0.001 |
| Nonterminal veins | 83.116 (5.215) | 82.352 (4.854) | 83.014 (5.201) | 84.520 (5.523) | <0.001 |
| **Complexity** | | | | | |
| **Fractal Dimension** | | | | | |
| All vessels | 1.723 (0.032) | 1.734 (0.028) | 1.722 (0.030) | 1.706 (0.034) | <0.001 |
| Arteries | 1.526 (0.039) | 1.540 (0.035) | 1.525 (0.036) | 1.506 (0.040) | <0.001 |
| Veins | 1.565 (0.025) | 1.572 (0.023) | 1.564 (0.024) | 1.554 (0.026) | <0.001 |
| **Level (N)** | | | | | |
| Vessels in macular region | 3.169 (0.714) | 3.185 (0.685) | 3.162 (0.723) | 3.153 (0.745) | 0.275 |
| Arteries | 4.131 (0.558) | 4.223 (0.537) | 4.123 (0.555) | 3.993 (0.568) | <0.001 |
| Veins | 3.990 (0.452) | 4.044 (0.430) | 3.975 (0.454) | 3.923 (0.473) | <0.001 |
| **Strahler (N)** | | | | | |
| Vessels in macular region | 1.661 (0.101) | 1.667 (0.097) | 1.658 (0.101) | 1.658 (0.108) | 0.028 |
| Arteries | 1.815 (0.073) | 1.826 (0.070) | 1.815 (0.073) | 1.800 (0.076) | <0.001 |
| Veins | 1.807 (0.064) | 1.814 (0.060) | 1.807 (0.065) | 1.797 (0.066) | <0.001 |
| **Number of segments (N)** | | | | | |
| Vessels in macular region | 86.317 (22.122) | 92.602 (20.607) | 85.852 (20.917) | 76.751 (22.781) | <0.001 |
| Arteries | 174.315 (45.284) | 191.447 (41.707) | 172.492 (42.080) | 149.081 (43.227) | <0.001 |
| Veins | 178.143 (34.127) | 189.640 (31.253) | 177.146 (32.267) | 160.866 (33.861) | <0.001 |

| | | | | | |
|---|---|---|---|---|---|
| **Number of trees (N)** | | | | | |
| Vessels in macular region | 11.216 (2.811) | 11.295 (2.775) | 11.200 (2.858) | 11.110 (2.795) | 0.215 |
| Arteries | 11.354 (3.068) | 11.700 (3.185) | 11.276 (2.977) | 10.905 (2.945) | <0.001 |
| Veins | 12.580 (3.052) | 12.722 (3.056) | 12.457 (3.066) | 12.537 (3.019) | 0.042 |
| **Number of branching** | | | | | |
| Vessels in macular region | 48.346 (14.408) | 51.824 (13.706) | 48.175 (13.833) | 42.918 (14.691) | <0.001 |
| Arteries | 98.794 (28.631) | 108.961 (26.543) | 97.718 (26.878) | 83.810 (27.621) | <0.001 |
| Veins | 102.407 (23.590) | 109.666 (22.286) | 101.486 (22.359) | 91.941 (23.340) | <0.001 |
| **Number of bifurcation (N)** | | | | | |
| Vessels in macular region | 26.899 (8.700) | 28.972 (8.543) | 26.612 (8.411) | 23.948 (8.482) | <0.001 |
| Arteries | 52.315 (15.038) | 57.304 (14.516) | 51.796 (14.114) | 44.950 (14.054) | <0.001 |
| Veins | 51.711 (11.307) | 54.672 (10.864) | 51.537 (10.922) | 47.135 (11.041) | <0.001 |
| **Number of Terminal point** | | | | | |
| Arteries | 84.348 (21.388) | 92.322 (19.632) | 83.514 (19.962) | 72.581 (20.557) | <0.001 |
| Veins | 86.769 (16.346) | 92.211 (15.128) | 86.261 (15.434) | 78.645 (16.120) | <0.001 |
| **Number of non-terminal point** | | | | | |
| Arteries | 89.923 (24.268) | 99.023 (22.432) | 88.962 (22.591) | 76.508 (23.096) | <0.001 |
| Veins | 91.285 (18.109) | 97.268 (16.415) | 90.814 (17.226) | 82.218 (18.188) | <0.001 |
| **Density** | | | | | |
| **Vessel Area Density (%)** | | | | | |
| Arteries | 5.731 (0.913) | 6.030 (0.866) | 5.692 (0.857) | 5.302 (0.887) | <0.001 |
| Arteries in macular region | 5.267 (1.146) | 5.590 (1.037) | 5.241 (1.094) | 4.777 (1.213) | <0.001 |
| Arteries outside macular region | 5.836 (0.937) | 6.124 (0.912) | 5.790 (0.888) | 5.436 (0.890) | <0.001 |
| Veins | 7.254 (0.736) | 7.440 (0.692) | 7.233 (0.725) | 6.981 (0.736) | <0.001 |
| Veins in macular region | 4.803 (0.930) | 5.003 (0.849) | 4.787 (0.926) | 4.499 (0.979) | <0.001 |
| Veins outside macular region | 7.852 (0.778) | 8.023 (0.748) | 7.829 (0.762) | 7.609 (0.780) | <0.001 |
| **Vessel Skeleton Density (%)** | | | | | |
| Arteries | 1.463 (0.244) | 1.552 (0.224) | 1.455 (0.226) | 1.330 (0.241) | <0.001 |
| Arteries in macular region | 1.675 (0.342) | 1.781 (0.294) | 1.673 (0.320) | 1.507 (0.377) | <0.001 |
| Arteries outside macular region | 1.406 (0.238) | 1.491 (0.226) | 1.397 (0.222) | 1.283 (0.225) | <0.001 |
| Veins | 1.589 (0.179) | 1.649 (0.163) | 1.584 (0.172) | 1.500 (0.178) | <0.001 |
| Veins in macular region | 1.591 (0.297) | 1.670 (0.262) | 1.587 (0.293) | 1.466 (0.315) | <0.001 |
| Veins outside macular region | 1.586 (0.167) | 1.639 (0.157) | 1.580 (0.159) | 1.507 (0.162) | <0.001 |
| **Branching Density (%)** | | | | | |
| Vessels in macular region | 1.468 (0.216) | 1.504 (0.204) | 1.469 (0.212) | 1.409 (0.227) | <0.001 |
| Arteries | 1.389 (0.171) | 1.438 (0.155) | 1.385 (0.163) | 1.314 (0.180) | <0.001 |
| Veins | 1.355 (0.142) | 1.389 (0.129) | 1.354 (0.139) | 1.304 (0.152) | <0.001 |
| **Bifurcation Density (%)** | | | | | |
| Vessels in macular region | 1.444 (0.243) | 1.479 (0.237) | 1.439 (0.243) | 1.396 (0.244) | <0.001 |

| | | | | | |
|---|---|---|---|---|---|
| Arteries | 1.335 (0.200) | 1.387 (0.183) | 1.334 (0.195) | 1.251 (0.203) | <0.001 |
| Veins | 1.300 (0.157) | 1.331 (0.149) | 1.296 (0.154) | 1.255 (0.165) | <0.001 |
| **Chord Length (µm)** | | | | | |
| Vessels in macular region | 535.854 (73.501) | 520.641 (63.606) | 535.910 (71.952) | 560.648 (83.572) | <0.001 |
| Arteries | 583.287 (81.348) | 558.501 (68.628) | 584.034 (77.135) | 622.682 (90.623) | <0.001 |
| Veins | 590.546 (62.379) | 574.416 (54.039) | 591.264 (60.002) | 615.827 (69.759) | <0.001 |
| **Arc Length (µm)** | | | | | |
| Vessels in macular region | 586.483 (81.027) | 569.848 (70.575) | 586.532 (79.260) | 613.613 (91.734) | <0.001 |
| Arteries | 632.882 (87.981) | 606.256 (73.948) | 633.455 (83.770) | 675.548 (97.963) | <0.001 |
| Veins | 640.692 (68.125) | 622.968 (59.086) | 641.485 (65.462) | 668.465 (76.084) | <0.001 |
| Nonterminal arteries | 665.752 (106.454) | 633.032 (88.616) | 667.121 (101.031) | 717.168 (119.862) | <0.001 |
| Nonterminal veins | 680.096 (80.434) | 658.757 (70.146) | 681.930 (77.581) | 712.191 (89.026) | <0.001 |
| Terminal arteries | 597.446 (80.655) | 577.669 (70.465) | 597.821 (79.134) | 629.216 (88.086) | <0.001 |
| Terminal veins | 599.675 (69.654) | 586.195 (62.599) | 599.336 (67.387) | 622.238 (77.823) | <0.001 |
| **Branching Angle** | | | | | |
| **Angular Asymmetry (Degree)** | | | | | |
| Vessels in macular region | 35.485 (4.520) | 35.589 (4.381) | 35.420 (4.494) | 35.414 (4.780) | 0.496 |
| Arteries | 40.973 (3.758) | 41.279 (3.546) | 41.021 (3.866) | 40.398 (3.867) | <0.001 |
| Veins | 39.129 (3.394) | 39.390 (3.361) | 39.052 (3.335) | 38.821 (3.508) | <0.001 |
| **Junctional Exponent Deviation** | | | | | |
| Vessels in macular region | 0.050 (0.107) | 0.055 (0.103) | 0.047 (0.109) | 0.047 (0.112) | 0.124 |
| Arteries | -0.095 (0.076) | -0.096 (0.072) | -0.097 (0.075) | -0.092 (0.081) | 0.279 |
| Veins | -0.008 (0.070) | -0.013 (0.068) | -0.008 (0.070) | 0.000 (0.072) | <0.001 |
| **Angular Asymmetry at edge (Degree)** | | | | | |
| Vessels in macular region | 39.760 (5.122) | 39.640 (4.788) | 39.703 (5.010) | 40.044 (5.774) | 0.339 |
| Arteries | 42.669 (3.983) | 42.870 (3.789) | 42.680 (3.987) | 42.324 (4.257) | 0.006 |
| Veins | 43.054 (3.477) | 43.279 (3.431) | 42.954 (3.342) | 42.838 (3.727) | 0.005 |
| **Asymmetry Ratio** | | | | | |
| Vessels in macular region | 1.784 (0.316) | 1.777 (0.305) | 1.786 (0.312) | 1.792 (0.341) | 0.679 |
| Arteries | 2.121 (0.328) | 2.125 (0.314) | 2.116 (0.334) | 2.123 (0.341) | 0.532 |
| Veins | 2.477 (0.427) | 2.466 (0.412) | 2.493 (0.429) | 2.468 (0.447) | 0.217 |
| **Branching Angle (Degree)** | | | | | |
| Vessels in macular region | 45.733 (2.178) | 45.650 (2.154) | 45.716 (2.136) | 45.893 (2.273) | 0.034 |
| Arteries | 47.011 (1.697) | 47.126 (1.667) | 47.021 (1.696) | 46.809 (1.731) | <0.001 |
| Veins | 46.678 (1.354) | 46.624 (1.328) | 46.671 (1.362) | 46.779 (1.378) | 0.049 |
| **Branching Angle at edge (Degree)** | | | | | |
| Vessels in macular region | 41.902 (2.937) | 41.943 (2.890) | 41.799 (2.793) | 41.991 (3.213) | 0.255 |
| Arteries | 43.386 (2.388) | 43.666 (2.324) | 43.362 (2.347) | 42.967 (2.491) | <0.001 |
| Veins | 42.483 (1.993) | 42.611 (1.923) | 42.402 (2.008) | 42.398 (2.075) | 0.009 |

| | | | | | |
|---|---|---|---|---|---|
| **Branching Coefficient** | | | | | |
| Vessels in macular region | 1.207 (0.091) | 1.200 (0.086) | 1.211 (0.092) | 1.214 (0.095) | <0.001 |
| Arteries | 1.288 (0.093) | 1.288 (0.094) | 1.290 (0.091) | 1.287 (0.095) | 0.399 |
| Veins | 1.225 (0.089) | 1.227 (0.090) | 1.225 (0.088) | 1.223 (0.088) | 0.357 |
| **Tortuosity** | | | | | |
| **Tortuosity Density** | | | | | |
| Vessels in macular region | 0.622 (0.055) | 0.621 (0.053) | 0.624 (0.056) | 0.620 (0.058) | 0.331 |
| Arteries | 0.543 (0.037) | 0.545 (0.036) | 0.544 (0.037) | 0.541 (0.040) | 0.085 |
| Veins | 0.591 (0.032) | 0.590 (0.032) | 0.592 (0.032) | 0.592 (0.033) | 0.231 |
| **Curve Angle (Degree)** | | | | | |
| Vessels in macular region | 11.872 (1.587) | 11.946 (1.597) | 11.869 (1.559) | 11.756 (1.606) | 0.027 |
| Arteries | 9.638 (1.491) | 9.884 (1.542) | 9.560 (1.417) | 9.355 (1.452) | <0.001 |
| Veins | 10.157 (0.998) | 10.199 (0.974) | 10.167 (0.982) | 10.072 (1.055) | 0.006 |
| **Fractal Tortuosity** | | | | | |
| Vessels in macular region | 0.834 (0.012) | 0.833 (0.012) | 0.835 (0.012) | 0.836 (0.012) | <0.001 |
| Arteries | 0.840 (0.009) | 0.839 (0.008) | 0.840 (0.009) | 0.842 (0.009) | <0.001 |
| Veins | 0.838 (0.008) | 0.837 (0.008) | 0.838 (0.008) | 0.840 (0.009) | <0.001 |
| **Inflection Count Tortuosity** | | | | | |
| Vessels in macular region | 0.260 (0.026) | 0.261 (0.026) | 0.260 (0.026) | 0.258 (0.028) | 0.011 |
| Arteries | 0.227 (0.017) | 0.228 (0.017) | 0.227 (0.017) | 0.225 (0.019) | <0.001 |
| Veins | 0.246 (0.016) | 0.247 (0.016) | 0.246 (0.016) | 0.244 (0.017) | <0.001 |
| **Distance-based Tortuosity** | | | | | |
| Vessels in macular region | 1.093 (0.009) | 1.093 (0.009) | 1.093 (0.009) | 1.093 (0.008) | 0.836 |
| Arteries | 1.084 (0.008) | 1.085 (0.009) | 1.083 (0.008) | 1.083 (0.008) | <0.001 |
| Veins | 1.084 (0.005) | 1.084 (0.005) | 1.084 (0.005) | 1.084 (0.005) | 0.583 |
| **Linear Regression Tortuosity** | | | | | |
| Vessels in macular region | 3.770 (2.694) | 3.692 (2.663) | 3.804 (2.674) | 3.845 (2.773) | 0.100 |
| Arteries | 3.908 (2.624) | 3.966 (2.605) | 3.802 (2.559) | 3.973 (2.747) | 0.045 |
| Veins | 3.257 (1.732) | 3.265 (1.721) | 3.247 (1.695) | 3.258 (1.807) | 0.636 |
| **Angle-based Tortuosity** | | | | | |
| Vessels in macular region | 0.074 (0.046) | 0.072 (0.045) | 0.074 (0.046) | 0.077 (0.049) | 0.100 |
| Arteries | 0.072 (0.036) | 0.072 (0.035) | 0.073 (0.035) | 0.071 (0.038) | 0.045 |
| Veins | 0.064 (0.030) | 0.064 (0.029) | 0.065 (0.030) | 0.065 (0.032) | 0.636 |
| **Squared Curvature Tortuosity** | | | | | |
| Vessels in macular region | 0.128 (0.016) | 0.128 (0.016) | 0.128 (0.016) | 0.128 (0.016) | 0.996 |
| Arteries | 0.116 (0.012) | 0.116 (0.012) | 0.116 (0.012) | 0.116 (0.012) | 0.966 |
| Veins | 0.116 (0.010) | 0.116 (0.009) | 0.116 (0.010) | 0.117 (0.010) | 0.107 |
| **Turning Angles** | | | | | |
| Vessels in macular region | 0.368 (0.017) | 0.367 (0.017) | 0.367 (0.017) | 0.368 (0.017) | 0.503 |

| | | | | | |
|---|---|---|---|---|---|
| Arteries | 0.377 (0.013) | 0.376 (0.012) | 0.376 (0.013) | 0.377 (0.014) | 0.176 |
| Veins | 0.370 (0.012) | 0.370 (0.012) | 0.370 (0.012) | 0.370 (0.012) | 0.996 |

Notes: Kolmogorov-Smirnov was performed to test distribution normality. For normally distributed measurements, the Analysis of Variance was performed to test the intergroup difference, otherwise, the Kruskal-Wallis H test was conducted. P value <0.05 was set for the significance of intergroup differences.

Supplementary Table 4. Correlation between retinal vascular parameters and health metrics

| Categories | Measures | Measurements | Age | IOP | SBP | HbA1c | BMI |
|---|---|---|---|---|---|---|---|
| | | | **r (p)** | **r (p)** | **r (p)** | **r (p)** | **r (p)** |
| Calibre | Artery-to-Vein Ratio equivalents | Artery-to-Vein Ratio equivalents | -0.040 (<0.001) | 0.031 (0.002) | -0.086 (<0.001) | -0.015 (1.000) | -0.047 (<0.001) |
| | Central Retinal Artery Equivalent | Central Retinal Artery Equivalent | -0.072 (<0.001) | 0.009 (1.000) | -0.115 (<0.001) | -0.022 (0.519) | -0.031 (0.001) |
| | Central Retinal Vein Equivalent | Central Retinal Vein Equivalent | -0.028 (0.020) | -0.023 (0.312) | -0.026 (0.070) | -0.004 (1.000) | 0.020 (1.000) |
| | Length Diameter Ratio | Vessels in macular region | 0.097 (<0.001) | 0.001 (1.000) | 0.060 (<0.001) | 0.024 (0.152) | 0.028 (0.010) |
| | | Arteries | 0.179 (<0.001) | -0.002 (1.000) | 0.126 (<0.001) | 0.048 (<0.001) | 0.046 (<0.001) |
| | | Veins | 0.132 (<0.001) | 0.012 (1.000) | 0.044 (<0.001) | 0.041 (<0.001) | 0.002 (1.000) |
| | Mean Width | Vessels in macular region | 0.033 (<0.001) | -0.014 (1.000) | -0.047 (<0.001) | 0.021 (1.000) | -0.021 (0.936) |
| | | Arteries | 0.060 (<0.001) | -0.008 (1.000) | -0.108 (<0.001) | 0.026 (0.060) | -0.036 (<0.001) |
| | | Veins | 0.119 (<0.001) | -0.028 (0.016) | 0.007 (1.000) | 0.051 (<0.001) | 0.025 (0.081) |
| | | Nonterminal veins | 0.104 (<0.001) | -0.026 (0.059) | -0.003 (1.000) | 0.046 (<0.001) | 0.030 (0.004) |
| | | Terminal veins | 0.151 (<0.001) | -0.017 (1.000) | 0.024 (0.145) | 0.059 (<0.001) | 0.011 (1.000) |
| | | Nonterminal arteries | 0.031 (0.002) | -0.010 (1.000) | -0.118 (<0.001) | 0.016 (1.000) | -0.034 (<0.001) |
| | | Terminal arteries | 0.143 (<0.001) | -0.003 (1.000) | -0.029 (0.006) | 0.048 (<0.001) | -0.014 (1.000) |
| Complexity | Fractal Dimension | Vessels | -0.243 (<0.001) | -0.005 (1.000) | -0.104 (<0.001) | -0.074 (<0.001) | -0.023 (0.345) |
| | | Arteries | -0.236 (<0.001) | 0.007 (1.000) | -0.125 (<0.001) | -0.072 (<0.001) | -0.044 (<0.001) |
| | | Veins | -0.190 (<0.001) | -0.017 (1.000) | -0.069 (<0.001) | -0.059 (<0.001) | 0.006 (1.000) |
| | Level | Vessels in macular region | -0.022 (0.487) | 0.008 (1.000) | -0.017 (1.000) | -0.006 (1.000) | -0.018 (1.000) |
| | | Arteries | -0.110 (<0.001) | -0.008 (1.000) | -0.038 (<0.001) | -0.028 (0.018) | -0.019 (1.000) |
| | | Veins | -0.082 (<0.001) | -0.019 (1.000) | -0.016 (1.000) | -0.031 (0.002) | 0.009 (1.000) |
| | Strahler | Vessels in macular region | -0.032 (0.002) | 0.003 (1.000) | -0.021 (0.744) | -0.005 (1.000) | -0.020 (1.000) |
| | | Arteries | -0.099 (<0.001) | -0.005 (1.000) | -0.044 (<0.001) | -0.029 (0.006) | -0.015 (1.000) |

| | | | | | | | |
|---|---|---|---|---|---|---|---|
| | | Veins | -0.068 (<0.001) | -0.014 (1.000) | -0.008 (1.000) | -0.022 (0.443) | 0.001 (1.000) |
| | Number of segments | Vessels in macular region | -0.188 (<0.001) | -0.004 (1.000) | -0.073 (<0.001) | -0.056 (<0.001) | -0.032 (<0.001) |
| | | Arteries | -0.262 (<0.001) | 0.004 (1.000) | -0.110 (<0.001) | -0.079 (<0.001) | -0.038 (<0.001) |
| | | Veins | -0.234 (<0.001) | -0.011 (1.000) | -0.059 (<0.001) | -0.076 (<0.001) | 0.000 (1.000) |
| | Number of trees | Vessels in macular region | -0.011 (1.000) | 0.007 (1.000) | -0.009 (1.000) | -0.007 (1.000) | -0.004 (1.000) |
| | | Arteries | -0.070 (<0.001) | -0.004 (1.000) | -0.017 (1.000) | -0.019 (1.000) | -0.010 (1.000) |
| | | Veins | -0.014 (1.000) | 0.003 (1.000) | -0.016 (1.000) | -0.011 (1.000) | -0.015 (1.000) |
| | Number of branching | Vessels in macular region | -0.162 (<0.001) | -0.004 (1.000) | -0.066 (<0.001) | -0.050 (<0.001) | -0.028 (0.022) |
| | | Arteries | -0.246 (<0.001) | 0.002 (1.000) | -0.105 (<0.001) | -0.074 (<0.001) | -0.034 (<0.001) |
| | | Veins | -0.210 (<0.001) | -0.009 (1.000) | -0.056 (<0.001) | -0.068 (<0.001) | 0.001 (1.000) |
| | Number of bifurcation | Vessels in macular region | -0.151 (<0.001) | -0.005 (1.000) | -0.057 (<0.001) | -0.046 (<0.001) | -0.021 (1.000) |
| | | Arteries | -0.226 (<0.001) | 0.003 (1.000) | -0.097 (<0.001) | -0.073 (<0.001) | -0.035 (<0.001) |
| | | Veins | -0.176 (<0.001) | -0.015 (1.000) | -0.043 (<0.001) | -0.059 (<0.001) | -0.001 (1.000) |
| | Number of nonterminal point | Nonterminal arteries | -0.257 (<0.001) | 0.004 (1.000) | -0.109 (<0.001) | -0.077 (<0.001) | -0.040 (<0.001) |
| | | Nonterminal veins | -0.230 (<0.001) | -0.013 (1.000) | -0.056 (<0.001) | -0.074 (<0.001) | -0.002 (1.000) |
| | Number of terminal point | Terminal arteries | -0.262 (<0.001) | 0.003 (1.000) | -0.110 (<0.001) | -0.081 (<0.001) | -0.035 (<0.001) |
| | | Terminal veins | -0.232 (<0.001) | -0.010 (1.000) | -0.061 (<0.001) | -0.076 (<0.001) | 0.001 (1.000) |
| Density | Vessel Area Density | Arteries | -0.214 (<0.001) | 0.007 (1.000) | -0.141 (<0.001) | -0.064 (<0.001) | -0.046 (<0.001) |
| | | Arteries in macular region | -0.180 (<0.001) | 0.005 (1.000) | -0.096 (<0.001) | -0.056 (<0.001) | -0.048 (<0.001) |
| | | Arteries outside macular region | -0.199 (<0.001) | 0.007 (1.000) | -0.139 (<0.001) | -0.058 (<0.001) | -0.042 (<0.001) |
| | | Veins | -0.153 (<0.001) | -0.024 (0.198) | -0.070 (<0.001) | -0.043 (<0.001) | 0.011 (1.000) |
| | | Veins in macular region | -0.155 (<0.001) | -0.007 (1.000) | -0.046 (<0.001) | -0.055 (<0.001) | -0.010 (1.000) |
| | | Veins outside macular region | -0.125 (<0.001) | -0.029 (0.008) | -0.070 (<0.001) | -0.031 (0.002) | 0.015 (1.000) |

| | | | | | | | |
|---|---|---|---|---|---|---|---|
| | Vessel Skeleton Density | Arteries | -0.253 (<0.001) | 0.006 (1.000) | -0.105 (<0.001) | -0.079 (<0.001) | -0.035 (<0.001) |
| | | Arteries in macular region | -0.206 (<0.001) | 0.008 (1.000) | -0.083 (<0.001) | -0.069 (<0.001) | -0.037 (<0.001) |
| | | Arteries outside macular region | -0.245 (<0.001) | 0.005 (1.000) | -0.103 (<0.001) | -0.076 (<0.001) | -0.031 (0.002) |
| | | Veins | -0.224 (<0.001) | -0.010 (1.000) | -0.062 (<0.001) | -0.075 (<0.001) | 0.000 (1.000) |
| | | Veins in macular region | -0.188 (<0.001) | -0.001 (1.000) | -0.044 (<0.001) | -0.065 (<0.001) | -0.006 (1.000) |
| | | Veins outside macular region | -0.208 (<0.001) | -0.015 (1.000) | -0.063 (<0.001) | -0.068 (<0.001) | 0.002 (1.000) |
| | Branching Density | Vessels in macular region | -0.101 (<0.001) | -0.001 (1.000) | -0.048 (<0.001) | -0.027 (0.025) | -0.020 (1.000) |
| | | Arteries | -0.193 (<0.001) | -0.000 (1.000) | -0.085 (<0.001) | -0.054 (<0.001) | -0.035 (<0.001) |
| | | Veins | -0.169 (<0.001) | -0.009 (1.000) | -0.046 (<0.001) | -0.052 (<0.001) | -0.012 (1.000) |
| | Bifurcation Density | Vessels in macular region | -0.080 (<0.001) | 0.005 (1.000) | -0.033 (<0.001) | -0.028 (0.013) | -0.021 (0.882) |
| | | Arteries | -0.184 (<0.001) | 0.002 (1.000) | -0.085 (<0.001) | -0.053 (<0.001) | -0.034 (<0.001) |
| | | Veins | -0.137 (<0.001) | -0.001 (1.000) | -0.038 (<0.001) | -0.048 (<0.001) | -0.013 (1.000) |
| | Chord Length | Vessels in macular region | 0.119 (<0.001) | -0.004 (1.000) | 0.050 (<0.001) | 0.033 (<0.001) | 0.023 (0.326) |
| | | Arteries | 0.225 (<0.001) | -0.001 (1.000) | 0.101 (<0.001) | 0.065 (<0.001) | 0.037 (<0.001) |
| | | Veins | 0.192 (<0.001) | 0.004 (1.000) | 0.049 (<0.001) | 0.064 (<0.001) | 0.007 (1.000) |
| | Arc Length | Vessels in macular region | 0.121 (<0.001) | -0.004 (1.000) | 0.051 (<0.001) | 0.034 (<0.001) | 0.022 (0.656) |
| | | Arteries | 0.227 (<0.001) | -0.001 (1.000) | 0.102 (<0.001) | 0.067 (<0.001) | 0.036 (<0.001) |
| | | Veins | 0.193 (<0.001) | 0.004 (1.000) | 0.050 (<0.001) | 0.065 (<0.001) | 0.007 (1.000) |
| | | Nonterminal arteries | 0.222 (<0.001) | 0.000 (1.000) | 0.097 (<0.001) | 0.067 (<0.001) | 0.034 (<0.001) |
| | | Nonterminal veins | 0.189 (<0.001) | 0.005 (1.000) | 0.046 (<0.001) | 0.065 (<0.001) | 0.002 (1.000) |
| | | Terminal arteries | 0.175 (<0.001) | -0.002 (1.000) | 0.087 (<0.001) | 0.048 (<0.001) | 0.031 (0.001) |
| | | Terminal veins | 0.144 (<0.001) | 0.005 (1.000) | 0.042 (<0.001) | 0.045 (<0.001) | 0.012 (1.000) |
| Branching Agnle | Angular Asymmetry | Vessels in macular region | -0.006 (1.000) | 0.002 (1.000) | 0.004 (1.000) | 0.001 (1.000) | 0.011 (1.000) |
| | | Arteries | -0.052 (<0.001) | -0.004 (1.000) | -0.024 (0.218) | -0.013 (1.000) | 0.007 (1.000) |

| | | | | | | | |
|---|---|---|---|---|---|---|---|
| | | Veins | -0.062 (<0.001) | 0.007 (1.000) | -0.027 (0.029) | -0.017 (1.000) | -0.011 (1.000) |
| | Junctional Exponent Deviation | Vessels in macular region | -0.024 (0.203) | -0.010 (1.000) | -0.029 (0.006) | -0.015 (1.000) | 0.015 (1.000) |
| | | Arteries | 0.002 (1.000) | 0.007 (1.000) | -0.050 (<0.001) | -0.011 (1.000) | -0.002 (1.000) |
| | | Veins | 0.028 (0.021) | -0.008 (1.000) | -0.005 (1.000) | 0.002 (1.000) | 0.026 (0.054) |
| | Asymmetry Ratio | Vessels in macular region | 0.002 (1.000) | 0.005 (1.000) | -0.018 (1.000) | 0.011 (1.000) | 0.011 (1.000) |
| | | Arteries | -0.020 (1.000) | -0.005 (1.000) | -0.051 (<0.001) | 0.005 (1.000) | -0.009 (1.000) |
| | | Veins | 0.005 (1.000) | 0.006 (1.000) | -0.013 (1.000) | 0.014 (1.000) | 0.002 (1.000) |
| | Branching Agnle | Vessels in macular region | 0.033 (<0.001) | 0.004 (1.000) | 0.004 (1.000) | 0.014 (1.000) | -0.017 (1.000) |
| | | Arteries | -0.041 (<0.001) | -0.007 (1.000) | -0.035 (<0.001) | -0.004 (1.000) | -0.023 (0.370) |
| | | Veins | 0.028 (0.020) | 0.003 (1.000) | 0.012 (1.000) | 0.015 (1.000) | -0.013 (1.000) |
| | Branching Coefficient | Vessels in macular region | 0.033 (<0.001) | 0.004 (1.000) | 0.024 (0.194) | 0.010 (1.000) | -0.021 (0.667) |
| | | Arteries | 0.004 (1.000) | -0.002 (1.000) | 0.026 (0.057) | 0.003 (1.000) | -0.005 (1.000) |
| | | Veins | -0.004 (1.000) | -0.004 (1.000) | 0.010 (1.000) | -0.009 (1.000) | -0.016 (1.000) |
| | Angular Asymmetry (edge) | Vessels in macular region | 0.019 (1.000) | 0.008 (1.000) | 0.010 (1.000) | 0.005 (1.000) | 0.001 (1.000) |
| | | Arteries | -0.030 (0.004) | -0.015 (1.000) | -0.017 (1.000) | -0.007 (1.000) | 0.007 (1.000) |
| | | Veins | -0.035 (<0.001) | 0.004 (1.000) | -0.009 (1.000) | -0.005 (1.000) | -0.021 (0.957) |
| | Branching Agnle (edge) | Vessels in macular region | 0.010 (1.000) | -0.002 (1.000) | 0.000 (1.000) | 0.007 (1.000) | -0.026 (0.036) |
| | | Arteries | -0.059 (<0.001) | -0.019 (1.000) | -0.037 (<0.001) | -0.002 (1.000) | -0.026 (0.053) |
| | | Veins | -0.030 (0.005) | -0.000 (1.000) | -0.005 (1.000) | -0.002 (1.000) | -0.022 (0.390) |
| Tortuosity | Tortuosity Density | Vessels in macular region | 0.013 (1.000) | -0.011 (1.000) | 0.017 (1.000) | 0.006 (1.000) | -0.009 (1.000) |
| | | Arteries | 0.010 (1.000) | -0.007 (1.000) | 0.022 (0.586) | 0.003 (1.000) | -0.009 (1.000) |
| | | Veins | 0.023 (0.429) | -0.016 (1.000) | 0.018 (1.000) | 0.014 (1.000) | -0.001 (1.000) |
| | Curve Angle | Vessels in macular region | -0.012 (1.000) | -0.004 (1.000) | -0.000 (1.000) | 0.001 (1.000) | -0.020 (1.000) |
| | | Arteries | -0.080 (<0.001) | -0.009 (1.000) | -0.036 (<0.001) | -0.018 (1.000) | -0.040 (<0.001) |

| | | | | | | | |
|---|---|---|---|---|---|---|---|
| | | Veins | -0.029 (0.009) | -0.007 (1.000) | -0.008 (1.000) | 0.001 (1.000) | -0.001 (1.000) |
| | Distance-based Tortuosity | Vessels in macular region | 0.019 (1.000) | -0.002 (1.000) | 0.006 (1.000) | 0.011 (1.000) | -0.022 (0.596) |
| | | Arteries | -0.028 (0.020) | -0.016 (1.000) | -0.003 (1.000) | 0.002 (1.000) | -0.029 (0.005) |
| | | Veins | 0.017 (1.000) | -0.000 (1.000) | 0.002 (1.000) | 0.017 (1.000) | -0.009 (1.000) |
| | Fractal Tortuosity | Vessels in macular region | 0.060 (<0.001) | -0.004 (1.000) | 0.024 (0.171) | 0.004 (1.000) | 0.002 (1.000) |
| | | Arteries | 0.104 (<0.001) | 0.001 (1.000) | 0.047 (<0.001) | 0.027 (0.029) | 0.004 (1.000) |
| | | Veins | 0.086 (<0.001) | 0.003 (1.000) | 0.025 (0.096) | 0.027 (0.024) | 0.015 (1.000) |
| | Linear Regression Tortuosity | Vessels in macular region | 0.014 (1.000) | 0.005 (1.000) | 0.005 (1.000) | 0.005 (1.000) | -0.002 (1.000) |
| | | Arteries | 0.000 (1.000) | -0.005 (1.000) | 0.006 (1.000) | 0.002 (1.000) | -0.015 (1.000) |
| | | Veins | 0.009 (1.000) | 0.006 (1.000) | 0.009 (1.000) | 0.015 (1.000) | -0.001 (1.000) |
| | Angle-based Tortuosity | Vessels in macular region | 0.014 (1.000) | -0.002 (1.000) | -0.005 (1.000) | 0.010 (1.000) | -0.014 (1.000) |
| | | Arteries | -0.007 (1.000) | -0.001 (1.000) | -0.006 (1.000) | 0.003 (1.000) | -0.017 (1.000) |
| | | Veins | 0.014 (1.000) | -0.001 (1.000) | -0.009 (1.000) | 0.014 (1.000) | -0.009 (1.000) |
| | Squared Curvature Tortuosity | Vessels in macular region | 0.016 (1.000) | -0.002 (1.000) | 0.005 (1.000) | -0.001 (1.000) | -0.018 (1.000) |
| | | Arteries | 0.006 (1.000) | -0.010 (1.000) | -0.000 (1.000) | 0.002 (1.000) | -0.018 (1.000) |
| | | Veins | 0.039 (<0.001) | 0.001 (1.000) | 0.011 (1.000) | 0.017 (1.000) | 0.004 (1.000) |
| | Turning Angles | Vessels in macular region | -0.001 (1.000) | -0.011 (1.000) | -0.002 (1.000) | -0.003 (1.000) | -0.005 (1.000) |
| | | Arteries | 0.002 (1.000) | 0.002 (1.000) | 0.003 (1.000) | 0.003 (1.000) | -0.001 (1.000) |
| | | Veins | -0.005 (1.000) | 0.001 (1.000) | -0.004 (1.000) | 0.009 (1.000) | 0.004 (1.000) |
| | Inflection Count Tortuosity | Vessels in macular region | -0.021 (0.834) | -0.009 (1.000) | 0.001 (1.000) | -0.003 (1.000) | -0.004 (1.000) |
| | | Arteries | -0.032 (0.002) | -0.007 (1.000) | -0.005 (1.000) | -0.015 (1.000) | -0.010 (1.000) |
| | | Veins | -0.038 (<0.001) | -0.009 (1.000) | 0.002 (1.000) | -0.002 (1.000) | -0.001 (1.000) |

Notes: The Anderson-Darling test was used for the normality test. If variables were normally distributed, Pearson's correlation coefficient was used, otherwise, Kendall's rank correlation coefficient was calculated.

Supplementary Table 5. Definitions of variables used in the current study

| Variable | Definition and collection | Link |
|---|---|---|
| Age | Age when attended assessment centre | https://biobank.ndph.ox.ac.uk/showcase/field.cgi?id=21003 |
| Sex | Sex of participant. Acquired from central registry at recruitment, but in some cases updated by the participant. Hence this field may contain a mixture of the sex the NHS had recorded for the participant and self-reported sex. | http://biobank.ndph.ox.ac.uk/ukb/field.cgi?id=31 |
| Ethnicity | This is an amalgam of sequential branching questions asked during the initial Assessment Centre visit as part of the touchscreen questionnaire. The question was dropped from the touchscreen protocol on 24/10/2016. | http://biobank.ndph.ox.ac.uk/ukb/field.cgi?id=21000 |
| Indices of Multiple Deprivation | Indices of Multiple Deprivation come from a UK government qualitative study of deprived areas in British local councils. The study is conducted separately in England (IMD), Scotland (SIMD) and Wales (WIMD). Scores were gathered from open data published by the UK government. | https://biobank.ndph.ox.ac.uk/showcase/label.cgi?id=76 |
| Education | ACE touchscreen question \Which of the following qualifications do you have? (You can select more than one)\" The following checks were performed: If code -7 was selected then no additional choices were allowed. If code -3 was selected then no additional choices were allowed. If the participant activated the Help button they were shown the message: - A levels/AS levels and equivalent includes the Higher School Certificate. - O levels/GCSEs and equivalent includes the School Certificate. " High/Intermediate/Low. Education level was classified as follows: high (college or university degree), intermediate (A levels/AS levels or equivalent, O levels/GCSEs or equivalent, CSEs or equivalent, NVQ or HND or HNC or equivalent, and other professional qualifications), and low (none of the aforementioned). | http://biobank.ndph.ox.ac.uk/ukb/field.cgi?id=10722 |
| Body mass index (BMI) | Defined as weight in kilogrammes divided by height in metres squared. BMI value here is constructed from height and weight measured during the initial Assessment Centre visit. Value is not present if either of these readings were omitted. | http://biobank.ndph.ox.ac.uk/ukb/field.cgi?id=21001 |

| | | |
|---|---|---|
| Diabetes | ACE touchscreen question \Has a doctor ever told you that you have diabetes?\" If the participant activated the Help button they were shown the message: If you are unsure if you have been told you had diabetes select Do not know and you will be asked about this by an interviewer later during this visit. " | http://biobank.ndph.ox.ac.uk/ukb/field.cgi?id=2443 |
| Smoking status | This field summarises the current/past smoking status of the participant. Only those who answered "Never" are considered non-smokers | http://biobank.ndph.ox.ac.uk/ukb/field.cgi?id=20116 |
| Intra-ocular pressure | goldmann-correlated intraocular pressure. Units of measurement are mmHg. | https://biobank.ndph.ox.ac.uk/showcase/field.cgi?id=5263; https://biobank.ndph.ox.ac.uk/showcase/field.cgi?id=5255 |
| Visual acuity | final logMAR | https://biobank.ndph.ox.ac.uk/showcase/field.cgi?id=5208; https://biobank.ndph.ox.ac.uk/showcase/field.cgi?id=5201 |
| Low-Density Lipoprotein | Measured by enzyme immunoinhibition analysis on a Beckman Coulter AU5800. Units of measurement are mmol/L. | https://biobank.ndph.ox.ac.uk/showcase/field.cgi?id=30780 |
| High-Density Lipoprotein | Measured by enzyme immunoinhibition analysis on a Beckman Coulter AU5800. Units of measurement are mmol/L. | https://biobank.ndph.ox.ac.uk/showcase/field.cgi?id=30760 |
| Cholesterol | Measured by Choline (CHO) - Pseudocholinesterase (POD) analysis on a Beckman Coulter AU5800 | https://biobank.ndph.ox.ac.uk/showcase/field.cgi?id=30690 |
| glycosylated hemoglobin | Measured by HPLC analysis on a Bio-Rad VARIANT II Turbo | https://biobank.ndph.ox.ac.uk/showcase/field.cgi?id=30750 |
| Systolic Blood Pressure | Blood pressure, automated reading, systolic. Omron device. Units of measurement are mmHg. Measured twice and the mean value was used. | https://biobank.ndph.ox.ac.uk/showcase/field.cgi?id=4080 |
| Diastolic Blood Pressure | Blood pressure, automated reading, diastolic. Omron device. Units of measurement are mmHg. Measured twice and the mean value was used. | https://biobank.ndph.ox.ac.uk/showcase/field.cgi?id=4079 |

| | | |
|---|---|---|
| Cardiovascular Diseases | Prior record of event was determined using data coding "1065, 1066, 1067, 1068, 1072, 1074, 1075, 1076, 1077, 1078, 1-79, 1080, 1081, 1082, 1083, 1086, 1087, 1088, 1093, 1094" from Field ID 20002 for cardiovascular diseases, including hypertension, heart or cardiac problems, peripheral vascular disease, venous thromboembolic disease, essential hypertension, gestational hypertension or pre-eclampsia, angina, heart attack or myocardial infarction, heart failure or pulmonary edema, heart arrhythmia, heart valve problem or heart murmur, cardiomyopathy, pericardial problem, stroke, transient ischemic attack, subdural hemorrhage or hematoma, subarachnoid hemorrhage, leg claudication or intermittent claudication, arterial embolism, pulmonary embolism with or without deep vein thrombosis, deep vein thrombosis.<br>In addition, those reported vascular/heart problems at interview: ACE touchscreen question \Has a doctor ever told you that you have had any of the following conditions? | https://biobank.ndph.ox.ac.uk/showcase/field.cgi?id=20002; http://biobank.ndph.ox.ac.uk/ukb/field.cgi?id=6150; https://biobank.ndph.ox.ac.uk/showcase/coding.cgi?id=6 |
| Neurodegenerative Diseases | Prior record of event was determined using data coding "1258, 1259, 1260, 1261, 1262, 1263, 1397" from Field ID 20002 for diseases, including chronic/degenerative neurological problem, motor neurone disease, myasthenia gravis, multiple sclerosis, Parkinson's disease, dementia/Alzheimer's/cognitive impairment, other demyelinating disease (not multiple sclerosis). | https://biobank.ndph.ox.ac.uk/showcase/field.cgi?id=20002 https://biobank.ndph.ox.ac.uk/showcase/coding.cgi?id=6 |
| Ocular Diseases | Prior record of event was determined using data coding "1274, 1275, 1276, 1277, 1278, 1279, 1281, 1282, 1527, 1528" from Field ID 20002 for eye infection, retinal problem, diabetic eye disease, glaucoma, cataract, eye trauma, retinal detachment, retinal artery/vein occlusion, retinitis pigmentosa, macular degeneration.<br>In addition, those reported eye problems at interview: ACE touchscreen question \Has a doctor ever told you that you have had any of the following conditions? | https://biobank.ndph.ox.ac.uk/showcase/field.cgi?id=20002; http://biobank.ndph.ox.ac.uk/ukb/field.cgi?id=6148; https://biobank.ndph.ox.ac.uk/showcase/coding.cgi?id=6 |

Supplementary Table 6. Definitions of measure types

| Category | Measure Type | Definition |
|---|---|---|
| Tortuosity | Turning angles | a measure of the distribution of angles between successive line segments along the curve. A higher mean, higher range indicates a more tortuous curve |
| | Linear regression tortuosity | calculates a tortuosity measure by estimating a line that start and ends with the first and last points of the given curve, then samples a number of pixels from the given line and calculates its determination coefficient, if this value is closer to 1, then the given curve is similar to a line |
| | Angle based tortuosity | A measure of tortuosity based on the local direction variation of the vessel, in which the average of the angles between consecutive vessel samples is computed. |
| | Squared curvature tortuosity | Squared curvature tortuosity analyzes the intensity of vessel bends at each point, squaring these values to emphasize sharpness, and then summing them along the entire segement. |
| | Distance-based Tortuosity | ratio of the total vessel arc length to the distance between endpoints |
| | Inflection Count Tortuosity | Calculated by counting the number of inflection points along the vessel.This method detects the inflection points of a given curve y=f(x) by applying a convolution to the y values and checking for changes in the sign of this convolution, each sign change is interpreted as an inflection point. |
| | Fractal tortuosity | the Minkowski–Bouligand dimension of the vessel segment |
| | Tortuosity density | this measure evaluates vessel tortuosity by summing local contributions to tortuosity by assessing how much each turn curve is different from a smooth curve |
| | Curve angle | the mean segment angles between each branch sampled at a length of 10 pixels |
| Complexity | Number | number of vascular segments, or vascular trees, or bifurcation and branching points, or terminal and non-terminal points |
| | Level | number of segments passing by an endpoint/branching/bifurcation point |
| | Strahler | a numerical measure of its branching complexity using Strahler number |
| | Fractal Dimension | calculated by the box-counting method, expressing the density and overall complexity of retinal vasculature in one number |

| | | |
|---|---|---|
| Density | Vessel Area Density | ratio of the area occupied by vessels divided by the total area |
| | Vessel Skeleton Density | the ratio of the total length of vessels to the total area of the image at 1 pixel, a measure of overall vessel length within a fundus image |
| | Branching Density | number of identified branchpoints divided by total vessel length |
| | Bifurcation Density | number of identified bifurcations divided by total vessel length |
| | Arc length | the distance between two points along a section of a vessel curve |
| | Chord length | the line segment joining any two points on the circumference of a vessel curve |
| Caliber | Mean width of all segments | mean value of diameters of all identified vessel segments |
| | Length Diameter Ratio | the ratio of the length between two branching points to the trunk vessel width |
| | CRAE/CRVE | the summarization of the vessel calibers from the six largest arteries/veins in the Standard Zone |
| | AVRe | a ratio calculated from CRAE/CRVE within the standard zone |
| Branching Angle | Asymmetry Ratio | (min (d1, d2)/max(d1/d2)) ^2, d1 and d2 denote diameters of the daughters of the root segment of vessel |
| | Angular Asymmetry | the absolute difference between the angles of each daughter vessel with the root vessel |
| | Branching Coefficient | (d1+d2) ^2/d0^2, d0 = parent vessel diameter (px), d1 = daughter's vessel diamater (px), d2 = diameter of daughter vessel two (μm), it explains the relationship between the calibers of the parent vessel compared to the daughter vessels at a bifurcation. |
| | Junctional Exponent Deviation | the extent to which the relationship between the diameter of the parent vessel and the daughter vessels deviates from theoretically defined optimum (Murray's law). |
| | Branching Angle | the angles between the sampled centerline of the root segments of vessel and its daughter segments s1 and s2 near the branching point. Murray proposed in 1926, the optimal arteriolar branching angle to be 75 degrees, and any deviation from the optimal angle was seen as less optimal for the retinal circulatory system. |
